# A systematic framework for assessing the clinical impact of polygenic risk scores

**DOI:** 10.1101/2020.04.06.20055574

**Authors:** Scott Kulm, Andrew Marderstein, Jason Mezey, Olivier Elemento

**Affiliations:** Department of Physiology and Biophysics, Weill Cornell Medicine, New York, NY; Caryl and Israel Englander Institute of Precision Medicine, Weill Cornell Medicine, New York, NY; Physiology, Biophysics and Systems Biology Graduate Program, Weill Cornell Medicine, New York, NY; Department of Genetic Medicine, Weill Cornell Medicine, New York, NY; Department of Computational Biology, Cornell University, Ithaca, NY

## Abstract

Risk prediction models provide empirical recommendations that ultimately aim to deliver optimal patient outcomes. Genetic information, in the form of a polygenic risk score (PRS), may be included in these models to significantly increase their accuracy. Several analyses of PRS accuracy have been completed, nearly all focus on only a few diseases and report limited statistics. This narrow approach has limited our ability to assess as a whole whether PRSs can provide actionable disease predictions. This investigation aims to address this uncertainty by comprehensively analyzing 23 diseases within the UK Biobank. Our results show that including the PRS to a base model containing age, sex and the top ten genetic principal components significantly improves prediction accuracy, as measured by ROC curves, in a majority 21 of 23 diseases and reclassifies on average 68% of the individuals in the top 5% risk group. For heart failure, breast cancer, prostate cancer and gout, decision curve analyses using the 5% risk threshold determined that including the PRS in the base model would correctly identity at least 60 more individuals who develop the disease for every 1000 individuals screened, without making any incorrect predictions. Analyses that included disease-specific risk factors, such as Body-Mass Index, and consider time of disease onset found similar PRS benefits. The improved prediction accuracy was translated to 10 instances in which medications/supplements and 94 instances in which lifestyle modifications lead to significantly greater reduction in disease risk for individuals in the top PRS quintile compared to the bottom PRS quintile. Finally we provide guidance for tailored, future PRS generation by comprehensively ranking methods that generate PRS weights and identifying genome wide association study characteristics that influence PRS predictions. The unification of significantly enhanced disease predictions, novel risk mitigation opportunities and improved methodological clarity indicate that PRSs carry far greater clinical impact than previously known.

## Introduction

Stratifying a population’s disease risk can greatly improve public health. Individuals with high risk can receive preventative care and those at low risk can avoid unnecessary tests and procedures^1^. Many clinical risk models utilize age, sex and pre-existing conditions to form predictions that shape preventative recommendations^2^. These models nearly universally exclude genetic information, despite evidence that it may significantly increase prediction accuracy^3^. The exception to this rule is specific gene mutations. For example, deleterious BRCA mutations significantly increase risk of breast cancer, and therefore mutation carriers are advised to start mammogram screening earlier in life^4^. However, the heritable component for the vast majority of traits is not captured by a few relatively rare mutations, but rather is determined by more common variants scattered across the entire genome^5^. Polygenic risk scores (PRSs) condense this diffuse information into one metric, by summing the product between a genetic variant’s externally determined effect size and its number of alternative alleles, over multiple genetic locations. We now know that resultant PRSs are as predictive as clinically relevant monogenic mutations, such as BRCA^6^, while affecting a sometimes much greater number of individuals. For many diseases, it is hoped that these risk predictions may ultimately translated into improved patient outcomes. PRSs often act independent from classical clinical and demographic risk factors, and in a small number of cases have been shown to identify high-risk groups that experience maximal benefit from a drug or lifestyle change^7^. While relevant investigations to date have shown the great potential of individual PRSs in specific diseases, there has been limited exploration of their predictive potential across many diseases, no general assessment of candidate strategies that may modulate high risk and poor accepted processes for score generation. As a result, uncertainties remain around the extent to which PRSs can generally provide clinical benefit.

For example, studies involving coronary artery disease PRSs have achieved conflicting results. One study within the UK Biobank found that individuals within the top 8% of the PRS distribution carried threefold more risk than all remaining individuals^6^. Whereas, another study within the Atherosclerosis Risk in Communities cohort found that the addition of a PRS to a model containing a specific set of common clinical factors decreased the model’s C-statistic by 0.001^8^. These differing results likely do not signify fundamental deficiencies within the theoretical basis of PRSs, but rather underscore how slight changes in methods and models can lead to vastly different interpretations. The systematic assessment and comparison of PRS methods across many diseases may greatly reduce this variability in PRS perception. The conclusions drawn from such a comprehensive approach will likely better reflect the intrinsic worth of the PRS.

A PRS with high predictive accuracy is necessary but not sufficient for impacting clinical care. Translation from prediction to outcomes requires a recommendation connecting a strata of individuals to an evidence-backed action. Some publications provide this evidence, for example individuals with a high coronary artery disease PRS saw a greater risk reduction with the use of statins as compared to low scorers^7^. However, most do not, thereby obscuring the true utility of the related PRS.

The difficulties associated with PRS analysis compound when deriving the score directly from genome wide association study (GWAS) results, rather from a pre-compiled set of genetic variants. Over a dozen PRS generative methods exist, each indicating their own superiority^9–11^. The absence of a well-documented, unbiased evaluation of these methods leads to sub-optimal PRS generation and ultimately a diminished ability to identify at-risk individuals in clinical settings.

These shortcomings in the wider PRS field complicate clear conclusions concerning PRS use in the clinic. To remedy this problem, our investigation has rigorously analyzed PRSs of 23 diseases within the 500,000 person strong UK Biobank^12^ (Figure 1). Specifically, we began by applying all 15 known and available methods^9–11,13–22^ to each disease’s GWAS summary statistics (Table M7, M8, M2). From all the scores created for a single disease, cross-validation within the training-phase of the UK Biobank determined the single best score. ICD codes and self-reports determined disease status (Table M3-M4), and the individuals under analysis were of White, British ancestry. The best score for each disease was fully assessed in the withheld testing phase; both logistic regression and Fine and Gray models were fit, from which several accuracy statistics were derived. Additional clinical factors both included in the models and used to stratify the model predictions further expanded the statistics assessed (Table M5-M6).

**Figure 1.**
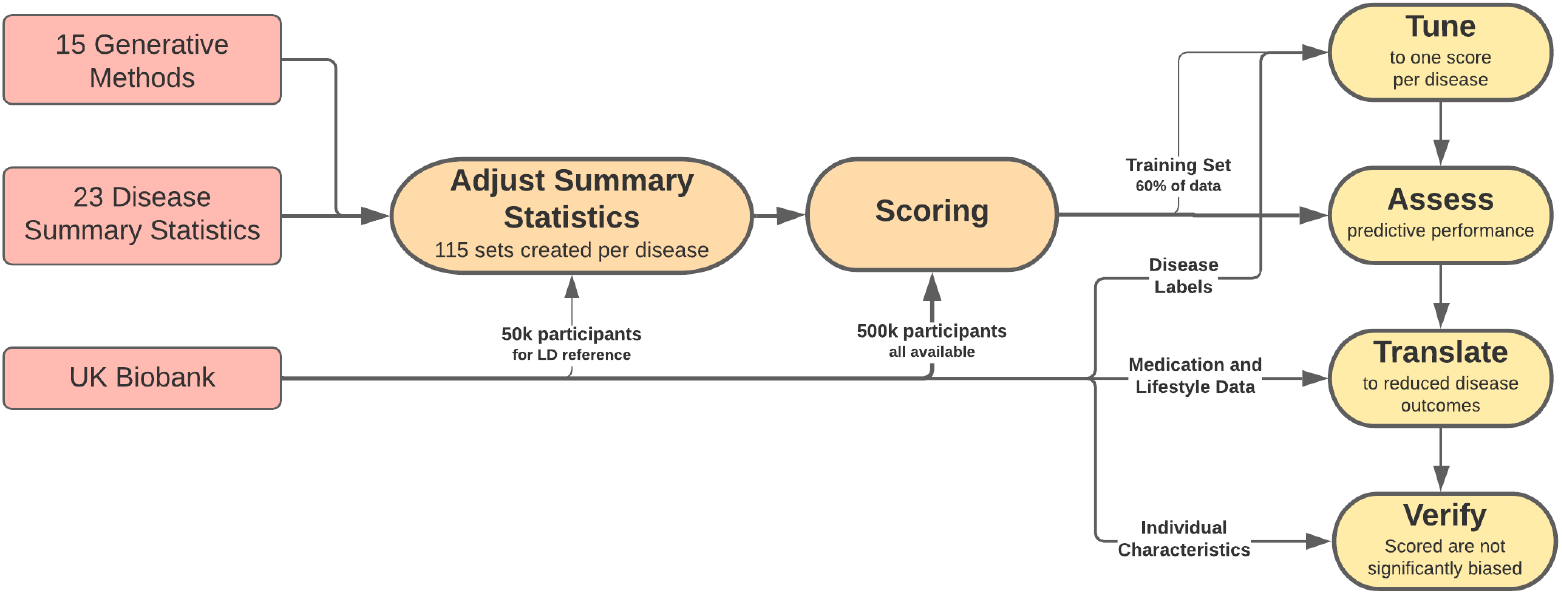
Flow chart of the process utilized in this investigation. On the left side, in red boxes, the necessary data is first gathered. Both the summary statistics and UK Biobank data were thoroughly quality controlled. In the middle, in the orange boxes, the intensive computations that create the polygenic risk scores were completed. Specifically, the summary statistics were adjusted by each of the 15 methods and these updated weights were then used to create the PRSs. On the right side, in the yellow boxes, the PRSs are comprehensively analyzed. First, all PRSs were evaluated in a training phase of the data, next the best PRS for each disease was fully assessed in a testing phase of the data, followed by attempts to translate the PRS predictions through clinical actions to reduced risk, and lastly verification of limited PRS bias. An extensive description of the methods utilized are provided in Supplementary II.

To uncover risk modulation strategies, we grouped individuals according to both lifestyle measurements, such as time spent watching TV or exercising, and medication/supplement consumption status (Table M10). The absolute risk reduction associated with moving from one lifestyle group or medication status to another were computed with respect to a sub-sample of individuals with either low, intermediate or high PRSs. To investigate possible PRS bias, accuracy statistics derived from groups split by individuals characteristics were compared. Lastly, the accuracy of PRSs derived from all generative methods were compared directly and weighted by GWAS characteristics, such as the total number of variants, to produce comprehensive guidance on PRS creation and analysis. The results derived from this investigation indicate that PRSs carry significant predictive capacity with limited drawbacks when modeled correctly.

## Results

### Assessing Predictive Performance

To determine whether each PRS has predictive value within a clinical risk model we assessed its ability to predict the onset of disease. For each disease, we generated multiple PRSs by applying 15 generative methods to GWAS summary statistics. Each PRS was assessed through cross-validation in the 246,062 person strong training-phase. Addition of the PRS with the highest Area Under the receiver operator Curve (AUC) to the basic model, consisting of age, sex and the top ten genetic principal components, formed the PRS-included model. Both model types were fit upon the training-phase and assessed on the withheld, 162,119 person strong testing-phase. The AUC of the PRS-included model ranged from 0.568 (NAFLD) to 0.817 (Heart Failure). An AUC of 0.5 signifies a set of random predictions and an AUC of 1.0 signifies a set of perfect predictions. The metric of AUC improvement, or the difference in AUC generated from the PRS-included and basic model, ranged from 0.0040 (Stroke) to 0.146 (Ulcerative Colitis)(Table 1). Comparison of each disease’s basic and PRS-included model through the DeLong Test generated 21 p-values that were less than 0.0001, indicating an overwhelming number of diseases (21 of 23) are predicted significantly better when the PRS is included (Table S1). Interestingly, the correlation between AUC and AUC improvement was only 0.10 (p = 0.643) (Table S2). The divergence between these statistics is likely related to the average age of onset of each disease, with AUC improvement better representing the independent worth of the PRS.

**Table 1.**
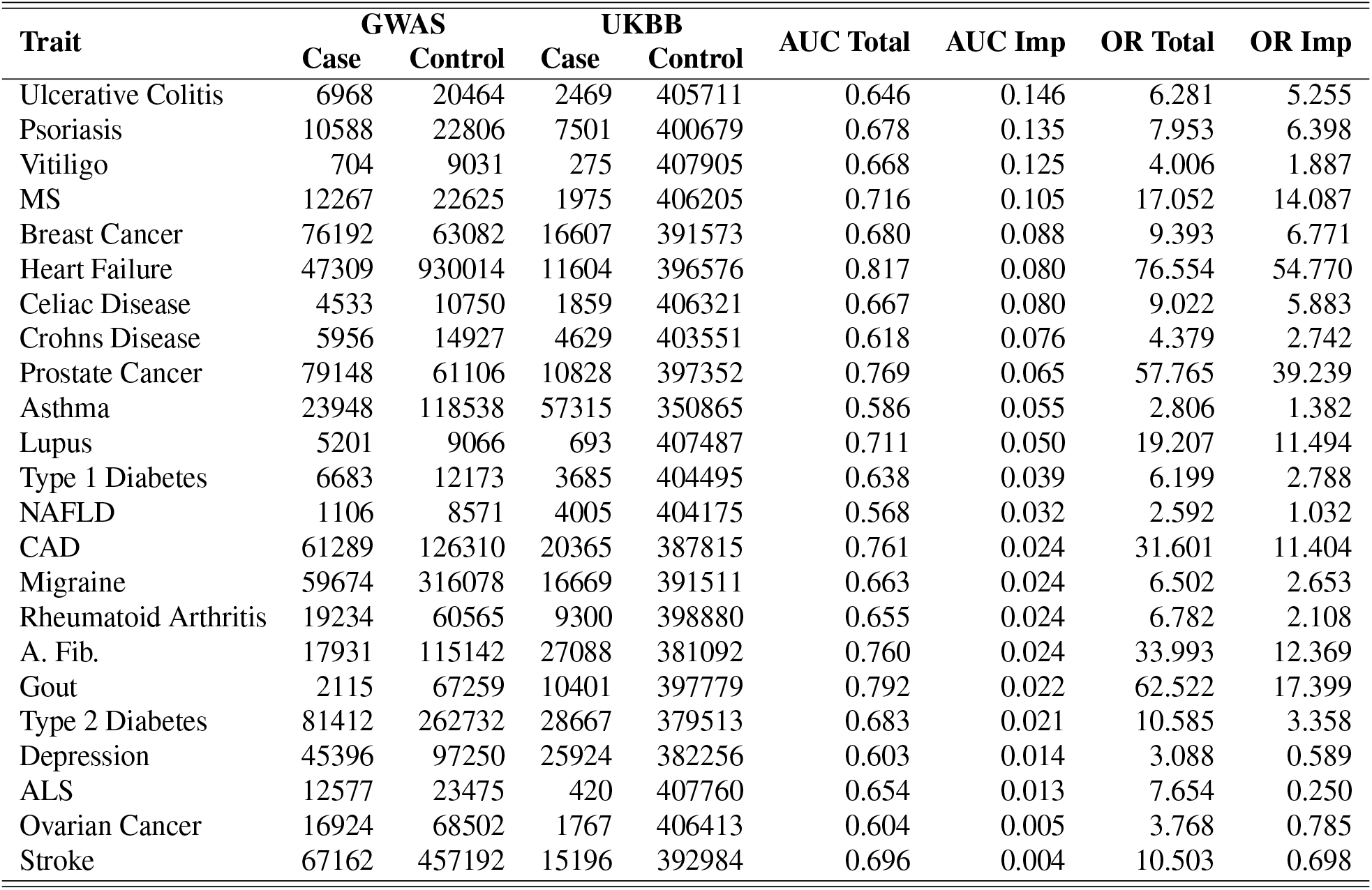
Predictive Performance Statistics for All Diseases. For each disease, the first four columns list case/control information for both the originating GWAS that created the summary statistics, and the UK Biobank. The remaining columns provide the AUC and odds ratio values generated from models fit on the training data and used to predict the testing data. Specifically, the total statistics are derived from model containing age, sex, top ten genetic principal components and the respective PRS. The improvement statistics are the difference between the total statistics and the statistic generated from models containing age, sex, and top ten genetic principal components. Odds ratios were calculated by establishing a 2×2 contingency table in which individuals predicted to develop the disease retained a model prediction greater than the 95^*th*^ percentile of predictions and non-predicted individuals retained a model prediction less than the 20^*th*^ percentile of predictions. Disease abbreviations are defined in Table M1.

While the AUC statistic captures overall model fit, odds ratio better characterizes the high risk strata - the group most easily targeted for intervention. We therefore generated odds ratios by constructing a 2×2 table in which the individuals predicted to develop the disease retain a model prediction above a specified percentile, and non-predicted individuals retain a model prediction below the 20^*th*^ percentile. At the 95^*th*^ percentile cut-off, with risk determined by the PRS-included model, odds ratios ranged from 2.59 (NAFLD) to 76.6 (Heart Failure)(Table 1). If the base model is alternatively used, the odds for heart failure drops to 21.8, an odds ratio improvement of 54.8. To put these values in perspective, the odds ratio of BMI upon heart failure is 2.0^23^, and the odds ratio of smoking upon lung cancer is 40^24^. As both of these risks are considered clinically important, and PRSs carry risk of a similar magnitude, it would be logical to conclude that PRSs have clinical value.

The ability of PRSs to accurately identify high risk individuals was additionally analyzed by enumerating the individuals reclassified as high risk when a base model includes a PRS. In this analysis, individuals above and below the 95^*th*^ percentile of risk are defined to be high-risk and low-risk, respectively. On average 68.5% (SE 3.4%) of high-risk individuals are classified into a lower risk group when replacing the PRS-included model predictions with base model predictions (Figure 2). For example, ulcerative colitis would see 90.3% of individuals in its high-risk group re-classified as low-risk when the PRS is removed from the model (Table S3). To extend this analysis the net reclassification index statistic^25^ (NRI) was computed, providing a means to determine whether the magnitude of reclassification reflects meaningful changes or random reshuffling. The formal definition of NRI is: the difference between the proportion of individuals who develop the disease and move into and out of the high risk group, minus the difference between the proportion of individuals who do not develop the disease and move into and out of the high risk group. The mean NRI was 9.48% (SE 1.77%), and NRI values for all but two diseases indicate that inclusion of the PRS significantly improves disease classification (Table S4). We also computed the Brier Scores and integrated discrimination indices to further show that PRSs improve disease classification in the high-risk strata (Table S5-S6).

**Figure 2.**
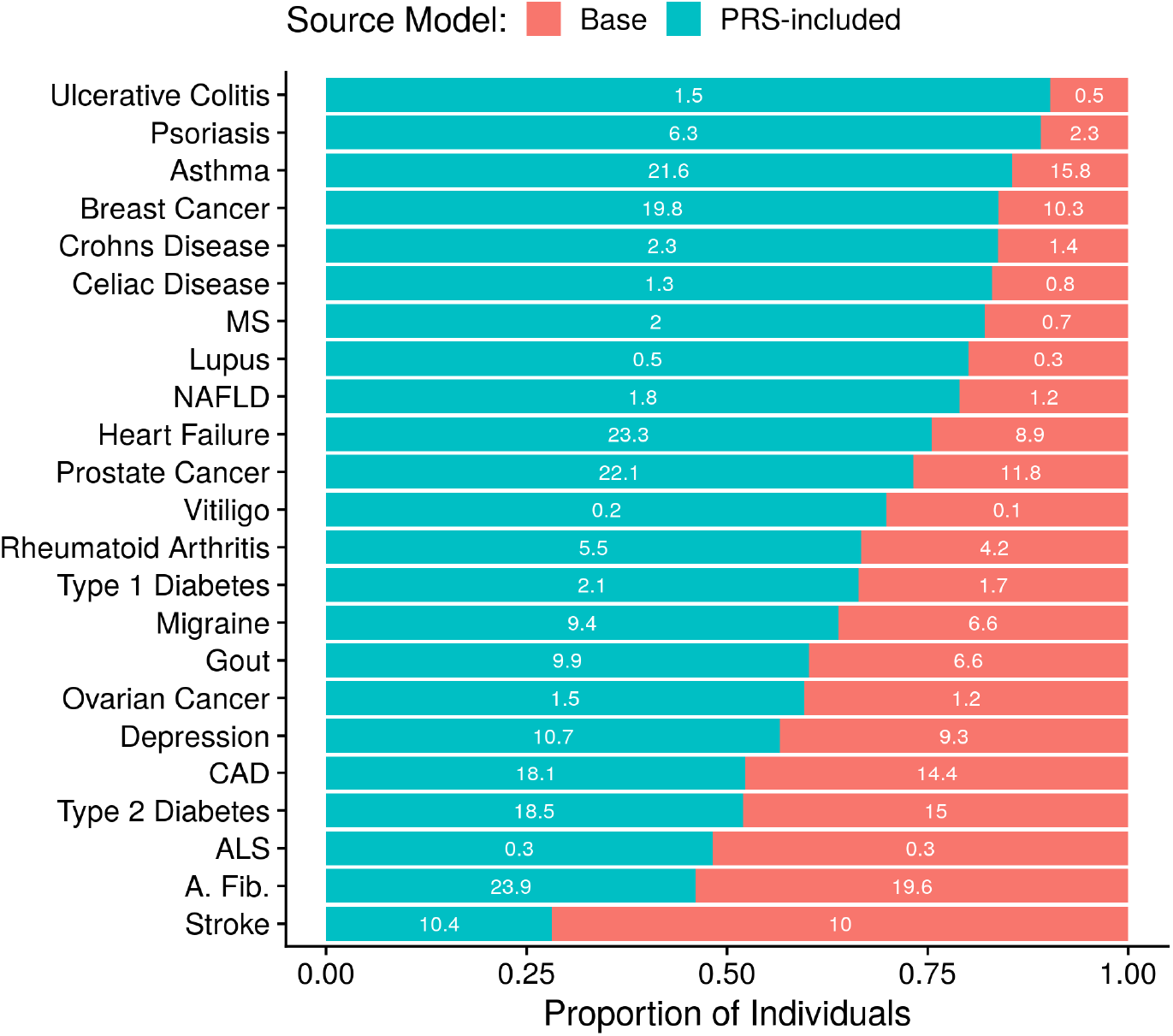
Reclassification of Individuals into High Risk Groups and the Corresponding True Positive Rates. For each disease, the individuals in the top 5% of risk as determined by a model containing age, sex, top ten genetic principal components and respective PRS are separated into classification groups. The Base group counts the proportion of individuals who would remain in the risk group if the PRS was removed from the model, the Score Incl. group counts the proportion of individuals that would be reclassified downward if the PRS was removed. The number within each group is the respective true positive rate under the assumption that all individuals in the top 5% of model predictions risk are cases.

As the NRI has been criticized^26^, we carried out a final examination of high risk individuals by comparing true positive rates. In this analysis, individuals are predicted to develop the disease if their risk exceeds the 95^*th*^ percentile. Shifting from the base to PRS-included model increased the average true positive rate by 65.4% (SE 12.4%). For eleven diseases the improvement in true positive rates exceeded 50%. One specific example, breast cancer, corresponds to a true positive rate of 10.2% when using the base model and 19.8% when including the PRS, an increase of 92.5% (Table S7). This clear improvement in accuracy, common across all statistics reported, implies that the inclusion of a PRS in a clinical risk model may identify many more individuals that will develop a disease while reducing the number of individuals falsely labeled as high-risk.

### Expanded Forms of Polygenic Risk Score Analyses

In addition to the logistic regression results obtained thus far, we carried out expanded forms of analyses to get a more comprehensive understanding of each PRS’s predictive ability. First, we fit Fine and Gray models, which adjusts for death as a competing outcome, to better study the time of disease onset. Repeating the logistic regression model assessment, the addition of a PRS to the basic model, consisting of age, sex and the top ten genetic principal components, formed the PRS-included model. These time-incorporated models were slightly complicated by mixing of electronic health record diagnoses, which occurred from 1999-2020 and self-assessment diagnoses, which occurred from 2006-2010. Nevertheless, the difference in the final time-point cumulative hazard between the base and PRS-included models could still be computed. (Figure 3A). For example, with respect to asthma, the addition of the PRS to the base model increased the cumulative hazard by 0.04 for individuals in the high-risk group defined by the 5^*th*^ quintile of risk. The same model comparison led to a cumulative hazard drop of 0.026 within the low-risk group defined by the 1^*st*^ quintile of risk. (Figure 3B). These gain and drop patterns are mirrored in many other diseases (Tables S8-S9). Since cumulative hazards, as compared to odds ratios, better reflect associations to time of disease onset, these results indicate that models including PRSs generate robust predictions that achieve high accuracy in multiple analytic regimes.

**Figure 3.**
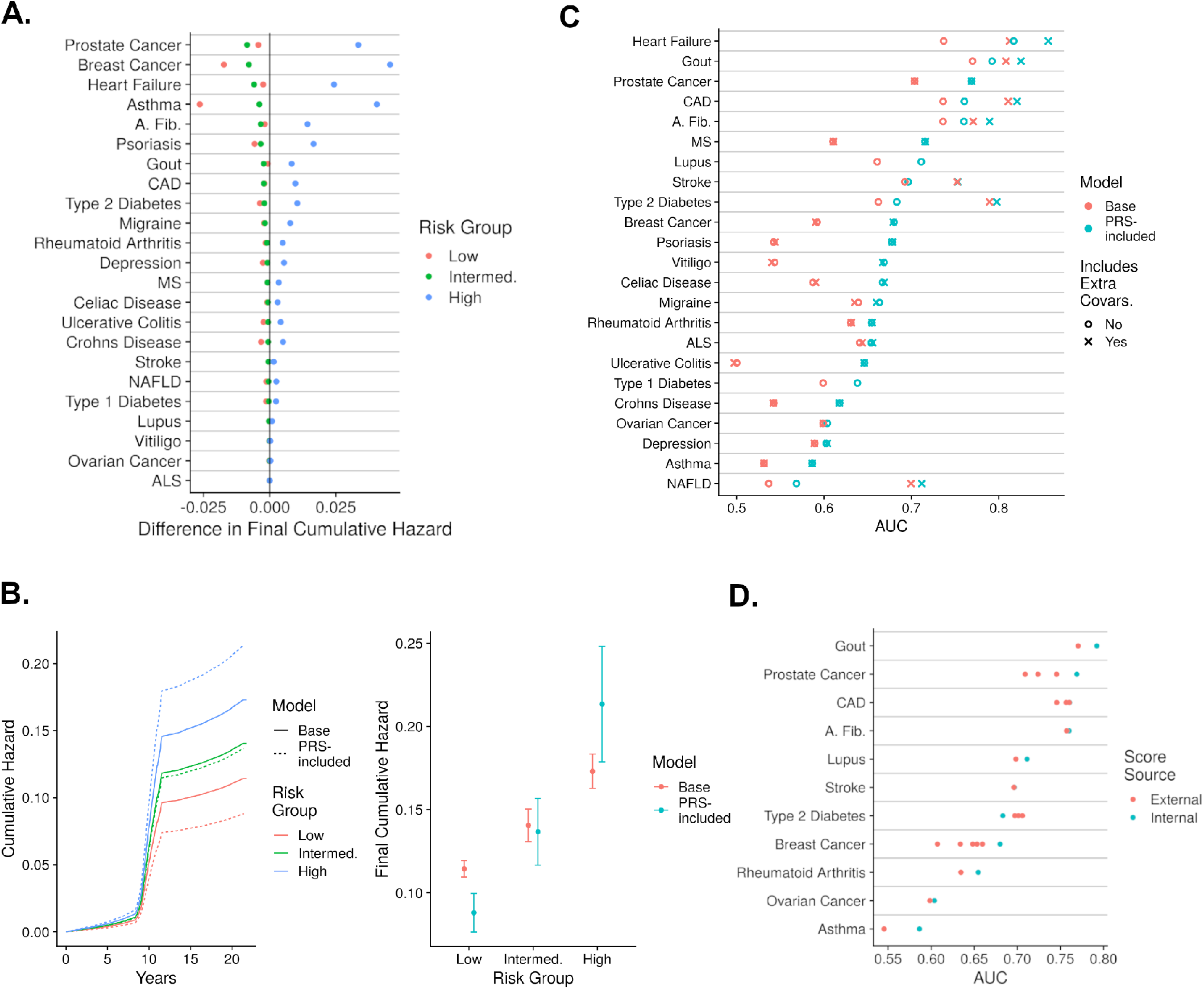
Assessment of PRS predictive performance for 23 diseases. A: Difference of cumulative hazard at the final time-point in the study generated from Fine and Gray models that either contain just age, sex,and the top ten principal components, or also include the respective PRS. These hazards are broken into three risk groups, the high risk group are individuals in the top quintile of hazards, the low risk group are individuals in the bottom quintile of hazards, and intermediate risk group are the remaining individuals. B: Analysis of Asthma using the three risk groups and the two model types described in panel A. The left sub-figure describes the cumulative hazard increase throughout time, and the right sub-figure describes the cumulative hazards at the final time-point and their standard errors. C: The AUC values generated from models that include age, sex, the top ten genetic principal components (base), and optionally either the PRS (score), and/or extra relevant covariates (yes to includes extra covariates). D: The AUCs generated from models that include age, sex, top ten principal components and either an internal PRS analyzed previously or an external PRS whose variant weights were extracted from the PGS Catalog.

Second, additional clinical covariates were considered within the base model to provide a more realistic estimate of a PRS’s clinical value. For example, a base model predicting gout with the covariates of age, sex and the top ten genetic principal components generated an AUC of 0.770. The base model’s AUC increased by 0.017 to 0.825 by including the PRS. An extra-covariate model containing the base model’s covariates, obesity, hypertension and diabetes generated an AUC 0.808. The extra-covariate model’s AUC increased by 0.017 to 0.825 by including the PRS. This 0.017 increase in AUC indicates that the PRS has predictive value, although this value is relatively diminished by the extra covariates since the base model’s AUC increases by 0.022 by including the PRS. Amongst the 21 diseases with relevant extra covariates, their inclusion to the the PRS-included model increased the total AUC by an average of 0.023 (SE 0.0089) compared to the base model (Figure 3C, Table S10). Therefore, consideration of extra covariates may diminish the effect of the PRS but in no way completely eliminates their predictive value.

Third, externally generated PRSs listed in the PGS Catalog^27^ were compared to internally generated PRSs derived from GWAS summary statistics and described thus far. Amongst the 11 diseases with internal and external scores, only Type II diabetes saw higher predictive performance from an external score (AUC increase of 0.023). On average, the AUCs generated from models with the internally developed PRS were 0.011 (SE 0.0050) greater than models with external PRSs (Figure 3D, Table S12). Altogether, these expanded analyses indicate that the our PRSs are well-suited to various modelling approaches, and therefore, can likely be readily adapted to various types of clinical applications.

### Translating Polygenic Risk Scores to Clinical Benefit

We then attempted to quantify the potential clinical benefit of PRS predictions. We first use decision curve analyses to consider the cost of incorrect predictions. Specifically, net benefit values, which represent the proportion of true positives found without making any false predictions, formed the decision curves. Each net benefit corresponds to a threshold. Individuals with a risk prediction greater than the threshold are assumed to develop the disease. If the net benefit improvement, or the difference between base and PRS-included models’ net benefits, is positive then the PRS provides a clinical benefit at that threshold. For example, the net-benefit for atrial fibrillation under the base model was 0.0086 when using the 89^*th*^ percentile of risk as the threshold. Meaning that out of 100,000 assessed, 86 would be correctly classified as developing atrial fibrillation without making a false prediction. Under the PRS-included model and the same threshold the net benefit increased by 44.2% to 0.0124. Meaning that for every 100,000 individuals assessed 124 would be correctly classified as developing atrial fibrillation without making a false prediction - 38 more than with the base model^28^. Across all diseases the net benefit improvement was positive for at least one threshold (Table S12). At the common threshold of the 95^*th*^ risk percentile, 13 diseases saw a positive net benefit improvement. At the threshold of the 99^*th*^ risk percentile, a different six disease saw a positive net benefit improvement. Therefore, different diseases require different thresholds for the PRS to provide a predictive benefit (Figure 4A, Table S13). Eight diseases had a range of thresholds greater than 10 risk percentiles in which the net benefit difference was positive. For example, prostate cancer’s net benefit improvement was positive for all thresholds between the 66^*th*^ and 99^*th*^ risk percentiles. The other diseases include heart failure, prostate cancer, and type 2 diabetes - all severe but with modifiable risk factors (Table S14). Therefore, for the most harmful diseases even a poorly threshold that does not maximize the net benefit improvement can empower decisions that are more clinically beneficial than cautious approaches that exclude the PRS altogether.

**Figure 4.**
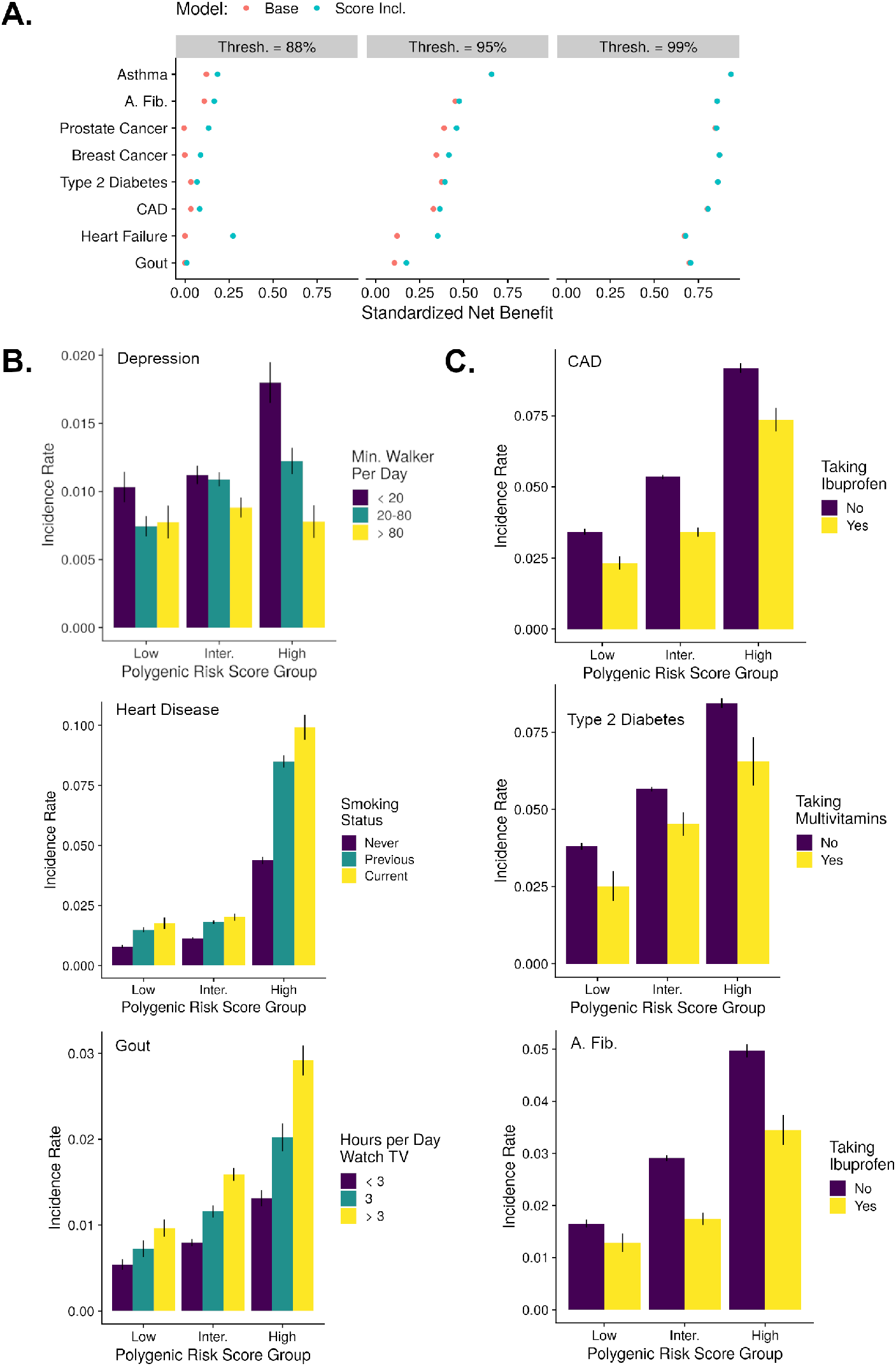
Translation of PRS Accuracy to Clinical Utility. A: For a selection of diseases, the net benefits, standardized by disease prevalence, corresponding to three thresholds are shown. The threshold indicates the proportion of top risk individuals that are predicted to develop the disease. B: The net incidence of disease within groups defined by PRS and a lifestyle feature. Low, intermediate and high PRS groups correspond to the 1st, 2nd-4th, and 5th quintiles, respectively. C: The net incidence of disease within groups defined by PRS and a medication/supplement status. The PRS groups maintained the same definitions as in the lifestyle analysis.

To further the translation of improved decisions to outcomes, we sought to identify specific subsequent actions that may significantly and specifically aid individuals with high PRSs. First, we considered lifestyle modifications, such as an improved diet or increased frequency of exercise. The absolute risk was computed within groups of individuals with specified lifestyle values and either low, intermediate, or high PRS, defined as the 1^*st*^, 2^*nd*^-4^*th*^ and 5^*th*^ quintiles, respectively. In this analysis, as compared to all previous analyses, the PRS was directly used instead of model predictions so the results may align with previous, related analyses^7,29^. To limit confounding, each PRS was adjusted for age, sex and the top ten genetic principal components. We found 94 instances in which the risk reduction, or the difference in risk associated with high and low lifestyle groups, was both greater in the high PRS group as compared to the low PRS group and significant in at least one PRS group (Fisher Exact Test P-Value < 5 *×* 10^*−*4^). For example, with respect to depression, individuals in the high PRS risk group see an absolute risk reduction of 1.0% (SE 0.19%) when increasing the minutes walked per day from less than 20 to over 80, whereas those in the bottom PRS risk group only see an absolute risk reduction of 0.26% (SE 0.16%) (Figure 4B). Therefore, from the same lifestyle modification individuals in the high PRS group see a 284% greater risk reduction as compared to individuals in the low PRS groups. In another example, individuals experience an absolute risk reduction from heart disease of 2.58% and 0.45% in the high and low PRS groups, respectively, when going from current to previous smokers. Lastly, individuals experience an absolute risk reduction from Gout of 1.60% and 0.42% in the high and low PRS groups, respectively, when going from more than 3 to less than 3 hours of TV watched per day. Similar differences in risk were produced by the lifestyle factors processed meat intake, walking pace, alcohol consumption, sleep duration and daily driving time (Table S15, S16). By identifying individuals with high PRS, application of lifestyle modifications can lead to relatively large risk reductions.

Instances in which clinical actions may associate PRS predictions with enhanced care were again sought by evaluating medications and supplements. Similar to before, the absolute risk was computed within groups of individuals that either do or do not take the medication/supplement and hold either low, intermediate or high adjusted PRS. We find 10 instances in which the absolute risk reduction brought by taking a medications or supplement was greater in the high PRS group compared to the low PRS group, and the effect of the medication or supplement was significant. For example, with respect to atrial fibrillation, individuals in the high PRS risk group see an absolute risk reduction of 1.94% (SE 0.44%) when taking ibuprofen, whereas those in the bottom PRS risk group only see an absolute risk reduction of 1.10% (SE 0.26%) (Figure 4C). Alternatively, individuals experienced an absolute risk reduction from type 2 diabetes of 1.30% and 1.11% in the high and low PRS groups, respectively, when taking multivitamins. While the prevalence of non-clinically validated supplements in these findings underscore their prospective nature (Table S17, S18), other reports of significant medication-PRS associations promotes their general potential^7,30,31^. The totality of the medication, lifestyle and decision curve results illustrate a potential pathway from accurate PRSs to improved clinical decisions with many choices of effective clinical actions resulting in realized risk reduction.

### Modifications of Polygenic Risk Score Accuracy by Individual characteristics

Polygenic risk scoress should ideally function with equal accuracy within any sample from a larger population. However, the effects from individual characteristics, such as age, income and home location, may break this ideal assumption. To determine if these characteristics could significantly bias PRS predictions, we split the larger testing sample into several pairs of groupings according to various individual characteristics. Within each group, models were independently fit then assessed to determine PRS accuracy. First, male and female groupings were considered for all diseases except for breast, prostate and ovarian cancers. The mean AUC difference between the sexes was 2.18 *×* 10^*−*3^ (SE 5.06 *×* 10^*−*3^) with male generated AUCs besting female in 11 diseases. Four pairs of AUC values were significantly different between the sexes, the most significant among them was Gout (p = 8.22 *×* 10^*−*7^, AUC difference = 0.045) (Table S19). Second, young and old groupings formed, defined by the median age of the disease cases (Table M11). The mean AUC difference between age groups was 0.0325 (SE 6.46 *×* 10^*−*3^), with AUCs generated in the younger group besting the older group in 21 diseases. Ten AUC pairs were significantly different, with prostate cancer and atrial fibrillation both having p-values less than 1 *×* 10^*−*50^ (AUC difference 0.118 and 0.086, respectively) (Table S20). The ability to detect disease at a relatively early age of onset is likely clinically more difficult, therefore, the observed predictive bias of PRSs towards younger individuals may in fact be beneficial.

Next, we used socially-focused individual characteristics, such as income and education, to similarly investigate potential PRS bias. A common trend of significant differences between groups emerged for the top five performing traits: heart failure, gout, CAD, prostate cancer and atrial fibrillation. Specifically, individuals with either income less than £ 40,000, fewer than 3 persons in their household or time at their current address exceeding 20 years generated AUCs greater than individuals with each of the opposite possible groupings. The average difference for these three groupings and five diseases was 0.0427 (SE 3.39 *×* 10^*−*3^), whereas for all other diseases it was 0.0242 (SE 3.15 *×* 10^*−*3^) (Table S21-S23). Among the other 17 significant AUC differences within these three groupings all but three maintained the same directional bias. Two of the three contradictory differences involved the disease of depression (Table S24-S28). Among the five other characteristics analyzed, which included years of education and various census^32^ measurements, only four significant differences were found without any patterns emerging(Table S23-S27).

The characteristic expected to have the greatest effect upon a PRS is the underlying ancestral population. The individuals under analysis were therefore expanded to evaluate differences between the British and each of the non-British populations, specifically, European, African and Asian. The PRS distributions were directly compared as the small quantity of disease cases in each non-British group would lead to large uncertainties in any predictive performance metric. 94.4% of British to any non-British score distribution comparisons were significantly different across all diseases. Ovarian cancer maintained the most similar distributions with a maximal 13.6% difference between the mean of the British and any of the non-British score distribution means. Whereas, NAFLD generated the most varied distributions with a minimal 74.6% difference between the mean of the British and any of the non-British score distribution means (Table S29-S30). The score distributions most similar to the British was Asian, with a mean percent difference between distribution means of 15.6% (SE 3.70%), followed by European with a mean of 17.3% (SE 3.93%) and lastly African with a mean of 25.5% (SE 7.96%). These mean statistics may mask drastic shifts of the high score strata, which could be highly detrimental to non-British populations assessed by PRSs in the clinic.

drastic, detrimental shifts in risk of certain individuals from the British distributions.

Lastly, we analyzed the disease labels used throughout these analyses by considering alternative labeling methods. For example, disease labels could be derived solely from self-reports or the electronic health records. These comparisons reveal the sensitivity in the analyses of 7 diseases, which experience an AUC difference between the primary and an alternative labeling method greater than 0.01. The disease with the greatest AUC difference is Celiac Disease, which experiences an increase in AUC from 0.667 to 0.911 when removing self-reports and only considering individuals with positive ICD codes (Table S31). This ICD-only labeling method generated all of the instances in which AUCs differed greater than 0.01, indicating the uniformity of all other labeling methods. The totality of individual characteristic comparisons show that PRSs may be partially biased. However, since the bias is often small and even sometimes beneficial, the PRSs are likely still valuable to a clinical model.

### Dissection of the Optimal Polygenic Risk Score Generation

Realistic clinical implementation of PRSs may require calibration of variant weights produced in projects such as this, thereby necessitating a strong understanding of optimal PRS generative processes. When generating a PRS the first critical step is the selection of a method that adjusts a GWAS’s variant weights, such that the ensuing PRS has the greatest possible predictive accuracy. We surveyed all 15 methods available that do not require phenotypic information, and determined that prsCS generated the highest average AUC (0.674, SE 0.0143). The lassosum, LDpred2 and WC-2D methods generated the next three highest mean AUC values (Figure 6A, Table S32). While WC-2D is the least accurate in this group it was relatively easy to use and its decline in performance from prsCS is marginal (decrease mean AUC = 0.010). Alternatively ranking the methods by odds ratio, concordance or cumulative hazard generated the same top three methods with WC-2D only slipping to fifth rank for cumulative hazard and sixth rank for odds ratio (Figure 6B, Table S33-S35). Therefore, when maximal accuracy is required or the practitioner is relatively skilled either the prsCS, lassosum or LDpred2 methods can be relied on, otherwise the WC-2D method is an ideal alternative for creating a fast and reasonably accurate score.

Fine-scale method recommendations that additionally consider attributes of the originating GWAS, such as a relatively large sample size, were also computed. Specifically, we weighted the AUCs generated from PRS-included models by a GWAS attribute (Table M12). The methods corresponding to the top ten weighted AUCs were enumerated. Across all attributes analyzed lassosum was in the most top ten counts, on average 4.38 times (SE 0.155), followed by the two previously reported well-performing methods, prsCS (mean 2.81, SE 0.188) and LDpred2 (mean 1.19, SE 0.164) (Figure 6C, Table S36-S37). The consistent method rankings indicate that GWAS attributes need not be computed nor considered in PRS generation.

While nearly all efforts to develop clinical PRSs will aim to maximize predictive performance, in specific circumstances creating a score with a biological meaning or containing certain characteristics may be useful. For example, some clinicians may considers PRSs with fewer variants to be less error-prone or PRSs with variants that all reside in the same functional annotation to be more interpretable. To this end, we first investigated polygenicity. The number of variants that produced the best PRS for each disease ranged from 21 to 6,129,416 (mean = 548,084, SD = 1,298,793) (Table S38). Next, the variants that produced each PRS were ordered by their absolute effect, and then split into four groups such that the sum of effects of each group was equal to the others (Figure 5A). The size of the group with the smallest effects compared to the group with the largest effects ranged from a factor of 3 to 1,224,967 times larger (Table S39). This wide range of polygenicity indicates that it would be generally difficult to maximize a score’s performance while minimizing the number of component variants.

**Figure 5.**
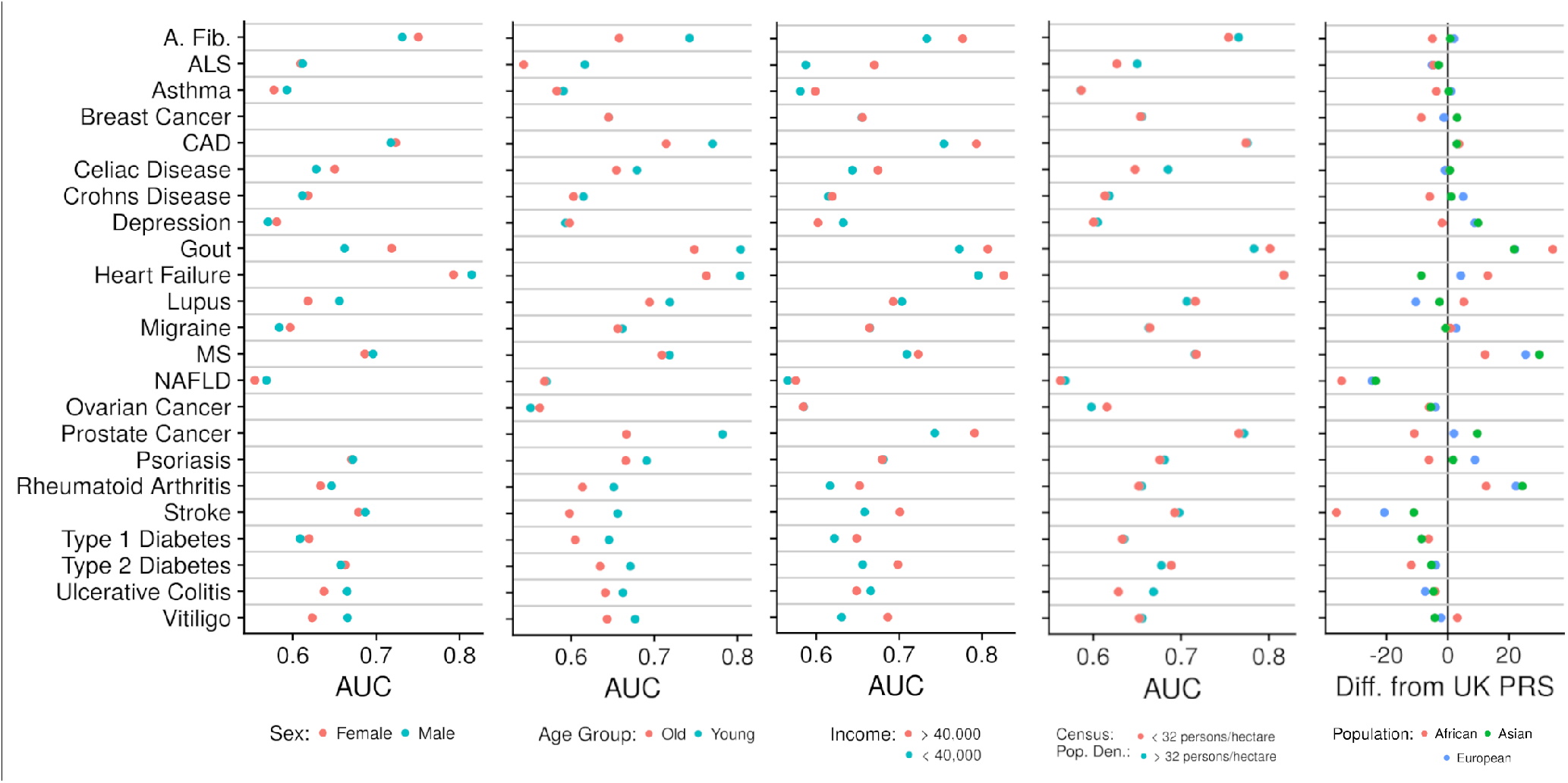
Predictive performance and score distribution statistics stratified by individual characteristics. From left to right the first four panels illustrate AUCs generated when the UK Biobank was split according to individual characteristics described in the respective key, followed by model fitting and assessment. The models include the covariates of age, sex, the top ten genetic principal components and PRS. The young and old splits were determined by calculating median age amongst the cases for each disease. The rightmost panel depicts the mean of the PRS distributions for non-British population subtracted from the British PRS distribution mean

Continuing to investigate the variants composing PRSs, we developed a statistic titled functional annotation weight. Specifically, this weight was computed as the mean of the variants’ absolute effects in each of 44 functional categories^33^, normalized so the sum of values for each disease equaled one. Ancient human promoters had the greatest mean functional annotation weight across all diseases (mean = 0.0456), whereas repressed regions generated the smallest functional annotation weight (mean = 0.0049). Logical clustering of the functional annotation weight emerged, including NAFLD weighting liver cell types higher than any other disease, and autoimmune disease (ulcerative colitis, crohn’s disease, celiac disease and rheumatoid arthritis) all weighting enhancers-1 higher than any other disease (Figure 6D, Table S40-S46). Similar deleterious weights were created from SIFT, PROVEAN and other scores^34,35^, although a meaningful trend did not emerge (Table S47-S48). The clear patterns that emerged from these analyses show that it is likely possible to limit a PRS’s component variants to a strict functional class and still maintain high predictive accuracy.

**Figure 6.**
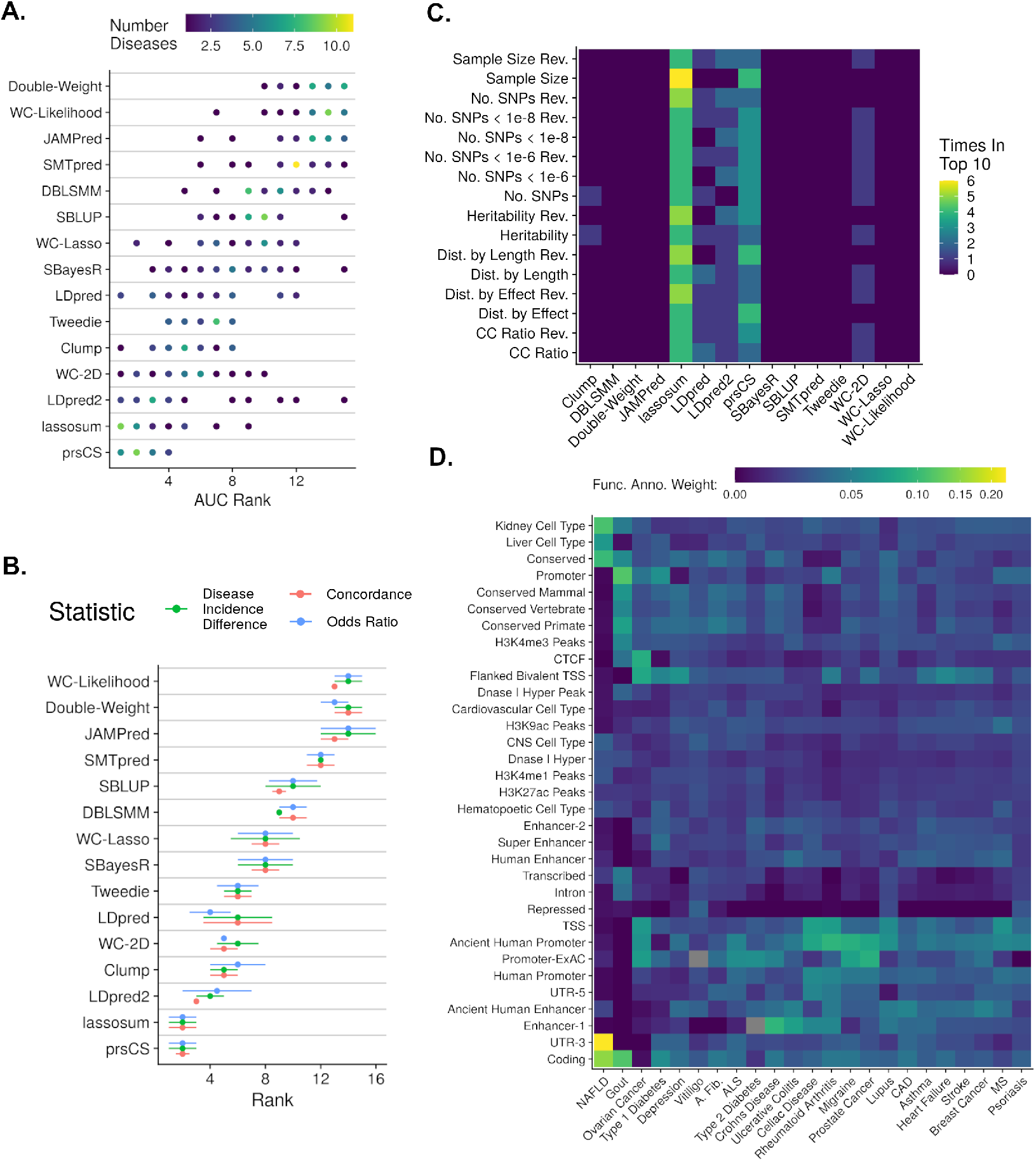
Evidence used to guide PRS construction. A: For each disease, all scores were ranked according to their AUC values determined in the training set of data. The unique ranks each method achieved is shown, shaded according to the number of diseases corresponding to that rank. B: Similarly, the the best score for each method-disease combination was given a rank across the statistics of concordance, odds ratio and disease incidence difference. The mean and standard error of these ranks across all diseases are shown. C: Products between the AUCs corresponding to each method-disease combinations and a GWAS summary statistic attribute, both normalized to a range from 0 to 1, were calculated. The methods in the top 10 product values were counted and plotted. The reversed label denotes when an attribute was sorted such that smaller values rather than larger values were preferred. D: For each disease, the variants of the PRS for each disease were identified as belonging to any one of 44 functional annotation groups. The mean of the absolute effects of the variants in each group were calculated, and normalized such that the sum of resulting functional annotation weights for each disease summed to one.

Finally, to aid in the creation and interpretation of novel PRSs, we provide deeply detailed documentation from this investigation at https://kulmsc.github.io/pgs_book/index.html. From this guide and the previous methodological results, a clinician is given a toolbox that can transform any PRS from acting as an insignificant predictor to a valuable clinical measure.

## Discussion

This comprehensive investigation of PRSs resulted in impressive predictive performances accompanied by definitive caveats that may limit generalizability. Yet, the overwhelming potential benefit and lack of apparent harm associated with including PRSs to current approaches demonstrate that they should be considered for clinical use.

Assessment of each disease’s best PRS proves that most scores can significantly improve basic clinical models. In the analysis of breast cancer, the addition of the best performing PRS to a model of age and the top ten genetic principal components increases the AUC by 0.088. In practical terms, the addition of the PRS reclassifies 74% of individuals in a risk group defined by the top 10% of risk. The incidence of cancer in this risk group would similarly change from 0.82% to 1.12%. We observed similarly impressive predictive performances for heart failure and crohn’s disease, amongst others. Although, a few diseases did see little benefit from an added PRS, such as stroke, indicating the difficulty in making PRS-focused assumptions and the importance of research in this field.

A prevalent misunderstanding of PRSs compares their use to diagnostic tests^36^, such as a biopsy for cancer, leading to the conclusion that PRSs are ineffective. A more useful comparison is to current screening recommendations, for example women beginning mammogram screening at age 50^37^. The results show that combining age and a PRS into a model, then using calibrated risk cut-offs to form recommendations would likely reveal a greater share of individuals with the disease as compared to an age-only approach; thereby exhibiting how PRSs are effective.

Beyond prediction statistics, PRSs are uniformly shown to improve clinical decisions that may be directly translated to more efficacious interventions. A decision curve analysis for all diseases revealed that under at least one risk threshold the inclusion of PRSs in a risk model leads to more “profitable” predictions^28^. Once an individual is predicted to be of high risk, analysis of lifestyle modifications and medications indicate that targeted care can be provided that will aid the individual far more than if they were in another risk group. For example, individuals with high compared to low PRS will experience an 550% greater absolute risk reduction in CAD when quitting smoking. By targeting this modification to the high PRS individuals as compared to an equally sized random sample of individuals, approximately 1,040 fewer people out of 100,00 are associated with developing CAD. A similar methodology applied to medications and supplements indicated that ibuprofen used in individuals with high PRS can reduce the risk of atrial fibrillation, type 2 diabetes and heart failure better than for individuals with low PRS. A few interesting results involving cheese, alcohol and evening primrose oil underscore the need for additional, targeted research in this area. These results on their own merits still ably prove that PRSs may extend from simple predictors of risk to instruments that provide beneficial clinical action recommendations.

Well reported confounding factors within traditional genetic analyses^38,39^ have led to concerns that PRSs are heavily and improperly influenced by individual characteristics. We show that assessments of PRSs within characteristic groupings do partially discredit some scores. However, many of grouping comparisons either reveal areas in which PRSs have greater clinical impact, such as younger individuals, or find no differences whatsoever, such as all census comparisons. Furthermore, after the addition of clinically relevant covariates to the base model, the further inclusion of the PRS always increases the predictive accuracy as measured by both AUC and net benefit. In the pursuit of an optimal PRS within a specific clinical population these small effects of individual characteristic must nevertheless be considered, and properly adjusted for.

To aid in the calibration of PRSs to clinic-specific populations, we completed a full-analysis of generative methods. The prsCS method performed best, although the simpler clumping method still generated impressive scores irrespective of the GWAS’s starting characteristics. By following the detailed documentation provided, interested clinicians should be able to easily produce PRSs for their own practice.

Several methodological assumptions may limit the generalizability of the results presented. First, all predictive assessments were made solely on a white-British population. Several studies have clearly shown the limited portability of such PRSs, and current disparities in healthcare would be exacerbated by their current use^40,41^. Even within a single ethnic group, fine-scale population structure^42,43^ and sample bias^44,45^ have been shown to exist, yet were not mediated here. Lastly, use of only a single biobank, which may contain a host of unique biases, adds further possible confounding to the results displayed. Despite these definite limitations, the care taken to use only external summary statistics, reduce overfitting, and check PRS validity renders these results as authentic as possible.

The totality of our results suggests that PRSs are fit for clinical use. Opposition to this stance does exist, with antagonists pointing out costly implementation and remaining uncertainty in real-world environments^46,47^. However, genome sequencing is a relatively affordable procedure that provides lifetime insight with respect to all ailments; and rather than focusing on the risk of applying PRSs one should consider the harm done in not doing so. The detrimental inaction of leaving an individual with an extremely high PRS in a low risk class where they will not receive beneficial interventions has been understood by several forward-thinking research teams who are building systems to score their patients^48,49^. Restricted acceptance by the entire medical community will likely exacerbate economic healthcare disparities and provoke unethical applications^50^. The future work required to fully realize PRS benefits includes calibration to specific populations and inclusion of all possibly relevant covariates within the risk model. This investigation serves as a guide to these final steps and moreover aims to motivates the entire process of utilizing polygenic risk scores within the clinic.

## Data Availability

All resources utilized are thoroughly described within https://kulmsc.github.io/pgs_book/index.html. In short, data originated from: UK Biobank (Application #47137) (https://www.ukbiobank.ac.uk/), 1000 Genomes Project (https://www.cog-genomics.org/plink/2.0/resources#1kg_phase3), GWAS Catalog (https://www.ebi.ac.uk/gwas/downloads/summary-statistics), PGS Catalog (https://www.pgscatalog.org/browse/all/), Functional Annotations (https://alkesgroup.broadinstitute.org/LDSCORE/), Deleterious Scores (https://pcingola.github.io/SnpEff/ss_dbnsfp/). All intermediate data that does not contain individual-level UK Biobank records is available at: https://wcm.box.com/s/oe3oayaoi3mxszf38c0bqpaa8r8ftmif. All custom written scripts are available at https://github.com/kulmsc/pgs_scripts. Additional generative method software are described in Supplementary II. The programming languages R, version 3.6, and Python, version 3.6.5, were utilized.

https://github.com/kulmsc

## Code and Data Availability

All resources utilized are thoroughly described within https://kulmsc.github.io/pgs_book/index.html. In short, data originated from: UK Biobank (Application 47137) (https://www.ukbiobank.ac.uk/), 1000 Genomes Project (https://www.cog-genomics.org/plink/2.0/resources#1kg_phase3), GWAS Catalog (https://www.ebi.ac.uk/gwas/downloads/summary-statistics), PGS Catalog (https://www.pgscatalog.org/browse/all/), Functional Annotations (https://alkesgroup.broadinstitute.org/LDSCORE/), Deleterious Scores (https://pcingola.github.io/SnpEff/ss_dbnsfp/). All intermediate data that does not contain individual-level UK Biobank records is available at: https://wcm.box.com/s/oe3oayaoi3mxszf38c0bqpaa8r8ftmif. All custom written scripts are available at https://github.com/kulmsc/pgs_scripts. Additional generative method software are described in Supplementary II. The programming languages R, version 3.6, and Python, version 3.6.5, were utilized.

## Author Contributions

S.K., J.M. and O.E. conceived and designed the study. S.K., J.M. and O.E. all aided in data acquisition. S.K. principally analyzed and interpreted the data with aid from A.M. S.K. drafted the manuscript. A.M., J.M. and O.E. provided critical feedback on the manuscript.

## References

1. Torkamani, A., Wineinger, N. E. & Topol, E. J. The personal and clinical utility of polygenic risk scores. Nat. Rev. Genet. 19, 581–590, DOI: 10.1038/s41576-018-0018-x (2018).

2. ho Lee, Y., Bang, H. & Kim, D. J. How to establish clinical prediction models. Endocrinol. Metab. 31, 38, DOI: 10.3803/enm.2016.31.1.38 (2016).

3. Mars, N. J. et al. Polygenic and clinical risk scores and their impact on age at onset of cardiometabolic diseases and common cancers. DOI: 10.1101/727057 (2019).

4. Vreemann, S. et al. The added value of mammography in different age-groups of women with and without BRCA mutation screened with breast MRI. Breast Cancer Res. 20,DOI: 10.1186/s13058-018-1019-6 (2018).

5. Golan, D., Lander, E. S. & Rosset, S. Measuring missing heritability: Inferring the contribution of common variants. Proc. Natl. Acad. Sci. 111, E5272–E5281, DOI: 10.1073/pnas.1419064111 (2014).

6. Khera, A. V. et al. Genome-wide polygenic scores for common diseases identify individuals with risk equivalent to monogenic mutations. Nat. Genet. 50, 1219–1224, DOI: 10.1038/s41588-018-0183-z (2018).

7. Natarajan, P. et al. Polygenic risk score identifies subgroup with higher burden of atherosclerosis and greater relative benefit from statin therapy in the primary prevention setting. Circulation 135, 2091–2101, DOI: 10.1161/circulationaha.116.024436 (2017).

8. Mosley, J. D. et al. Predictive accuracy of a polygenic risk score compared with a clinical risk score for incident coronary heart disease. JAMA 323, 627, DOI: 10.1001/jama.2019.21782 (2020).

9. Ge, T., Chen, C.-Y., Ni, Y., Feng, Y.-C.A. & Smoller, J. W. Polygenic prediction via Bayesian regression and continuous shrinkage priors. Nat. Commun. 10, 1–10, DOI: 10.1038/s41467-019-09718-5 (2019).

10. Lloyd-Jones, L. R. et al. Improved polygenic prediction by Bayesian multiple regression on summary statistics. Nat. Commun. 10, 1–11, DOI: 10.1038/s41467-019-12653-0 (2019).

11. Mak, T. S. H., Porsch, R. M., Choi, S. W., Zhou, X. & Sham, P. C. Polygenic scores via penalized regression on summary statistics. Genet. Epidemiol. 41, 469–480, DOI: 10.1002/gepi.22050 (2017).

12. Bycroft, C. et al. The UK biobank resource with deep phenotyping and genomic data. Nature 562, 203–209, DOI: 10.1038/s41586-018-0579-z (2018).

13. Chang, C. C. et al. Second-generation PLINK: rising to the challenge of larger and richer datasets. GigaScience 4, 7, DOI: 10.1186/s13742-015-0047-8 (2015).

14. Läll, K., Mägi, R., Morris, A., Metspalu, A. & Fischer, K. Personalized risk prediction for type 2 diabetes: the potential of genetic risk scores. Genet. Medicine 19, 322–329, DOI: 10.1038/gim.2016.103 (2016).

15. Shi, J. et al. Winner’s Curse Correction and Variable Thresholding Improve Performance of Polygenic Risk Modeling Based on Genome-Wide Association Study Summary-Level Data. PLOS Genet. 12, e1006493, DOI: 10.1371/journal.pgen.1006493 (2016).

16. So, H.-C. & Sham, P. C. Improving polygenic risk prediction from summary statistics by an empirical Bayes approach. Sci. Reports 7, 1–11, DOI: 10.1038/srep41262 (2017).

17. Vilhjálmsson, B. J. et al. Modeling Linkage Disequilibrium Increases Accuracy of Polygenic Risk Scores. Am. J. Hum. Genet. 97, 576–592, DOI: 10.1016/j.ajhg.2015.09.001 (2015).

18. Privé, F., Arbel, J. & Vilhjálmsson, B. J. LDpred2: better, faster, stronger. Bioinformatics DOI: 10.1093/bioinformatics/btaa1029 (2020).

19. Robinson, M. R. et al. Genetic evidence of assortative mating in humans. Nat. Hum. Behav. 1, DOI: 10.1038/s41562-016-0016 (2017).

20. Yang, S. & Zhou, X. Accurate and scalable construction of polygenic scores in large biobank data sets. The Am. J. Hum. Genet. 106, 679–693, DOI: 10.1016/j.ajhg.2020.03.013 (2020).

21. Newcombe, P. J., Nelson, C. P., Samani, N. J. & Dudbridge, F. A flexible and parallelizable approach to genome-wide polygenic risk scores. Genet. Epidemiol. 43, 730–741, DOI: 10.1002/gepi.22245 (2019).

22. Maier, R. M. et al. Improving genetic prediction by leveraging genetic correlations among human diseases and traits. Nat. Commun. 9, 1–17, DOI: 10.1038/s41467-017-02769-6 (2018).

23. Dunlay, S. M., Weston, S. A., Jacobsen, S. J. & Roger, V. L. Risk factors for heart failure: A population-based case-control study. The Am. J. Medicine 122, 1023–1028, DOI: 10.1016/j.amjmed.2009.04.022 (2009).

24. Steven D. Stellman, L. W. Y. C. M. L. C. M. V. D. S. H. J. E. M. A. I. N. E. L. W. H. O. K. T., Toshiro Takezaki & Aoki, K. Smoking and lung cancer risk in american and japanese men: An international case-control study. Cancer Epidemiol. Biomarkers Prev. 10, DOI: NA (2001).

25. Pencina, M. J., Agostino, R. B. D., Agostino, R. B. D. & Vasan, R. S. Evaluating the added predictive ability of a new marker: From area under the ROC curve to reclassification and beyond. Stat. Medicine 27, 157–172, DOI: 10.1002/sim.2929 (2007).

26. Pepe, M. S., Fan, J., Feng, Z., Gerds, T. & Hilden, J. The net reclassification index (NRI): A misleading measure of prediction improvement even with independent test data sets. Stat. Biosci. 7, 282–295, DOI: 10.1007/s12561-014-9118-0 (2014).

27. Lambert, S. A., Abraham, G. & Inouye, M. Towards clinical utility of polygenic risk scores. Hum. Mol. Genet. 28, R133–R142, DOI: 10.1093/hmg/ddz187 (2019).

28. Vickers, A. J., van Calster, B. & Steyerberg, E. W. A simple, step-by-step guide to interpreting decision curve analysis. Diagn. Progn. Res. 3, DOI: 10.1186/s41512-019-0064-7 (2019).

29. Ye, Y. et al. Interactions between enhanced polygenic risk scores and lifestyle for cardiovascular disease, diabetes, and lipid levels. Circ. Genomic Precis. Medicine 14, DOI: 10.1161/circgen.120.003128 (2021).

30. Kogelman, L. J. et al. Migraine polygenic risk score associates with efficacy of migraine-specific drugs. Neurol. Genet. 5, e364, DOI: 10.1212/nxg.0000000000000364 (2019).

31. Hettige, N. C., Cole, C. B., Khalid, S. & Luca, V. D. Polygenic risk score prediction of antipsychotic dosage in schizophrenia. Schizophr. Res. 170, 265–270, DOI: 10.1016/j.schres.2015.12.015 (2016).

32. for National Statistics, O. 2011.

33. Finucane, H. K. et al. Partitioning heritability by functional annotation using genome-wide association summary statistics. Nat. Genet. 47, 1228–1235, DOI: 10.1038/ng.3404 (2015).

34. Liu, X., Jian, X. & Boerwinkle, E. dbNSFP: A lightweight database of human nonsynonymous SNPs and their functional predictions. Hum. Mutat. 32, 894–899, DOI: 10.1002/humu.21517 (2011).

35. Liu, X., Li, C., Mou, C., Dong, Y. & Tu, Y. dbNSFP v4: a comprehensive database of transcript-specific functional predictions and annotations for human nonsynonymous and splice-site SNVs. Genome Medicine 12, DOI: 10.1186/s13073-020-00803-9 (2020).

36. Wald, N. J. & Old, R. The illusion of polygenic disease risk prediction. Genet. Medicine 21, 1705–1707, DOI: 10.1038/s41436-018-0418-5 (2019).

37. and, A. L. S. Screening for breast cancer: U.s. preventive services task force recommendation statement. Annals Intern. Medicine 164, 279, DOI: 10.7326/m15-2886 (2016).

38. Mathieson, I. & McVean, G. Differential confounding of rare and common variants in spatially structured populations. Nat. Genet. 44, 243–246, DOI: 10.1038/ng.1074 (2012).

39. Vilhjálmsson, B. J. & Nordborg, M. The nature of confounding in genome-wide association studies. Nat. Rev. Genet. 14, 1–2, DOI: 10.1038/nrg3382 (2012).

40. Martin, A. R. et al. Clinical use of current polygenic risk scores may exacerbate health disparities. Nat. Genet. 51, 584–591, DOI: 10.1038/s41588-019-0379-x (2019).

41. Duncan, L. et al. Analysis of polygenic risk score usage and performance in diverse human populations. Nat. Commun. 10, DOI: 10.1038/s41467-019-11112-0 (2019).

42. Sakaue, S. et al. Dimensionality reduction reveals fine-scale structure in the japanese population with consequences for polygenic risk prediction. Nat. Commun. 11, DOI: 10.1038/s41467-020-15194-z (2020).

43. Mostafavi, H. et al. Variable prediction accuracy of polygenic scores within an ancestry group. eLife 9, DOI: 10.7554/elife.48376 (2020).

44. Tyrrell, J. et al. Genetic predictors of participation in optional components of UK biobank. DOI: 10.1101/2020.02.10.941328 (2020).

45. Pirastu, N. et al. Genetic analyses identify widespread sex-differential participation bias. DOI: 10.1101/2020.03.22.001453 (2020).

46. Vega, F.M.D.L. & Bustamante, C. D. Polygenic risk scores: a biased prediction? Genome Medicine 10, DOI: 10.1186/s13073-018-0610-x (2018).

47. Wray, N. R. et al. Pitfalls of predicting complex traits from SNPs. Nat. Rev. Genet. 14, 507–515, DOI: 10.1038/nrg3457 (2013).

48. Ashenhurst, J. R. et al. A generalized method for the creation and evaluation of polygenic scores. https://medical.23andme. com/wp-content/uploads/2020/06/23_21-PRSMethodology_May2020.pdf (2020). Accessed: 2021-01-28.

49. Plc, G. Polygenic risk scores. https://www.genomicsplc.com/wp-content/uploads/2020/11/Genomics-plc-PRS-details_White-Paper-April-2019.pdf (2019). Accessed: 2021-01-28.

50. Ganna, A. et al. Large-scale GWAS reveals insights into the genetic architecture of same-sex sexual behavior. Science 365, eaat7693, DOI: 10.1126/science.aat7693 (2019).

