## Supplemental I: Tables and Figures for "A systematic framework for assessing the clinical impact of polygenic risk scores"

| Disease | P-Value |
| --- | --- |
| Lupus | 7.98e-05 |
| A. Fib. | 4.56e-101 |
| Asthma | 4.5e-113 |
| Celiac Disease | 1.2e-16 |
| Migraine | 1.79e-41 |
| MS | 3.28e-27 |
| Vitiligo | 3.83e-08 |
| Gout | 1.58e-45 |
| Crohns Disease | 1.86e-23 |
| Ulcerative Colitis | 1.41e-23 |
| Type 2 Diabetes | 8.57e-57 |
| Stroke | 3e-06 |
| Breast Cancer | 3.79e-126 |
| NAFLD | 2.73e-05 |
| CAD | 2.35e-68 |
| Rheumatoid Arthritis | 6.39e-16 |
| Type 1 Diabetes | 2.62e-11 |
| Ovarian Cancer | 0.382 |
| ALS | 0.131 |
| Prostate Cancer | 4.86e-114 |
| Heart Failure | 6.25e-91 |
| Psoriasis | 8.65e-94 |
| Depression | 1.84e-17 |

**Supplementary Table S1.** The P-values generated from a DeLong test between ROCs generated from models containing age, sex and the top ten genetic principal components and a model containing age, sex, the top ten genetic principal components and the respective polygenic risk score.

| | AUC | AUC-Imp. | OR | OR-Imp. | Conc. | Conc.-Imp. | Haz. | Haz.-Imp. |
|---|---|---|---|---|---|---|---|---|
| AUC | 1 | 0.102 | 0.963 | 0.799 | 0.943 | 0.13 | 0.92 | -0.884 |
| AUC-Imp. | 0.102 | 1 | 0.0208 | 0.465 | 0.307 | 0.975 | 0.146 | 0.169 |
| OR | 0.963 | 0.0208 | 1 | 0.808 | 0.86 | 0.0623 | 0.908 | -0.886 |
| OR-Imp. | 0.799 | 0.465 | 0.808 | 1 | 0.845 | 0.486 | 0.765 | -0.617 |
| Conc. | 0.943 | 0.307 | 0.86 | 0.845 | 1 | 0.32 | 0.893 | -0.806 |
| Conc.-Imp. | 0.13 | 0.975 | 0.0623 | 0.486 | 0.32 | 1 | 0.151 | 0.162 |
| Haz. | 0.92 | 0.146 | 0.908 | 0.765 | 0.893 | 0.151 | 1 | -0.885 |
| Haz.-Imp. | -0.884 | 0.169 | -0.886 | -0.617 | -0.806 | 0.162 | -0.885 | 1 |

**Supplementary Table S2.** The spearman correlations between the vectors of 23 statistics, one value for each disease. Imp. refers to the improvement, or difference in models that include or do not include the polygenic risk score.

| Disease | Orig. Base | Score Added |
|---|---|---|
| Lupus | 1614 | 6492 |
| A. Fib. | 4371 | 3731 |
| Asthma | 1171 | 6922 |
| Celiac Disease | 1374 | 6732 |
| Migraine | 2926 | 5176 |
| MS | 1447 | 6657 |
| Vitiligo | 2441 | 5665 |
| Gout | 3229 | 4876 |
| Crohns Disease | 1314 | 6788 |
| Ulcerative Colitis | 788 | 7315 |
| Type 2 Diabetes | 3894 | 4211 |
| Stroke | 5819 | 2283 |
| Breast Cancer | 713 | 3696 |
| NAFLD | 1707 | 6399 |
| CAD | 3865 | 4233 |
| Rheumatoid Arthritis | 2700 | 5404 |
| Type 1 Diabetes | 2726 | 5379 |
| Ovarian Cancer | 1786 | 2637 |
| ALS | 4199 | 3907 |
| Prostate Cancer | 987 | 2696 |
| Heart Failure | 1988 | 6117 |
| Psoriasis | 884 | 7220 |
| Depression | 3518 | 4588 |

**Supplementary Table S3.** Count of individuals moving into and out of the top 5% risk group with the incusion of the polygenic risk score in the underlying risk model.

| **Disease** | **NRI** | **NRI - 95% CI Low** | **NRI - 95% CI High** |
|---|---|---|---|
| Lupus | 0.1128 | 0.0674 | 0.1582 |
| A. Fib. | 0.0458 | 0.0415 | 0.0502 |
| Asthma | 0.0591 | 0.0542 | 0.0641 |
| Celiac Disease | 0.1393 | 0.1093 | 0.1693 |
| Migraine | 0.0552 | 0.0482 | 0.0622 |
| MS | 0.1706 | 0.1412 | 0.2001 |
| Vitiligo | 0.0113 | -0.0111 | 0.0336 |
| Gout | 0.1172 | 0.1061 | 0.1284 |
| Crohns Disease | 0.1019 | 0.0822 | 0.1216 |
| Ulcerative Colitis | 0.1224 | 0.0961 | 0.1487 |
| Type 2 Diabetes | 0.0433 | 0.0386 | 0.0479 |
| Stroke | 0.011 | 0.0077 | 0.0143 |
| Breast Cancer | 0.1775 | 0.1661 | 0.189 |
| NAFLD | 0.0638 | 0.0457 | 0.0818 |
| CAD | 0.0811 | 0.074 | 0.0881 |
| Rheumatoid Arthritis | 0.0668 | 0.0566 | 0.077 |
| Type 1 Diabetes | 0.049 | 0.0345 | 0.0635 |
| Ovarian Cancer | 0.016 | 0.0034 | 0.0287 |
| ALS | 0.0026 | -0.0075 | 0.0127 |
| Prostate Cancer | 0.1142 | 0.1038 | 0.1247 |
| Heart Failure | 0.3741 | 0.3593 | 0.3889 |
| Psoriasis | 0.2272 | 0.2096 | 0.2449 |
| Depression | 0.0176 | 0.0139 | 0.0214 |

**Supplementary Table S4.** The Net Reclassification Improvement (NRI) values for each disease using the 95th percentile risk as the cutoff within the categorical computation.

| Disease | IDI | IDI 95% CI Low | IDI 95% CI High |
|---|---|---|---|
| Lupus | 8e-04 | 4e-04 | 0.0011 |
| A. Fib. | 0.0167 | 0.0157 | 0.0177 |
| Asthma | 0.0106 | 0.0101 | 0.0112 |
| Celiac Disease | 0.0018 | 0.0014 | 0.0021 |
| Migraine | 0.0038 | 0.0035 | 0.0042 |
| MS | 0.0039 | 0.0033 | 0.0044 |
| Vitiligo | 2e-04 | 1e-04 | 3e-04 |
| Gout | 0.0099 | 0.009 | 0.0109 |
| Crohns Disease | 0.0018 | 0.0015 | 0.002 |
| Ulcerative Colitis | 0.0021 | 0.0018 | 0.0024 |
| Type 2 Diabetes | 0.0081 | 0.0075 | 0.0086 |
| Stroke | 0.0013 | 0.001 | 0.0015 |
| Breast Cancer | 0.0247 | 0.0233 | 0.026 |
| NAFLD | 5e-04 | 4e-04 | 7e-04 |
| CAD | 0.0144 | 0.0134 | 0.0154 |
| Rheumatoid Arthritis | 0.0033 | 0.0028 | 0.0037 |
| Type 1 Diabetes | 0.0011 | 9e-04 | 0.0013 |
| Ovarian Cancer | 3e-04 | 1e-04 | 5e-04 |
| ALS | 1e-04 | 0 | 1e-04 |
| Prostate Cancer | 0.0337 | 0.0315 | 0.0358 |
| Heart Failure | 0.1365 | 0.1302 | 0.1428 |
| Psoriasis | 0.0104 | 0.0095 | 0.0112 |
| Depression | 0.0023 | 0.0021 | 0.0026 |

**Supplementary Table S5.** The integrated discrimination index (IDI) values computed for each disease. The definition of IDI is provided within *Evaluating the added predictive ability of a new marker: From area under the ROC curve to reclassification and beyond* by Pescina et al.

| Disease | Orig. Base | Score Added |
|---|---|---|
| Lupus | 0.00168 | 0.00168 |
| A. Fib. | 0.0597 | 0.0587 |
| Asthma | 0.122 | 0.121 |
| Celiac Disease | 0.00439 | 0.00438 |
| Migraine | 0.0381 | 0.038 |
| MS | 0.00451 | 0.00449 |
| Vitiligo | 0.000617 | 0.000617 |
| Gout | 0.0247 | 0.0245 |
| Crohns Disease | 0.0106 | 0.0106 |
| Ulcerative Colitis | 0.00587 | 0.00586 |
| Type 2 Diabetes | 0.0652 | 0.0647 |
| Stroke | 0.0352 | 0.0351 |
| Breast Cancer | 0.0664 | 0.0647 |
| NAFLD | 0.00992 | 0.00992 |
| CAD | 0.0465 | 0.0458 |
| Rheumatoid Arthritis | 0.0221 | 0.022 |
| Type 1 Diabetes | 0.00888 | 0.00888 |
| Ovarian Cancer | 0.00784 | 0.00784 |
| ALS | 0.000917 | 0.000917 |
| Prostate Cancer | 0.0525 | 0.0506 |
| Heart Failure | 0.0282 | 0.0234 |
| Psoriasis | 0.0182 | 0.018 |
| Depression | 0.0591 | 0.059 |

**Supplementary Table S6.** Brier values computed from models that either contain the base covariates, or a model that also includes the best respective polygenic risk score.

| **Disease** | **Base** | **Score** |
|---|---|---|
| Lupus | 0.00308 | 0.0053 |
| A. Fib. | 0.196 | 0.239 |
| Asthma | 0.158 | 0.216 |
| Celiac Disease | 0.0079 | 0.0132 |
| Migraine | 0.0664 | 0.0942 |
| MS | 0.00728 | 0.0196 |
| Vitiligo | 0.00111 | 0.00185 |
| Gout | 0.0664 | 0.0992 |
| Crohns Disease | 0.0143 | 0.0228 |
| Ulcerative Colitis | 0.00494 | 0.0153 |
| Type 2 Diabetes | 0.15 | 0.185 |
| Stroke | 0.1 | 0.104 |
| Breast Cancer | 0.103 | 0.198 |
| NAFLD | 0.0116 | 0.0183 |
| CAD | 0.144 | 0.181 |
| Rheumatoid Arthritis | 0.0418 | 0.055 |
| Type 1 Diabetes | 0.0171 | 0.0212 |
| Ovarian Cancer | 0.0122 | 0.0154 |
| ALS | 0.00296 | 0.00259 |
| Prostate Cancer | 0.118 | 0.221 |
| Heart Failure | 0.0886 | 0.233 |
| Psoriasis | 0.0231 | 0.0631 |
| Depression | 0.0925 | 0.107 |

**Supplementary Table S7.** The true positive rates determined for the top 5% risk group.

| Disease | Low Risk | Intermediate Risk | High Risk |
|---|---|---|---|
| Lupus | 0.000241 | 0.00109 | 0.0039 |
| A. Fib. | 0.0101 | 0.0429 | 0.142 |
| Asthma | 0.088 | 0.137 | 0.214 |
| Celiac Disease | 0.00125 | 0.00343 | 0.00922 |
| Migraine | 0.014 | 0.0356 | 0.0763 |
| MS | 8e-04 | 0.00287 | 0.0109 |
| Vitiligo | 0.000206 | 0.000483 | 0.0011 |
| Gout | 0.00253 | 0.0149 | 0.0657 |
| Crohns Disease | 0.00502 | 0.00897 | 0.0161 |
| Ulcerative Colitis | 0.00207 | 0.00458 | 0.0103 |
| Type 2 Diabetes | 0.0245 | 0.0556 | 0.118 |
| Stroke | 0.0094 | 0.026 | 0.056 |
| Breast Cancer | 0.0243 | 0.058 | 0.137 |
| NAFLD | 0.00663 | 0.00826 | 0.012 |
| CAD | 0.00826 | 0.0314 | 0.11 |
| Rheumatoid Arthritis | 0.00804 | 0.018 | 0.0395 |
| Type 1 Diabetes | 0.00342 | 0.00628 | 0.0118 |
| Ovarian Cancer | 0.00331 | 0.00483 | 0.00682 |
| ALS | 8.39e-05 | 0.000189 | 0.000446 |
| Prostate Cancer | 0.00793 | 0.0366 | 0.134 |
| Heart Failure | 0.00198 | 0.0108 | 0.0701 |
| Psoriasis | 0.00808 | 0.0134 | 0.0374 |
| Depression | 0.0328 | 0.0578 | 0.0985 |

**Supplementary Table S8.** The hazards at the final time-point in the study generated from models that include age, sex, top ten genetic principal components and the respective polygenic risk score

| Disease | Low Risk | Intermediate Risk | High Risk |
|---|---|---|---|
| Lupus | -0.000124 | -0.000274 | 0.000908 |
| A. Fib. | -0.00186 | -0.00336 | 0.0143 |
| Asthma | -0.0264 | -0.00389 | 0.0405 |
| Celiac Disease | -0.000981 | -0.000651 | 0.003 |
| Migraine | -0.00211 | -0.00177 | 0.00776 |
| MS | -0.000803 | -0.000849 | 0.00342 |
| Vitiligo | -0.000108 | -4.52e-05 | 0.000253 |
| Gout | -0.000721 | -0.00224 | 0.0083 |
| Crohns Disease | -0.00323 | -0.000566 | 0.00504 |
| Ulcerative Colitis | -0.00231 | -0.000581 | 0.00417 |
| Type 2 Diabetes | -0.00365 | -0.00201 | 0.0104 |
| Stroke | -0.000317 | -0.000459 | 0.00152 |
| Breast Cancer | -0.0173 | -0.00787 | 0.0455 |
| NAFLD | -0.00127 | -0.000413 | 0.00251 |
| CAD | -0.00196 | -0.00221 | 0.00968 |
| Rheumatoid Arthritis | -0.0015 | -0.00107 | 0.00492 |
| Type 1 Diabetes | -0.00131 | -0.000351 | 0.00242 |
| Ovarian Cancer | -0.000168 | -4.5e-05 | 0.000318 |
| ALS | -1.4e-05 | -9.45e-06 | 4.3e-05 |
| Prostate Cancer | -0.00427 | -0.0085 | 0.0334 |
| Heart Failure | -0.00245 | -0.00588 | 0.0243 |
| Psoriasis | -0.00569 | -0.00336 | 0.0166 |
| Depression | -0.00253 | -0.000858 | 0.00542 |

**Supplementary Table S9.** The hazards at the final time-point in the study generated from models that included the polygenic risk score minus the hazards at the same time point but for models without the polygenic risk score.

| Disease | Score - No Extra | Base - No Extra | Score - Extra | Base - Extra |
|---|---|---|---|---|
| Lupus | NA | 0.711 | NA | 0.661 |
| A. Fib. | 0.789 | 0.76 | 0.771 | 0.736 |
| Asthma | 0.586 | 0.586 | 0.531 | 0.531 |
| Celiac Disease | 0.669 | 0.667 | 0.59 | 0.587 |
| Migraine | 0.66 | 0.663 | 0.635 | 0.639 |
| MS | 0.716 | 0.716 | 0.611 | 0.61 |
| Vitiligo | 0.667 | 0.668 | 0.54 | 0.543 |
| Gout | 0.825 | 0.792 | 0.808 | 0.77 |
| Crohns Disease | 0.618 | 0.618 | 0.542 | 0.542 |
| Ulcerative Colitis | 0.646 | 0.646 | 0.497 | 0.5 |
| Type 2 Diabetes | 0.798 | 0.683 | 0.789 | 0.662 |
| Stroke | 0.753 | 0.696 | 0.752 | 0.692 |
| Breast Cancer | 0.679 | 0.68 | 0.59 | 0.592 |
| NAFLD | 0.712 | 0.568 | 0.7 | 0.537 |
| CAD | 0.821 | 0.761 | 0.811 | 0.736 |
| Rheumatoid Arthritis | 0.655 | 0.655 | 0.631 | 0.631 |
| Type 1 Diabetes | NA | 0.638 | NA | 0.599 |
| Ovarian Cancer | 0.601 | 0.604 | 0.599 | 0.599 |
| ALS | 0.656 | 0.654 | 0.644 | 0.641 |
| Prostate Cancer | 0.769 | 0.769 | 0.704 | 0.704 |
| Heart Failure | 0.857 | 0.817 | 0.812 | 0.737 |
| Psoriasis | 0.678 | 0.678 | 0.544 | 0.543 |
| Depression | 0.604 | 0.603 | 0.589 | 0.589 |

**Supplementary Table S10.** The AUCs generated from models that include age, sex, top ten genetic principal components and either a polygenic risk score or extra covariates as specified.

| Disease | AUC | Score |
|---|---|---|
| Lupus | 0.711 | Internal |
| Lupus | 0.698 | Knevel-2020 |
| A. Fib. | 0.76 | Internal |
| A. Fib. | 0.757 | Khera-2018 |
| Asthma | 0.586 | Internal |
| Asthma | 0.545 | Belsky-2013 |
| Gout | 0.792 | Internal |
| Gout | 0.771 | Knevel-2020 |
| Type 2 Diabetes | 0.683 | Internal |
| Type 2 Diabetes | 0.701 | Khera-2018 |
| Type 2 Diabetes | 0.697 | Lall-2016 |
| Type 2 Diabetes | 0.706 | Vassy-2014 |
| Stroke | 0.696 | Internal |
| Stroke | 0.696 | Rutten-2018 |
| Breast Cancer | 0.68 | Internal |
| Breast Cancer | 0.653 | Mavaddat-2018 |
| Breast Cancer | 0.66 | Kuchenbaecker-2018 |
| Breast Cancer | 0.634 | Kuchenbaecker-2018 |
| Breast Cancer | 0.648 | Khera-2018 |
| Breast Cancer | 0.607 | Shieh-2016 |
| CAD | 0.761 | Internal |
| CAD | 0.746 | Tada-2015 |
| CAD | 0.76 | Abraham-2016 |
| CAD | 0.757 | Khera-2018 |
| Rheumatoid Arthritis | 0.655 | Internal |
| Rheumatoid Arthritis | 0.635 | Knevel-2020 |
| Rheumatoid Arthritis | 0.635 | Knevel-2020 |
| Ovarian Cancer | 0.604 | Internal |
| Ovarian Cancer | 0.599 | Graff-2020 |
| Prostate Cancer | 0.769 | Internal |
| Prostate Cancer | 0.746 | Schumacher-2018 |
| Prostate Cancer | 0.709 | Pashayan-2015 |
| Prostate Cancer | 0.724 | Lecarpentier-2017 |

**Supplementary Table S11.** The AUCs generated from models that contain age, sex, top ten genetic principal components and the polygenic risk score specified.

| Disease | Base - NB | Base - Thresh | Score Incl. - NB | Score Incl. -Thresh |
|---|---|---|---|---|
| Lupus | 0 | 0.01 | 0.00232 | 0.01 |
| A. Fib. | 0.135 | 0.11 | 0.194 | 0.11 |
| Asthma | 0.0212 | 0.14 | 0.1 | 0.14 |
| Celiac Disease | 0 | 0.01 | 0.0302 | 0.01 |
| Migraine | -0.00657 | 0.07 | 0.0408 | 0.07 |
| MS | -0.00181 | 0.01 | 0.112 | 0.01 |
| Gout | 0.0326 | 0.06 | 0.113 | 0.06 |
| Crohns Disease | 0.117 | 0.01 | 0.213 | 0.01 |
| Ulcerative Colitis | 0 | 0.01 | 0.0569 | 0.01 |
| Type 2 Diabetes | 0.113 | 0.09 | 0.151 | 0.09 |
| Stroke | 0.0551 | 0.07 | 0.0703 | 0.07 |
| Breast Cancer | 0.0362 | 0.09 | 0.176 | 0.09 |
| NAFLD | 0.0211 | 0.01 | 0.0833 | 0.01 |
| CAD | 0.0488 | 0.11 | 0.108 | 0.11 |
| Rheumatoid Arthritis | 0.0699 | 0.03 | 0.106 | 0.03 |
| Type 1 Diabetes | 0.083 | 0.01 | 0.141 | 0.01 |
| Ovarian Cancer | 0.0404 | 0.01 | 0.0533 | 0.01 |
| Prostate Cancer | 0.039 | 0.1 | 0.199 | 0.1 |
| Heart Failure | 0.00612 | 0.09 | 0.294 | 0.09 |
| Psoriasis | 0.0174 | 0.02 | 0.241 | 0.02 |
| Depression | 0.00275 | 0.09 | 0.031 | 0.09 |

**Supplementary Table S12.** The maximum standardized net benefits corresponding to both base models that include the covariates of age, sex, and the top ten genetic principal components, and the score included models also includes the polygenic risk score to the base covariates.

| Disease | Base - NB at 0.01 | Base - NB at 0.05 | Base - NB at 0.12 | Score Incl. - NB at 0.01 | Score Incl. - NB at 0.05 | Score Incl. - NB at 0.12 |
|---|---|---|---|---|---|---|
| Lupus | 0 | 0 | 0 | 0.00232 | 0 | 0 |
| A. Fib. | 0.856 | 0.452 | 0.11 | 0.857 | 0.474 | 0.166 |
| Asthma | 0.934 | 0.658 | 0.122 | 0.934 | 0.658 | 0.185 |
| Celiac Disease | 0 | 0 | 0 | 0.0302 | -7.08e-05 | 0 |
| Migraine | 0.763 | 0.125 | 0 | 0.763 | 0.161 | 0.000488 |
| MS | -0.00181 | 0 | 0 | 0.112 | -0.00157 | 0 |
| Vitiligo | 0 | 0 | 0 | 0 | 0 | 0 |
| Gout | 0.699 | 0.107 | 0 | 0.706 | 0.174 | 0.0104 |
| Crohns Disease | 0.117 | 0 | 0 | 0.213 | 0.000297 | 0 |
| Ulcerative Colitis | 0 | 0 | 0 | 0.0569 | -5.97e-05 | 0 |
| Type 2 Diabetes | 0.86 | 0.375 | 0.0336 | 0.86 | 0.393 | 0.0683 |
| Stroke | 0.733 | 0.168 | 0.00111 | 0.732 | 0.17 | 0.00273 |
| Breast Cancer | 0.87 | 0.344 | -0.0013 | 0.869 | 0.416 | 0.0886 |
| NAFLD | 0.0211 | 0 | 0 | 0.0833 | 0 | 0 |
| CAD | 0.8 | 0.328 | 0.0335 | 0.804 | 0.365 | 0.0836 |
| Rheumatoid Arthritis | 0.566 | -0.00123 | 0 | 0.572 | 0.018 | 0.000152 |
| Type 1 Diabetes | 0.083 | 0 | 0 | 0.141 | -3.77e-05 | 0 |
| Ovarian Cancer | 0.0404 | 0 | 0 | 0.0533 | 0 | 0 |
| ALS | 0 | 0 | 0 | 0 | 0 | 0 |
| Prostate Cancer | 0.846 | 0.388 | -0.00534 | 0.853 | 0.459 | 0.134 |
| Heart Failure | 0.67 | 0.121 | -0.000205 | 0.677 | 0.352 | 0.273 |
| Psoriasis | 0.43 | 0 | 0 | 0.45 | 0.0309 | -0.00691 |
| Depression | 0.85 | 0.252 | 6.65e-05 | 0.85 | 0.269 | -0.00119 |

**Supplementary Table S13.** Standardized net benefits corresponding to both base models that include the covariates of age, sex, and the top ten genetic principal components, and the score included models also includes the polygenic risk score to the base covariates, computed at the thresholds described in the column headers.

| Disease | Start Threshold | End Threshold |
|---|---|---|
| Lupus | 0.01 | 0.01 |
| A. Fib. | 0.01 | 0.45 |
| Asthma | 0.08 | 0.22 |
| Celiac Disease | 0.01 | 0.03 |
| Migraine | 0.02 | 0.11 |
| MS | 0.01 | 0.03 |
| Gout | 0.01 | 0.13 |
| Crohns Disease | 0.01 | 0.02 |
| Ulcerative Colitis | 0.01 | 0.02 |
| Type 2 Diabetes | 0.02 | 0.24 |
| Stroke | 0.02 | 0.12 |
| Breast Cancer | 0.02 | 0.34 |
| NAFLD | 0.01 | 0.01 |
| CAD | 0.01 | 0.27 |
| Rheumatoid Arthritis | 0.01 | 0.08 |
| Type 1 Diabetes | 0.01 | 0.02 |
| Ovarian Cancer | 0.01 | 0.01 |
| Prostate Cancer | 0.01 | 0.44 |
| Heart Failure | 0.01 | 0.93 |
| Psoriasis | 0.01 | 0.07 |
| Depression | 0.04 | 0.11 |

**Supplementary Table S14.** The range of thresholds for each disease in which the score included model generated standardized net benefits that exceeded those of the base model. Base models include the covariates of age, sex, and the top ten genetic principal components, and the score included models also include the polygenic risk score to the base covariates

| Disease | Lifestyle Factor | | PRS Low | | | PRS Inter. | | | PRS High | |
|---|---|---|---|---|---|---|---|---|---|---|
| | | LF Low | LF Inter. | LF High | LF Low | LF Inter. | LF High | LF Low | LF Inter. | LF High |
| A. Fib. | Alcohol | 0.035 | 0.03 | 0.036 | 0.06 | 0.047 | 0.052 | 0.11 | 0.083 | 0.088 |
| A. Fib. | Smoking Status | 0.026 | 0.043 | 0.037 | 0.041 | 0.066 | 0.052 | 0.073 | 0.11 | 0.096 |
| A. Fib. | BMI | 0.022 | 0.029 | 0.056 | 0.032 | 0.048 | 0.077 | 0.056 | 0.088 | 0.13 |
| A. Fib. | Walking Pace | 0.091 | 0.034 | 0.021 | 0.12 | 0.053 | 0.035 | 0.19 | 0.094 | 0.065 |
| A. Fib. | TV | 0.025 | 0.031 | 0.046 | 0.04 | 0.049 | 0.069 | 0.072 | 0.094 | 0.11 |
| A. Fib. | Meat | 0.028 | 0.032 | 0.038 | 0.044 | 0.052 | 0.058 | 0.076 | 0.096 | 0.099 |
| A. Fib. | Water Intake | 0.034 | 0.034 | 0.03 | 0.052 | 0.051 | 0.049 | 0.098 | 0.089 | 0.079 |
| Asthma | Smoking Status | 0.022 | 0.03 | 0.036 | 0.027 | 0.035 | 0.04 | 0.035 | 0.048 | 0.052 |
| Asthma | BMI | 0.018 | 0.023 | 0.044 | 0.021 | 0.028 | 0.05 | 0.032 | 0.038 | 0.061 |
| Asthma | Walking Pace | 0.071 | 0.025 | 0.02 | 0.082 | 0.032 | 0.021 | 0.12 | 0.041 | 0.027 |
| Asthma | TV | 0.021 | 0.027 | 0.034 | 0.024 | 0.03 | 0.042 | 0.033 | 0.037 | 0.057 |
| Migraine | Alcohol | 0.0063 | 0.0062 | 0.011 | 0.008 | 0.0092 | 0.015 | 0.011 | 0.012 | 0.023 |
| Migraine | Hours Sleep | 0.01 | 0.0063 | 0.0069 | 0.013 | 0.0093 | 0.011 | 0.019 | 0.013 | 0.014 |
| MS | Walking Pace | 0.0021 | 0.00041 | 0.00039 | 0.0039 | 0.00091 | 0.00054 | 0.011 | 0.002 | 0.0022 |
| Gout | Alcohol | 0.011 | 0.006 | 0.0061 | 0.016 | 0.011 | 0.0087 | 0.028 | 0.018 | 0.016 |
| Gout | Smoking Status | 0.0046 | 0.011 | 0.0065 | 0.0077 | 0.016 | 0.012 | 0.014 | 0.027 | 0.024 |
| Gout | BMI | 0.0016 | 0.0061 | 0.015 | 0.0026 | 0.0096 | 0.024 | 0.0056 | 0.016 | 0.043 |
| Gout | Walking Pace | 0.024 | 0.0072 | 0.0035 | 0.034 | 0.011 | 0.0066 | 0.049 | 0.021 | 0.012 |
| Gout | TV | 0.0054 | 0.0072 | 0.0097 | 0.008 | 0.012 | 0.016 | 0.013 | 0.02 | 0.029 |
| Gout | Driving | 0.0063 | 0.0064 | 0.011 | 0.0092 | 0.012 | 0.015 | 0.017 | 0.021 | 0.024 |
| Gout | Fruit | 0.0093 | 0.0062 | 0.0055 | 0.013 | 0.011 | 0.0096 | 0.022 | 0.017 | 0.018 |
| Gout | Meat | 0.0043 | 0.0073 | 0.01 | 0.0074 | 0.011 | 0.016 | 0.012 | 0.021 | 0.027 |
| Crohns Disease | Walking Pace | 0.0056 | 0.0031 | 0.0021 | 0.01 | 0.0052 | 0.0043 | 0.01 | 0.0084 | 0.0064 |
| Ulcerative Colitis | Walking Pace | 0.0037 | 0.0012 | 0.0012 | 0.0047 | 0.002 | 0.0017 | 0.0095 | 0.0047 | 0.0036 |
| Ulcerative Colitis | TV | 0.0013 | 0.0011 | 0.0019 | 0.0016 | 0.0023 | 0.0027 | 0.0031 | 0.0048 | 0.0069 |
| Type 2 Diabetes | Alcohol | 0.03 | 0.03 | 0.058 | 0.044 | 0.047 | 0.083 | 0.064 | 0.071 | 0.12 |
| Type 2 Diabetes | Smoking Status | 0.028 | 0.047 | 0.055 | 0.042 | 0.072 | 0.077 | 0.066 | 0.1 | 0.1 |
| Type 2 Diabetes | BMI | 0.01 | 0.024 | 0.12 | 0.013 | 0.039 | 0.15 | 0.02 | 0.062 | 0.19 |
| Type 2 Diabetes | Min. Walked | 0.04 | 0.032 | 0.032 | 0.057 | 0.05 | 0.049 | 0.084 | 0.076 | 0.074 |
| Type 2 Diabetes | Mod. Activity | 0.049 | 0.033 | 0.034 | 0.074 | 0.048 | 0.049 | 0.1 | 0.075 | 0.073 |
| Type 2 Diabetes | Walking Pace | 0.13 | 0.041 | 0.017 | 0.18 | 0.061 | 0.026 | 0.25 | 0.091 | 0.038 |
| Type 2 Diabetes | TV | 0.023 | 0.037 | 0.062 | 0.035 | 0.055 | 0.09 | 0.054 | 0.083 | 0.13 |
| Type 2 Diabetes | Hours Sleep | 0.048 | 0.03 | 0.039 | 0.068 | 0.043 | 0.062 | 0.1 | 0.069 | 0.087 |
| Type 2 Diabetes | Vegetable | 0.039 | 0.036 | 0.035 | 0.06 | 0.053 | 0.051 | 0.09 | 0.081 | 0.076 |
| Type 2 Diabetes | Meat | 0.028 | 0.037 | 0.05 | 0.042 | 0.056 | 0.074 | 0.064 | 0.086 | 0.1 |
| Type 2 Diabetes | Cheese Intake | 0.045 | 0.042 | 0.034 | 0.068 | 0.06 | 0.05 | 0.1 | 0.089 | 0.074 |
| Type 2 Diabetes | Water Intake | 0.035 | 0.037 | 0.041 | 0.053 | 0.058 | 0.058 | 0.079 | 0.083 | 0.089 |
| Stroke | Smoking Status | 0.015 | 0.023 | 0.034 | 0.018 | 0.029 | 0.039 | 0.023 | 0.034 | 0.052 |
| Stroke | BMI | 0.016 | 0.019 | 0.024 | 0.018 | 0.024 | 0.031 | 0.023 | 0.029 | 0.036 |
| Stroke | Walking Pace | 0.053 | 0.021 | 0.012 | 0.068 | 0.024 | 0.016 | 0.08 | 0.029 | 0.02 |
| Stroke | TV | 0.015 | 0.017 | 0.029 | 0.016 | 0.024 | 0.036 | 0.023 | 0.028 | 0.041 |
| Stroke | Meat | 0.018 | 0.02 | 0.021 | 0.021 | 0.024 | 0.028 | 0.025 | 0.032 | 0.033 |
| Stroke | Water Intake | 0.021 | 0.019 | 0.017 | 0.026 | 0.023 | 0.022 | 0.03 | 0.032 | 0.025 |
| Breast Cancer | Walking Pace | 0.027 | 0.019 | 0.014 | 0.052 | 0.038 | 0.03 | 0.088 | 0.079 | 0.068 |
| Breast Cancer | TV | 0.013 | 0.017 | 0.023 | 0.034 | 0.035 | 0.038 | 0.067 | 0.073 | 0.086 |
| NAFLD | Alcohol | 0.0044 | 0.0052 | 0.01 | 0.0079 | 0.0063 | 0.012 | 0.0092 | 0.0093 | 0.017 |
| NAFLD | Smoking Status | 0.0058 | 0.0066 | 0.01 | 0.0068 | 0.0089 | 0.015 | 0.0091 | 0.013 | 0.017 |
| NAFLD | BMI | 0.0016 | 0.0048 | 0.016 | 0.0025 | 0.0057 | 0.022 | 0.0036 | 0.0091 | 0.026 |
| NAFLD | Mod. Activity | 0.0074 | 0.0064 | 0.0045 | 0.011 | 0.0073 | 0.007 | 0.014 | 0.01 | 0.0097 |
| NAFLD | Walking Pace | 0.021 | 0.0068 | 0.0034 | 0.026 | 0.0085 | 0.0046 | 0.033 | 0.013 | 0.0058 |
| NAFLD | TV | 0.0035 | 0.0074 | 0.01 | 0.0058 | 0.0078 | 0.012 | 0.008 | 0.01 | 0.018 |
| NAFLD | Cheese Intake | 0.0086 | 0.0076 | 0.0052 | 0.011 | 0.0089 | 0.0072 | 0.014 | 0.014 | 0.0096 |
| NAFLD | Water Intake | 0.0059 | 0.0061 | 0.0078 | 0.0079 | 0.0079 | 0.0093 | 0.0098 | 0.01 | 0.016 |
| CAD | Alcohol | 0.016 | 0.013 | 0.021 | 0.028 | 0.025 | 0.032 | 0.046 | 0.045 | 0.055 |
| CAD | Smoking Status | 0.01 | 0.023 | 0.027 | 0.019 | 0.035 | 0.051 | 0.034 | 0.059 | 0.085 |
| CAD | BMI | 0.0077 | 0.016 | 0.026 | 0.014 | 0.027 | 0.041 | 0.023 | 0.047 | 0.068 |
| CAD | Walking Pace | 0.052 | 0.016 | 0.01 | 0.081 | 0.028 | 0.016 | 0.12 | 0.049 | 0.029 |

| | | | | | | | | | | |
|---|---|---|---|---|---|---|---|---|---|---|
| CAD | TV | 0.012 | 0.017 | 0.022 | 0.021 | 0.028 | 0.038 | 0.034 | 0.047 | 0.067 |
| CAD | Driving | 0.015 | 0.016 | 0.019 | 0.026 | 0.027 | 0.031 | 0.043 | 0.05 | 0.058 |
| CAD | Vegetable | 0.016 | 0.016 | 0.016 | 0.03 | 0.025 | 0.025 | 0.052 | 0.045 | 0.043 |
| CAD | Fruit | 0.018 | 0.015 | 0.014 | 0.032 | 0.023 | 0.026 | 0.057 | 0.044 | 0.041 |
| CAD | Meat | 0.012 | 0.017 | 0.02 | 0.022 | 0.028 | 0.033 | 0.036 | 0.048 | 0.062 |
| CAD | Cheese Intake | 0.019 | 0.018 | 0.014 | 0.031 | 0.028 | 0.026 | 0.052 | 0.052 | 0.044 |
| CAD | Water Intake | 0.019 | 0.015 | 0.013 | 0.03 | 0.027 | 0.025 | 0.053 | 0.048 | 0.041 |
| Rheumatoid Arthritis | Alcohol | 0.0064 | 0.0069 | 0.013 | 0.0081 | 0.0099 | 0.016 | 0.012 | 0.014 | 0.02 |
| Rheumatoid Arthritis | Smoking Status | 0.0065 | 0.01 | 0.013 | 0.0095 | 0.013 | 0.017 | 0.012 | 0.019 | 0.022 |
| Rheumatoid Arthritis | BMI | 0.0056 | 0.0074 | 0.014 | 0.0084 | 0.011 | 0.016 | 0.012 | 0.014 | 0.023 |
| Rheumatoid Arthritis | Walking Pace | 0.03 | 0.0087 | 0.0043 | 0.036 | 0.011 | 0.007 | 0.048 | 0.014 | 0.0098 |
| Rheumatoid Arthritis | TV | 0.0048 | 0.0087 | 0.014 | 0.0084 | 0.01 | 0.016 | 0.011 | 0.017 | 0.02 |
| Rheumatoid Arthritis | Cheese Intake | 0.011 | 0.009 | 0.0071 | 0.014 | 0.012 | 0.01 | 0.02 | 0.016 | 0.013 |
| Type 1 Diabetes | Alcohol | 0.003 | 0.0032 | 0.0069 | 0.0034 | 0.0048 | 0.01 | 0.0081 | 0.0078 | 0.014 |
| Type 1 Diabetes | Walking Pace | 0.02 | 0.0036 | 0.0019 | 0.025 | 0.0053 | 0.003 | 0.026 | 0.0091 | 0.0067 |
| Type 1 Diabetes | TV | 0.0031 | 0.0029 | 0.007 | 0.0044 | 0.0049 | 0.0093 | 0.0074 | 0.0098 | 0.013 |
| Type 1 Diabetes | Meat | 0.0026 | 0.0051 | 0.0053 | 0.0047 | 0.0063 | 0.0074 | 0.0077 | 0.0073 | 0.014 |
| Prostate Cancer | Smoking Status | 0.016 | 0.02 | 0.013 | 0.037 | 0.046 | 0.036 | 0.088 | 0.098 | 0.062 |
| Prostate Cancer | TV | 0.015 | 0.014 | 0.023 | 0.037 | 0.042 | 0.044 | 0.083 | 0.091 | 0.096 |
| Prostate Cancer | Driving | 0.017 | 0.02 | 0.012 | 0.041 | 0.042 | 0.037 | 0.093 | 0.098 | 0.071 |
| Prostate Cancer | Hours Sleep | 0.014 | 0.015 | 0.021 | 0.037 | 0.038 | 0.046 | 0.076 | 0.091 | 0.096 |
| Heart Failure | Alcohol | 0.014 | 0.0095 | 0.013 | 0.016 | 0.012 | 0.018 | 0.065 | 0.055 | 0.081 |
| Heart Failure | Smoking Status | 0.0078 | 0.015 | 0.018 | 0.011 | 0.018 | 0.02 | 0.044 | 0.085 | 0.099 |
| Heart Failure | BMI | 0.0057 | 0.01 | 0.022 | 0.0075 | 0.012 | 0.028 | 0.038 | 0.057 | 0.099 |
| Heart Failure | Walking Pace | 0.039 | 0.012 | 0.0061 | 0.047 | 0.015 | 0.0076 | 0.2 | 0.062 | 0.031 |
| Heart Failure | TV | 0.0073 | 0.012 | 0.018 | 0.011 | 0.013 | 0.022 | 0.045 | 0.058 | 0.096 |
| Heart Failure | Meat | 0.0092 | 0.013 | 0.012 | 0.011 | 0.016 | 0.017 | 0.052 | 0.064 | 0.08 |
| Heart Failure | Cheese Intake | 0.013 | 0.013 | 0.01 | 0.016 | 0.016 | 0.014 | 0.073 | 0.069 | 0.059 |
| Psoriasis | Smoking Status | 0.0029 | 0.0054 | 0.0043 | 0.0044 | 0.0067 | 0.01 | 0.0097 | 0.018 | 0.02 |
| Psoriasis | BMI | 0.0021 | 0.0035 | 0.0067 | 0.004 | 0.0052 | 0.0089 | 0.0091 | 0.012 | 0.022 |
| Psoriasis | Walking Pace | 0.009 | 0.0042 | 0.0024 | 0.013 | 0.0058 | 0.0043 | 0.034 | 0.013 | 0.011 |
| Psoriasis | TV | 0.0031 | 0.0039 | 0.005 | 0.0048 | 0.0052 | 0.0076 | 0.011 | 0.013 | 0.018 |
| Psoriasis | Meat | 0.0033 | 0.0038 | 0.0046 | 0.0052 | 0.0057 | 0.0065 | 0.0092 | 0.014 | 0.018 |
| Depression | Smoking Status | 0.0081 | 0.01 | 0.009 | 0.0093 | 0.012 | 0.019 | 0.012 | 0.016 | 0.021 |
| Depression | Min. Walked | 0.01 | 0.0074 | 0.0078 | 0.011 | 0.011 | 0.0088 | 0.018 | 0.012 | 0.0078 |
| Depression | Mod. Activity | 0.011 | 0.0082 | 0.0091 | 0.015 | 0.0099 | 0.01 | 0.019 | 0.013 | 0.012 |
| Depression | Walking Pace | 0.02 | 0.0094 | 0.0066 | 0.025 | 0.01 | 0.0098 | 0.036 | 0.013 | 0.011 |
| Depression | Walking Pace | 0.0199 | 0.00944 | 0.00655 | 0.025 | 0.0101 | 0.00981 | 0.0356 | 0.0129 | 0.0108 |

**Supplementary Table S15.** The absolute risks calculated for each lifestyle and PRS grouping, listed for all significant disease and lifestyle factor combinations.

| Disease | Lifestyle Factor | P - Low | P - Inter. | P - High | OR - Low | OR - Inter. | OR - High |
|---|---|---|---|---|---|---|---|
| A. Fib. | Alcohol | 0.726 | 0.000163 | 0.000197 | 0.968 | 1.16 | 1.22 |
| A. Fib. | Smoking Status | 0.000538 | 9.91e-07 | 3.01e-05 | 0.677 | 0.77 | 0.748 |
| A. Fib. | BMI | 1.6e-24 | 1.1e-85 | 3.58e-45 | 0.374 | 0.398 | 0.408 |
| A. Fib. | Walking Pace | 1.59e-51 | 1.25e-150 | 1.63e-69 | 4.7 | 3.64 | 3.3 |
| A. Fib. | TV | 2.75e-19 | 3.99e-66 | 1.21e-28 | 0.523 | 0.56 | 0.602 |
| A. Fib. | Meat | 3.92e-05 | 1.45e-15 | 4.8e-10 | 0.733 | 0.755 | 0.744 |
| A. Fib. | Water Intake | 0.178 | 0.0532 | 6.96e-06 | 1.12 | 1.08 | 1.26 |
| Asthma | Smoking Status | 5.64e-05 | 1.54e-10 | 2.36e-05 | 0.608 | 0.66 | 0.655 |
| Asthma | BMI | 3.62e-15 | 2.46e-48 | 3.46e-14 | 0.412 | 0.413 | 0.501 |
| Asthma | Walking Pace | 2.01e-29 | 7.4e-109 | 2.4e-60 | 3.81 | 4.16 | 4.71 |
| Asthma | TV | 5.06e-09 | 3.44e-38 | 9.74e-17 | 0.606 | 0.559 | 0.57 |
| Migraine | Alcohol | 0.0028 | 3.22e-13 | 5.02e-08 | 0.572 | 0.515 | 0.49 |
| Migraine | Hours Sleep | 0.00956 | 0.0396 | 0.00514 | 1.53 | 1.18 | 1.38 |
| MS | Walking Pace | 0.0107 | 5.21e-11 | 1.01e-08 | 5.55 | 7.27 | 5.26 |
| Gout | Alcohol | 0.00063 | 6.63e-13 | 6.29e-07 | 1.86 | 1.85 | 1.75 |
| Gout | Smoking Status | 0.162 | 0.000177 | 9.76e-05 | 0.707 | 0.652 | 0.574 |
| Gout | BMI | 1.05e-18 | 2.19e-86 | 2.1e-47 | 0.106 | 0.105 | 0.127 |
| Gout | Walking Pace | 6.2e-20 | 3.22e-68 | 3.58e-27 | 6.87 | 5.41 | 4.27 |
| Gout | TV | 0.000197 | 1.05e-22 | 5.55e-18 | 0.559 | 0.497 | 0.443 |
| Gout | Driving | 0.000781 | 5.05e-10 | 0.00045 | 0.577 | 0.607 | 0.686 |
| Gout | Fruit | 0.00112 | 4.58e-06 | 0.0204 | 1.68 | 1.4 | 1.25 |
| Gout | Meat | 1.64e-07 | 1.32e-25 | 5.67e-17 | 0.42 | 0.46 | 0.432 |
| Crohns Disease | Walking Pace | 0.0062 | 6.76e-09 | 0.0405 | 2.69 | 2.4 | 1.64 |
| Ulcerative Colitis | Walking Pace | 0.00795 | 9.29e-06 | 0.000519 | 3.23 | 2.73 | 2.64 |
| Ulcerative Colitis | TV | 0.239 | 0.00123 | 2.73e-05 | 0.668 | 0.586 | 0.451 |
| Type 2 Diabetes | Alcohol | 1.7e-17 | 1.74e-68 | 9.8e-36 | 0.502 | 0.503 | 0.486 |
| Type 2 Diabetes | Smoking Status | 2.95e-12 | 4.69e-40 | 1.81e-12 | 0.501 | 0.533 | 0.614 |
| Type 2 Diabetes | BMI | 5.04e-156 | 0 | 2.15e-247 | 0.0794 | 0.0727 | 0.0847 |
| Type 2 Diabetes | Min. Walked | 0.00664 | 0.000572 | 0.0441 | 1.29 | 1.17 | 1.14 |
| Type 2 Diabetes | Mod. Activity | 2.51e-05 | 1.91e-24 | 6.87e-10 | 1.44 | 1.53 | 1.46 |
| Type 2 Diabetes | Walking Pace | 2.73e-110 | 0 | 3.61e-216 | 8.6 | 8.32 | 8.22 |
| Type 2 Diabetes | TV | 2.07e-50 | 4.94e-208 | 4.98e-85 | 0.36 | 0.367 | 0.398 |
| Type 2 Diabetes | Hours Sleep | 0.00328 | 0.00484 | 0.0013 | 1.24 | 1.1 | 1.18 |
| Type 2 Diabetes | Vegetable | 0.103 | 1.04e-07 | 0.000276 | 1.12 | 1.19 | 1.19 |
| Type 2 Diabetes | Meat | 6.76e-17 | 5.44e-71 | 1.34e-27 | 0.555 | 0.549 | 0.588 |
| Type 2 Diabetes | Cheese Intake | 6.21e-05 | 2.53e-21 | 8.67e-10 | 1.36 | 1.41 | 1.38 |
| Type 2 Diabetes | Water Intake | 0.0191 | 0.0132 | 0.0134 | 0.838 | 0.914 | 0.879 |
| Stroke | Smoking Status | 1.82e-10 | 1.53e-29 | 1.13e-17 | 0.433 | 0.465 | 0.421 |
| Stroke | BMI | 0.00161 | 2.96e-17 | 3.97e-05 | 0.663 | 0.564 | 0.645 |
| Stroke | Walking Pace | 7.69e-28 | 5.27e-113 | 6.54e-46 | 4.45 | 4.62 | 4.41 |
| Stroke | TV | 2.09e-13 | 5.57e-58 | 3.01e-15 | 0.511 | 0.455 | 0.553 |
| Stroke | Meat | 0.131 | 2.06e-09 | 0.000569 | 0.863 | 0.738 | 0.759 |
| Stroke | Water Intake | 0.042 | 0.00223 | 0.0327 | 1.24 | 1.18 | 1.21 |
| Breast Cancer | Walking Pace | 0.00217 | 1.72e-10 | 0.011 | 1.92 | 1.76 | 1.33 |
| Breast Cancer | TV | 0.000554 | 0.14 | 0.00205 | 0.544 | 0.902 | 0.767 |
| NAFLD | Alcohol | 2.34e-05 | 2.01e-06 | 2.81e-05 | 0.428 | 0.638 | 0.539 |
| NAFLD | Smoking Status | 0.0128 | 3.94e-13 | 0.000194 | 0.58 | 0.453 | 0.532 |
| NAFLD | BMI | 2.43e-21 | 1.04e-75 | 1.04e-28 | 0.0951 | 0.112 | 0.137 |
| NAFLD | Mod. Activity | 0.0263 | 4.56e-05 | 0.012 | 1.65 | 1.54 | 1.49 |
| NAFLD | Walking Pace | 2.04e-17 | 1.95e-55 | 1.76e-24 | 6.37 | 5.75 | 5.85 |
| NAFLD | TV | 5.47e-11 | 6.77e-21 | 4.32e-11 | 0.334 | 0.466 | 0.454 |
| NAFLD | Cheese Intake | 0.00339 | 9.91e-06 | 0.00609 | 1.68 | 1.49 | 1.45 |
| NAFLD | Water Intake | 0.136 | 0.0719 | 0.000349 | 0.761 | 0.849 | 0.63 |

| | | | | | | | |
|---|---|---|---|---|---|---|---|
| CAD | Alcohol | 0.0284 | 0.012 | 0.0137 | 0.767 | 0.87 | 0.83 |
| CAD | Smoking Status | 2.22e-11 | 1.42e-60 | 3.51e-32 | 0.37 | 0.361 | 0.378 |
| CAD | BMI | 2.01e-16 | 2.11e-59 | 1.24e-34 | 0.29 | 0.34 | 0.324 |
| CAD | Walking Pace | 1.64e-30 | 6.6e-151 | 1.02e-74 | 5.18 | 5.29 | 4.74 |
| CAD | TV | 1.12e-08 | 7.36e-45 | 1.14e-32 | 0.549 | 0.525 | 0.484 |
| CAD | Driving | 0.0257 | 0.00206 | 5.57e-06 | 0.766 | 0.85 | 0.73 |
| CAD | Vegetable | 0.832 | 5.17e-05 | 0.00313 | 1.03 | 1.21 | 1.2 |
| CAD | Fruit | 0.056 | 1.29e-05 | 3.62e-08 | 1.23 | 1.22 | 1.41 |
| CAD | Meat | 1.05e-05 | 2.26e-17 | 4.92e-18 | 0.617 | 0.667 | 0.578 |
| CAD | Cheese Intake | 0.0167 | 0.000858 | 0.0169 | 1.34 | 1.19 | 1.18 |
| CAD | Water Intake | 0.0013 | 5.08e-05 | 7.94e-05 | 1.45 | 1.23 | 1.31 |
| Rheumatoid Arthritis | Alcohol | 6.06e-05 | 6.08e-16 | 3.6e-05 | 0.498 | 0.491 | 0.582 |
| Rheumatoid Arthritis | Smoking Status | 0.000718 | 3.09e-09 | 2.01e-05 | 0.51 | 0.556 | 0.528 |
| Rheumatoid Arthritis | BMI | 1.19e-06 | 2.06e-11 | 1.8e-06 | 0.39 | 0.526 | 0.513 |
| Rheumatoid Arthritis | Walking Pace | 8.26e-25 | 7.96e-70 | 2.31e-31 | 7.27 | 5.3 | 5.11 |
| Rheumatoid Arthritis | TV | 5.99e-14 | 1.15e-21 | 8.76e-10 | 0.34 | 0.512 | 0.513 |
| Rheumatoid Arthritis | Cheese Intake | 0.00258 | 8.17e-05 | 0.000648 | 1.59 | 1.37 | 1.49 |
| Type 1 Diabetes | Alcohol | 0.000736 | 1.32e-19 | 0.00024 | 0.428 | 0.329 | 0.559 |
| Type 1 Diabetes | Walking Pace | 1.03e-21 | 4.11e-69 | 1.04e-14 | 10.8 | 8.63 | 3.97 |
| Type 1 Diabetes | TV | 1.3e-05 | 8.57e-16 | 2.07e-05 | 0.433 | 0.47 | 0.568 |
| Type 1 Diabetes | Meat | 0.00101 | 8.58e-06 | 2.42e-06 | 0.48 | 0.636 | 0.538 |
| Prostate Cancer | Smoking Status | 0.374 | 0.615 | 0.000425 | 1.26 | 1.05 | 1.47 |
| Prostate Cancer | TV | 0.00232 | 0.000795 | 0.0253 | 0.64 | 0.825 | 0.857 |
| Prostate Cancer | Driving | 0.0496 | 0.0745 | 0.000116 | 1.43 | 1.12 | 1.35 |
| Prostate Cancer | Hours Sleep | 0.0107 | 0.000349 | 0.000853 | 0.643 | 0.794 | 0.768 |
| Heart Failure | Alcohol | 0.672 | 0.0496 | 0.000156 | 1.07 | 0.868 | 0.791 |
| Heart Failure | Smoking Status | 3.71e-06 | 1.01e-10 | 3.51e-33 | 0.442 | 0.558 | 0.416 |
| Heart Failure | BMI | 1.49e-15 | 1.48e-56 | 1.43e-42 | 0.259 | 0.259 | 0.361 |
| Heart Failure | Walking Pace | 2.87e-27 | 1.01e-102 | 1.19e-191 | 6.53 | 6.37 | 7.75 |
| Heart Failure | TV | 1.82e-13 | 1.87e-30 | 3.25e-54 | 0.406 | 0.496 | 0.446 |
| Heart Failure | Meat | 0.0413 | 6.19e-12 | 2.52e-17 | 0.761 | 0.635 | 0.631 |
| Heart Failure | Cheese Intake | 0.0926 | 0.0642 | 6.73e-05 | 1.26 | 1.14 | 1.26 |
| Psoriasis | Smoking Status | 0.211 | 2.37e-11 | 8.7e-06 | 0.666 | 0.413 | 0.487 |
| Psoriasis | BMI | 7.46e-05 | 1.27e-09 | 6.3e-09 | 0.308 | 0.445 | 0.414 |
| Psoriasis | Walking Pace | 1.18e-05 | 4.53e-15 | 3.41e-14 | 3.75 | 3.01 | 3.22 |
| Psoriasis | TV | 0.0188 | 2.86e-06 | 5.01e-05 | 0.612 | 0.635 | 0.633 |
| Psoriasis | Meat | 0.128 | 0.0246 | 9.4e-09 | 0.71 | 0.795 | 0.505 |
| Depression | Smoking Status | 0.633 | 1.38e-12 | 4.8e-05 | 0.902 | 0.495 | 0.542 |
| Depression | Min. Walked | 0.14 | 0.0199 | 4.18e-07 | 1.33 | 1.27 | 2.33 |
| Depression | Mod. Activity | 0.328 | 1.84e-05 | 0.0013 | 1.19 | 1.48 | 1.59 |
| Depression | Walking Pace | 2.93e-07 | 1.75e-20 | 4.28e-16 | 3.08 | 2.59 | 3.37 |

**Supplementary Table S16.** The P-values and odds ratios computed from Fisher Exact tests that considered lifestyle factors within low, intermediate and high polygenic risk score groups. Listed are the disease and lifestyle factor combinations that were determined to be significant.

| Disease | Medication/Supplement | PRS Low | | PRS Inter. | | PRS High | |
|---|---|---|---|---|---|---|---|
| | | Off | On | Off | On | Off | On |
| A. Fib. | ibuprofen | 0.0343 | 0.0233 | 0.0535 | 0.034 | 0.0917 | 0.0736 |
| A. Fib. | evening primrose oil | 0.0332 | 0.0078 | 0.0513 | 0.0372 | 0.0899 | 0.062 |
| Type 2 Diabetes | ibuprofen | 0.0387 | 0.0299 | 0.0582 | 0.0423 | 0.0865 | 0.0655 |
| Type 2 Diabetes | evening primrose oil | 0.0377 | 0.0347 | 0.0566 | 0.0352 | 0.0847 | 0.0364 |
| Type 2 Diabetes | multivitamins | 0.038 | 0.0251 | 0.0566 | 0.0453 | 0.0845 | 0.0655 |
| Type 2 Diabetes | multivitamin+mineral preparations | 0.0379 | 0.0155 | 0.0564 | 0.0415 | 0.0842 | 0.0569 |
| CAD | ibuprofen | 0.0165 | 0.0129 | 0.0291 | 0.0175 | 0.0497 | 0.0345 |
| CAD | evening primrose oil | 0.0162 | 0.0099 | 0.0279 | 0.00874 | 0.0483 | 0.0172 |
| Heart Failure | ibuprofen | 0.0118 | 0.00715 | 0.0154 | 0.00905 | 0.0683 | 0.0384 |
| Heart Failure | evening primrose oil | 0.0113 | 0.00821 | 0.0148 | 0.00617 | 0.0653 | 0.0188 |

**Supplementary Table S17.** The absolute risks calculated for each medication/supplement and PRS grouping, listed for all significant disease and lifestyle factor combinations. On means the group of people on or taking the medication/supplement.

| Disease | Medication/Supplement | P-Low | P-Inter. | P-High | OR-Low | OR-Inter. | OR-High |
|---|---|---|---|---|---|---|---|
| A. Fib. | ibuprofen | 0.000129 | 3.27e-21 | 0.000144 | 1.49 | 1.6 | 1.27 |
| A. Fib. | evening primrose oil | 0.000379 | 0.0101 | 0.0325 | 4.38 | 1.4 | 1.49 |
| Type 2 Diabetes | ibuprofen | 0.00635 | 2.39e-13 | 4.69e-06 | 1.3 | 1.4 | 1.35 |
| Type 2 Diabetes | evening primrose oil | 0.816 | 0.000117 | 2.09e-05 | 1.09 | 1.64 | 2.45 |
| Type 2 Diabetes | multivitamins | 0.0305 | 0.00747 | 0.0325 | 1.53 | 1.26 | 1.32 |
| Type 2 Diabetes | multivitamin+mineral preparations | 0.00855 | 0.0171 | 0.042 | 2.5 | 1.38 | 1.53 |
| CAD | ibuprofen | 0.0913 | 1.86e-14 | 1.36e-05 | 1.29 | 1.69 | 1.47 |
| CAD | evening primrose oil | 0.369 | 1.28e-07 | 0.000377 | 1.64 | 3.26 | 2.89 |
| Heart Failure | ibuprofen | 0.00935 | 9.44e-09 | 1.24e-14 | 1.66 | 1.71 | 1.84 |
| Heart Failure | evening primrose oil | 0.667 | 0.0023 | 1.13e-06 | 1.38 | 2.41 | 3.66 |

**Supplementary Table S18.** The P-values and odds ratios computed from Fisher Exact tests that considered medication/supplements within low, intermediate and high polygenic risk score groups. Listed are the disease and medication/supplement combinations that were determined to be significant.

| Disease | AUC Male | AUC CI Male | AUC Female | AUC CI Female |
|---|---|---|---|---|
| A. Fib. | 0.731 | 0.006 | 0.751 | 0.007 |
| ALS | 0.612 | 0.049 | 0.61 | 0.06 |
| Asthma | 0.593 | 0.006 | 0.577 | 0.006 |
| CAD | 0.718 | 0.005 | 0.723 | 0.01 |
| Celiac Disease | 0.628 | 0.034 | 0.65 | 0.024 |
| Crohns Disease | 0.612 | 0.017 | 0.618 | 0.018 |
| Depression | 0.57 | 0.01 | 0.581 | 0.007 |
| Gout | 0.662 | 0.009 | 0.719 | 0.02 |
| Heart Failure | 0.815 | 0.009 | 0.793 | 0.013 |
| Lupus | 0.656 | 0.09 | 0.618 | 0.04 |
| Migraine | 0.584 | 0.014 | 0.597 | 0.008 |
| MS | 0.696 | 0.035 | 0.686 | 0.022 |
| NAFLD | 0.568 | 0.021 | 0.555 | 0.021 |
| Psoriasis | 0.672 | 0.014 | 0.671 | 0.016 |
| Rheumatoid Arthritis | 0.646 | 0.016 | 0.633 | 0.011 |
| Stroke | 0.687 | 0.008 | 0.679 | 0.01 |
| Type 1 Diabetes | 0.609 | 0.018 | 0.62 | 0.022 |
| Type 2 Diabetes | 0.658 | 0.006 | 0.663 | 0.008 |
| Ulcerative Colitis | 0.665 | 0.026 | 0.637 | 0.025 |
| Vitiligo | 0.666 | 0.083 | 0.623 | 0.059 |

**Supplementary Table S19.** The AUC and their 95% CI generated from models that utilized either only male or female individuals.

| Disease | AUC Young | AUC CI Young | AUC Old | AUC CI Old |
|---|---|---|---|---|
| Lupus | 0.719 | 0.039 | 0.694 | 0.045 |
| A. Fib. | 0.742 | 0.006 | 0.658 | 0.008 |
| Asthma | 0.591 | 0.006 | 0.583 | 0.006 |
| Celiac Disease | 0.679 | 0.026 | 0.654 | 0.027 |
| Migraine | 0.662 | 0.009 | 0.656 | 0.009 |
| MS | 0.718 | 0.025 | 0.709 | 0.026 |
| Vitiligo | 0.677 | 0.065 | 0.643 | 0.074 |
| Gout | 0.804 | 0.008 | 0.748 | 0.01 |
| Crohns Disease | 0.615 | 0.018 | 0.603 | 0.018 |
| Ulcerative Colitis | 0.662 | 0.025 | 0.641 | 0.025 |
| Type 2 Diabetes | 0.671 | 0.007 | 0.635 | 0.008 |
| Stroke | 0.656 | 0.01 | 0.598 | 0.01 |
| Breast Cancer | 0.645 | 0.009 | 0.645 | 0.009 |
| NAFLD | 0.57 | 0.02 | 0.568 | 0.021 |
| CAD | 0.77 | 0.006 | 0.714 | 0.007 |
| Rheumatoid Arthritis | 0.651 | 0.012 | 0.614 | 0.013 |
| Type 1 Diabetes | 0.646 | 0.021 | 0.605 | 0.021 |
| Ovarian Cancer | 0.551 | 0.029 | 0.562 | 0.03 |
| ALS | 0.617 | 0.053 | 0.543 | 0.057 |
| Prostate Cancer | 0.782 | 0.009 | 0.667 | 0.012 |
| Heart Failure | 0.804 | 0.011 | 0.763 | 0.013 |
| Psoriasis | 0.691 | 0.015 | 0.666 | 0.016 |
| Depression | 0.593 | 0.008 | 0.598 | 0.008 |

**Supplementary Table S20.** The AUC and their 95% CI generated from models that utilized either only young or old individuals as determined by the median age of cases for each respective disease.

| Disease | AUC - Low Group | AUC CI - Low Group | AUC - Hi Group | AUC CI - Hi Group |
|---|---|---|---|---|
| Lupus | 0.717 | 0.039 | 0.706 | 0.047 |
| A. Fib. | 0.78 | 0.007 | 0.727 | 0.006 |
| Asthma | 0.588 | 0.006 | 0.584 | 0.007 |
| Celiac Disease | 0.669 | 0.026 | 0.662 | 0.029 |
| Migraine | 0.67 | 0.009 | 0.654 | 0.01 |
| MS | 0.719 | 0.023 | 0.707 | 0.03 |
| Vitiligo | 0.694 | 0.072 | 0.605 | 0.071 |
| Gout | 0.807 | 0.008 | 0.773 | 0.009 |
| Crohns Disease | 0.621 | 0.017 | 0.617 | 0.019 |
| Ulcerative Colitis | 0.657 | 0.025 | 0.648 | 0.028 |
| Type 2 Diabetes | 0.698 | 0.007 | 0.658 | 0.008 |
| Stroke | 0.711 | 0.009 | 0.666 | 0.009 |
| Breast Cancer | 0.657 | 0.009 | 0.649 | 0.009 |
| NAFLD | 0.583 | 0.02 | 0.553 | 0.023 |
| CAD | 0.789 | 0.007 | 0.755 | 0.006 |
| Rheumatoid Arthritis | 0.678 | 0.012 | 0.624 | 0.013 |
| Type 1 Diabetes | 0.655 | 0.019 | 0.632 | 0.022 |
| Ovarian Cancer | 0.61 | 0.031 | 0.597 | 0.03 |
| ALS | 0.67 | 0.061 | 0.582 | 0.051 |
| Prostate Cancer | 0.798 | 0.009 | 0.722 | 0.01 |
| Heart Failure | 0.833 | 0.01 | 0.796 | 0.011 |
| Psoriasis | 0.687 | 0.014 | 0.668 | 0.017 |
| Depression | 0.594 | 0.007 | 0.612 | 0.009 |

**Supplementary Table S21.** Validity check by splitting the total population according to time at Current Address. The low group is less than or equal to 20 years and the higher group is greater than 20 years.

| Disease | AUC - Low Group | AUC CI - Low Group | AUC - Hi Group | AUC CI - Hi Group |
|---|---|---|---|---|
| Lupus | 0.703 | 0.041 | 0.693 | 0.049 |
| A. Fib. | 0.733 | 0.006 | 0.777 | 0.008 |
| Asthma | 0.581 | 0.006 | 0.599 | 0.006 |
| Celiac Disease | 0.644 | 0.031 | 0.675 | 0.029 |
| Migraine | 0.665 | 0.01 | 0.664 | 0.009 |
| MS | 0.709 | 0.027 | 0.723 | 0.028 |
| Vitiligo | 0.631 | 0.084 | 0.686 | 0.066 |
| Gout | 0.773 | 0.01 | 0.807 | 0.009 |
| Crohns Disease | 0.615 | 0.018 | 0.619 | 0.02 |
| Ulcerative Colitis | 0.666 | 0.026 | 0.649 | 0.03 |
| Type 2 Diabetes | 0.656 | 0.007 | 0.698 | 0.009 |
| Stroke | 0.658 | 0.009 | 0.701 | 0.013 |
| Breast Cancer | 0.655 | 0.009 | 0.656 | 0.01 |
| NAFLD | 0.566 | 0.021 | 0.575 | 0.025 |
| CAD | 0.754 | 0.007 | 0.793 | 0.007 |
| Rheumatoid Arthritis | 0.617 | 0.013 | 0.652 | 0.018 |
| Type 1 Diabetes | 0.622 | 0.02 | 0.649 | 0.027 |
| Ovarian Cancer | 0.585 | 0.03 | 0.584 | 0.039 |
| ALS | 0.587 | 0.057 | 0.67 | 0.06 |
| Prostate Cancer | 0.743 | 0.01 | 0.791 | 0.01 |
| Heart Failure | 0.796 | 0.011 | 0.826 | 0.014 |
| Psoriasis | 0.681 | 0.016 | 0.679 | 0.016 |
| Depression | 0.632 | 0.007 | 0.602 | 0.009 |

**Supplementary Table S22.** Validity check by splitting the total population according to income. The low group is less than £40,000 and the higher group is greater than £40,000.

| Disease | AUC - Low Group | AUC CI - Low Group | AUC - Hi Group | AUC CI - Hi Group |
|---|---|---|---|---|
| Lupus | 0.731 | 0.034 | 0.656 | 0.058 |
| A. Fib. | 0.736 | 0.005 | 0.778 | 0.01 |
| Asthma | 0.584 | 0.005 | 0.594 | 0.007 |
| Celiac Disease | 0.656 | 0.023 | 0.685 | 0.033 |
| Migraine | 0.661 | 0.008 | 0.665 | 0.011 |
| MS | 0.721 | 0.022 | 0.713 | 0.031 |
| Vitiligo | 0.651 | 0.064 | 0.637 | 0.078 |
| Gout | 0.78 | 0.007 | 0.812 | 0.011 |
| Crohns Disease | 0.619 | 0.015 | 0.609 | 0.024 |
| Ulcerative Colitis | 0.652 | 0.021 | 0.641 | 0.033 |
| Type 2 Diabetes | 0.658 | 0.006 | 0.712 | 0.011 |
| Stroke | 0.667 | 0.007 | 0.704 | 0.016 |
| Breast Cancer | 0.654 | 0.007 | 0.643 | 0.012 |
| NAFLD | 0.569 | 0.018 | 0.57 | 0.028 |
| CAD | 0.759 | 0.006 | 0.794 | 0.01 |
| Rheumatoid Arthritis | 0.637 | 0.011 | 0.658 | 0.019 |
| Type 1 Diabetes | 0.618 | 0.017 | 0.673 | 0.027 |
| Ovarian Cancer | 0.578 | 0.024 | 0.574 | 0.047 |
| ALS | 0.618 | 0.042 | 0.653 | 0.079 |
| Prostate Cancer | 0.74 | 0.008 | 0.806 | 0.013 |
| Heart Failure | 0.8 | 0.009 | 0.836 | 0.016 |
| Psoriasis | 0.68 | 0.013 | 0.679 | 0.019 |
| Depression | 0.616 | 0.007 | 0.604 | 0.01 |

**Supplementary Table S23.** Validity check by splitting the total population according to number of individuals in the household. The low group is 1-2 persons and the higher group is 3 or more persons.

| Disease | AUC - Low Group | AUC CI - Low Group | AUC - Hi Group | AUC CI - Hi Group |
|---|---|---|---|---|
| Lupus | 0.718 | 0.036 | 0.566 | 0.192 |
| A. Fib. | 0.752 | 0.005 | 0.743 | 0.02 |
| Asthma | 0.583 | 0.005 | 0.589 | 0.017 |
| Celiac Disease | 0.661 | 0.025 | 0.663 | 0.07 |
| Migraine | 0.667 | 0.008 | 0.648 | 0.026 |
| MS | 0.713 | 0.023 | 0.745 | 0.075 |
| Vitiligo | 0.615 | 0.07 | 0.505 | 0.119 |
| Gout | 0.786 | 0.008 | 0.78 | 0.025 |
| Crohns Disease | 0.616 | 0.015 | 0.601 | 0.052 |
| Ulcerative Colitis | 0.647 | 0.022 | 0.603 | 0.081 |
| Type 2 Diabetes | 0.674 | 0.006 | 0.689 | 0.021 |
| Stroke | 0.683 | 0.008 | 0.677 | 0.027 |
| Breast Cancer | 0.654 | 0.008 | 0.634 | 0.026 |
| NAFLD | 0.557 | 0.017 | 0.567 | 0.068 |
| CAD | 0.768 | 0.005 | 0.778 | 0.018 |
| Rheumatoid Arthritis | 0.637 | 0.011 | 0.664 | 0.037 |
| Type 1 Diabetes | 0.634 | 0.017 | 0.625 | 0.067 |
| Ovarian Cancer | 0.597 | 0.026 | 0.572 | 0.089 |
| ALS | 0.628 | 0.047 | 0.523 | 0.127 |
| Prostate Cancer | 0.758 | 0.009 | 0.766 | 0.025 |
| Heart Failure | 0.809 | 0.009 | 0.817 | 0.031 |
| Psoriasis | 0.692 | 0.013 | 0.661 | 0.045 |
| Depression | 0.61 | 0.007 | 0.608 | 0.024 |

**Supplementary Table S24.** Validity check by splitting the total population according to the age education was completed. The low group is 1-19 years and the higher group is 20 or more years.

| Disease | AUC - Low Group | AUC CI - Low Group | AUC - Hi Group | AUC CI - Hi Group |
|---|---|---|---|---|
| Lupus | 0.731 | 0.042 | 0.691 | 0.041 |
| A. Fib. | 0.756 | 0.007 | 0.763 | 0.006 |
| Asthma | 0.588 | 0.006 | 0.585 | 0.005 |
| Celiac Disease | 0.649 | 0.029 | 0.677 | 0.025 |
| Migraine | 0.668 | 0.01 | 0.66 | 0.008 |
| MS | 0.725 | 0.028 | 0.71 | 0.024 |
| Vitiligo | 0.683 | 0.066 | 0.651 | 0.075 |
| Gout | 0.798 | 0.009 | 0.788 | 0.008 |
| Crohns Disease | 0.612 | 0.019 | 0.618 | 0.017 |
| Ulcerative Colitis | 0.64 | 0.028 | 0.657 | 0.024 |
| Type 2 Diabetes | 0.687 | 0.007 | 0.681 | 0.006 |
| Stroke | 0.691 | 0.01 | 0.699 | 0.008 |
| Breast Cancer | 0.655 | 0.01 | 0.655 | 0.009 |
| NAFLD | 0.569 | 0.025 | 0.565 | 0.019 |
| CAD | 0.774 | 0.007 | 0.776 | 0.006 |
| Rheumatoid Arthritis | 0.647 | 0.014 | 0.66 | 0.011 |
| Type 1 Diabetes | 0.631 | 0.023 | 0.635 | 0.018 |
| Ovarian Cancer | 0.623 | 0.032 | 0.596 | 0.028 |
| ALS | 0.631 | 0.055 | 0.65 | 0.049 |
| Prostate Cancer | 0.756 | 0.01 | 0.782 | 0.009 |
| Heart Failure | 0.815 | 0.012 | 0.819 | 0.009 |
| Psoriasis | 0.676 | 0.017 | 0.681 | 0.014 |
| Depression | 0.611 | 0.009 | 0.598 | 0.008 |

**Supplementary Table S25.** Validity check by splitting the total population according to the census measurement of median age. The lower group is less than 42 years and the higher group is greater than 42 years.

| Disease | AUC - Low Group | AUC CI - Low Group | AUC - Hi Group | AUC CI - Hi Group |
|---|---|---|---|---|
| Lupus | 0.709 | 0.04 | 0.715 | 0.044 |
| A. Fib. | 0.762 | 0.007 | 0.758 | 0.006 |
| Asthma | 0.586 | 0.006 | 0.588 | 0.005 |
| Celiac Disease | 0.667 | 0.028 | 0.661 | 0.027 |
| Migraine | 0.664 | 0.009 | 0.665 | 0.009 |
| MS | 0.716 | 0.026 | 0.717 | 0.025 |
| Vitiligo | 0.628 | 0.074 | 0.68 | 0.066 |
| Gout | 0.786 | 0.009 | 0.8 | 0.008 |
| Crohns Disease | 0.628 | 0.018 | 0.61 | 0.018 |
| Ulcerative Colitis | 0.659 | 0.026 | 0.643 | 0.026 |
| Type 2 Diabetes | 0.676 | 0.007 | 0.695 | 0.008 |
| Stroke | 0.694 | 0.009 | 0.698 | 0.01 |
| Breast Cancer | 0.658 | 0.009 | 0.65 | 0.008 |
| NAFLD | 0.579 | 0.019 | 0.554 | 0.023 |
| CAD | 0.769 | 0.006 | 0.782 | 0.007 |
| Rheumatoid Arthritis | 0.654 | 0.012 | 0.658 | 0.013 |
| Type 1 Diabetes | 0.631 | 0.02 | 0.633 | 0.022 |
| Ovarian Cancer | 0.595 | 0.03 | 0.62 | 0.029 |
| ALS | 0.615 | 0.058 | 0.659 | 0.047 |
| Prostate Cancer | 0.769 | 0.01 | 0.769 | 0.009 |
| Heart Failure | 0.817 | 0.01 | 0.819 | 0.011 |
| Psoriasis | 0.684 | 0.015 | 0.674 | 0.016 |
| Depression | 0.601 | 0.008 | 0.604 | 0.008 |

**Supplementary Table S26.** Validity check by splitting the total population according to the census measurement of unemployment. The lower group is less than 38 persons and the higher group is greater than 38 persons.

| Disease | AUC - Low Group | AUC CI - Low Group | AUC - Hi Group | AUC CI - Hi Group |
|---|---|---|---|---|
| Lupus | 0.688 | 0.045 | 0.731 | 0.04 |
| A. Fib. | 0.76 | 0.007 | 0.76 | 0.007 |
| Asthma | 0.585 | 0.006 | 0.589 | 0.005 |
| Celiac Disease | 0.659 | 0.028 | 0.669 | 0.027 |
| Migraine | 0.66 | 0.009 | 0.668 | 0.009 |
| MS | 0.711 | 0.025 | 0.722 | 0.025 |
| Vitiligo | 0.661 | 0.07 | 0.653 | 0.077 |
| Gout | 0.805 | 0.009 | 0.78 | 0.009 |
| Crohns Disease | 0.606 | 0.018 | 0.62 | 0.018 |
| Ulcerative Colitis | 0.648 | 0.025 | 0.648 | 0.026 |
| Type 2 Diabetes | 0.686 | 0.007 | 0.68 | 0.006 |
| Stroke | 0.698 | 0.009 | 0.691 | 0.009 |
| Breast Cancer | 0.657 | 0.008 | 0.651 | 0.009 |
| NAFLD | 0.575 | 0.021 | 0.563 | 0.02 |
| CAD | 0.779 | 0.006 | 0.77 | 0.007 |
| Rheumatoid Arthritis | 0.658 | 0.013 | 0.649 | 0.012 |
| Type 1 Diabetes | 0.647 | 0.021 | 0.632 | 0.019 |
| Ovarian Cancer | 0.617 | 0.03 | 0.593 | 0.028 |
| ALS | 0.652 | 0.054 | 0.627 | 0.051 |
| Prostate Cancer | 0.778 | 0.009 | 0.758 | 0.009 |
| Heart Failure | 0.81 | 0.011 | 0.824 | 0.01 |
| Psoriasis | 0.688 | 0.015 | 0.669 | 0.015 |
| Depression | 0.599 | 0.008 | 0.606 | 0.008 |

**Supplementary Table S27.** Validity check by splitting the total population according to census measurement of very good health. The lower group is less than 719 persons and the higher group is greater than 719 persons.

| Disease | AUC - Low Group | AUC CI - Low Group | AUC - Hi Group | AUC CI - Hi Group |
|---|---|---|---|---|
| Lupus | 0.707 | 0.042 | 0.716 | 0.043 |
| A. Fib. | 0.766 | 0.007 | 0.754 | 0.006 |
| Asthma | 0.586 | 0.006 | 0.586 | 0.005 |
| Celiac Disease | 0.685 | 0.026 | 0.648 | 0.028 |
| Migraine | 0.663 | 0.009 | 0.664 | 0.009 |
| MS | 0.716 | 0.026 | 0.717 | 0.025 |
| Vitiligo | 0.656 | 0.076 | 0.652 | 0.07 |
| Gout | 0.783 | 0.009 | 0.801 | 0.008 |
| Crohns Disease | 0.618 | 0.018 | 0.613 | 0.018 |
| Ulcerative Colitis | 0.669 | 0.025 | 0.629 | 0.028 |
| Type 2 Diabetes | 0.678 | 0.008 | 0.689 | 0.007 |
| Stroke | 0.698 | 0.009 | 0.693 | 0.01 |
| Breast Cancer | 0.655 | 0.009 | 0.654 | 0.009 |
| NAFLD | 0.568 | 0.02 | 0.563 | 0.023 |
| CAD | 0.776 | 0.007 | 0.774 | 0.007 |
| Rheumatoid Arthritis | 0.655 | 0.012 | 0.652 | 0.013 |
| Type 1 Diabetes | 0.635 | 0.02 | 0.633 | 0.021 |
| Ovarian Cancer | 0.598 | 0.029 | 0.616 | 0.03 |
| ALS | 0.65 | 0.049 | 0.627 | 0.055 |
| Prostate Cancer | 0.772 | 0.01 | 0.766 | 0.009 |
| Heart Failure | 0.817 | 0.01 | 0.817 | 0.011 |
| Psoriasis | 0.681 | 0.015 | 0.676 | 0.015 |
| Depression | 0.605 | 0.008 | 0.6 | 0.008 |

**Supplementary Table S28.** Validity check by splitting the total population according to the census measurement of population density. The lower group is less than 32 persons per hectare and the higher group is great than 32 persons per heactare.

| Disease | European | African | Asian |
|---|---|---|---|
| Lupus | -10.4 | 5.22 | -2.67 |
| A. Fib. | 2.06 | -5.01 | 0.784 |
| Asthma | 1.05 | -3.71 | 0.355 |
| Celiac Disease | -0.879 | 0.635 | 0.711 |
| Migraine | 2.74 | 0.91 | -0.717 |
| MS | 25.5 | 12.2 | 30 |
| Vitiligo | -2.18 | 3.17 | -4.2 |
| Gout | 22 | 34.4 | 21.7 |
| Crohns Disease | 5.05 | -5.92 | 1.13 |
| Ulcerative Colitis | -7.36 | -4.05 | -4.63 |
| Type 2 Diabetes | -3.92 | -11.9 | -5.31 |
| Stroke | -20.7 | -36.4 | -11.1 |
| Breast Cancer | -1.25 | -8.66 | 3.07 |
| NAFLD | -24.7 | -34.7 | -23.6 |
| CAD | 3.01 | 3.71 | 3 |
| Rheumatoid Arthritis | 22.3 | 12.6 | 24.4 |
| Type 1 Diabetes | -8.52 | -6.24 | -8.57 |
| Ovarian Cancer | -4.01 | -6.1 | -5.51 |
| ALS | -5.08 | -4.6 | -2.99 |
| Prostate Cancer | 2.03 | -10.9 | 9.71 |
| Heart Failure | 4.3 | 13.1 | -8.66 |
| Psoriasis | 8.91 | -6.15 | 1.76 |
| Depression | 8.81 | -1.88 | 10 |

**Supplementary Table S29.** The mean of each population group's polygenic risk score (that was utilized in the testing) phase minus the mean of the British population group's polygenic risk score. Before the mean was taken each polygenic risk score was standardized to have a minimum of zero and maximum of 100. No additional standardization/scaling was done.

| Disease | British-European | British-Asian | British-African | European-African | European-Asian | African-Asian |
|---|---|---|---|---|---|---|
| Lupus | 4.83e-199 | 1e-300 | 1e-300 | 1e-300 | 1e-300 | 1e-300 |
| A. Fib. | 1e-300 | 7.24e-217 | 1e-300 | 1e-300 | 2.96e-20 | 4.84e-198 |
| Asthma | 1e-300 | 1.14e-145 | 1.59e-15 | 4.81e-242 | 4.04e-08 | 5.89e-120 |
| Celiac Disease | 1e-300 | 3.82e-26 | 3.49e-17 | 1.92e-22 | 6.33e-40 | 0.68 |
| Migraine | 1e-300 | 9.09e-08 | 4.63e-12 | 1.98e-33 | 5.09e-159 | 1.21e-18 |
| Vitiligo | 0.00267 | 4.33e-125 | 1e-300 | 1e-300 | 1.38e-79 | 1e-300 |
| Gout | 0.014 | 0.624 | 1e-300 | 1e-300 | 0.118 | 1e-300 |
| Crohns Disease | 1e-300 | 8.69e-12 | 1e-300 | 1e-300 | 1.08e-209 | 1e-300 |
| Ulcerative Colitis | 1e-300 | 2.2e-54 | 1.54e-19 | 1.1e-118 | 2.27e-107 | 0.000951 |
| Type 2 Diabetes | 1e-300 | 9.08e-49 | 1.47e-276 | 1e-300 | 1.12e-28 | 1e-300 |
| Stroke | 3.84e-07 | 1e-300 | 1e-300 | 1e-300 | 1e-300 | 1e-300 |
| Breast Cancer | 1e-300 | 2.75e-27 | 1e-300 | 1e-300 | 2.48e-116 | 1e-300 |
| NAFLD | 0.000654 | 4.89e-13 | 1e-300 | 1e-300 | 3.59e-07 | 1e-300 |
| CAD | 1e-300 | 1e-300 | 6.42e-286 | 1.73e-06 | 0.969 | 8.15e-05 |
| Rheumatoid Arthritis | 1e-300 | 1e-300 | 1e-300 | 1e-300 | 1.92e-188 | 1e-300 |
| Type 1 Diabetes | 1e-300 | 5.72e-100 | 0.918 | 1.35e-59 | 0.696 | 4.15e-43 |
| Ovarian Cancer | 4.79e-154 | 0.00664 | 0.164 | 8.51e-34 | 2.47e-23 | 0.0043 |
| Prostate Cancer | 5.54e-225 | 1e-300 | 1e-300 | 1e-300 | 1e-300 | 1e-300 |
| Heart Failure | 1e-300 | 1e-300 | 1.41e-102 | 1e-300 | 1e-300 | 1e-300 |
| Psoriasis | 2.72e-34 | 1e-300 | 1e-300 | 1e-300 | 1e-300 | 1e-300 |
| Depression | 1e-300 | 7.56e-158 | 1e-300 | 1e-300 | 3.49e-22 | 1e-300 |

**Supplementary Table S30.** The P-values generated when comparing polygenic risk score distributions of one population group to another, as indicated in the column header, with a Student's T-Test. The polygenic risk score distributions used in the test are the same as the distributions described in the previous table. P-values less than 1e-300 are reported at 1e-300.

| Disease | ICD | Self-Reported | Any | Double-Reported |
|---|---|---|---|---|
| Lupus | 0.000402 | 0 | 0 | 0 |
| A. Fib. | 0.00151 | 0 | -3.34E-06 | 0 |
| Asthma | 0.00208 | 0 | -1.29E-07 | 0 |
| Celiac Disease | 0.244 | 0 | 0 | 0 |
| Migraine | -0.00971 | 0 | 0.000369 | 0 |
| MS | -0.000993 | 0 | 0.000249 | 0 |
| Vitiligo | -0.0771 | 0 | 0 | 0 |
| Gout | -0.00449 | 0 | 4.77E-05 | 0 |
| Crohns Disease | 0.00199 | 0 | 2.55E-05 | 0 |
| Ulcerative Colitis | 0.00595 | 0 | 0 | 0 |
| Type 2 Diabetes | 0.00144 | 0 | 0 | 0 |
| Stroke | 0.00181 | 0 | 0 | 0 |
| Breast Cancer | 0.0177 | 0 | -1.55E-05 | 0 |
| NAFLD | 0 | 0 | 0 | 0 |
| CAD | -0.0115 | 0 | 0 | 0 |
| Rheumatoid Arthritis | 0.0133 | 0 | -0.00253 | 0 |
| Type 1 Diabetes | 0.00102 | 0 | 0 | 0 |
| Ovarian Cancer | 0.00411 | 0 | 0 | 0 |
| ALS | 0.000965 | 0 | -0.00972 | 0 |
| Prostate Cancer | -0.00292 | 0 | -0.000384 | 0 |
| Heart Failure | -0.000993 | 0 | 0 | 0 |
| Psoriasis | -0.0138 | 0 | 0 | 0 |
| Depression | -0.017 | 0 | 0 | 0 |

**Supplementary Table S31.** The AUC values generated from models assessed on the testing set, with phenotype defintions as described in the column headers, minus the AUC values generated from models assessed on the testing set and the phenotype definition of either ICD or Self-Reported. The ICD or self-reported method is the same method used throughout the majority of analyses. Please note that values of 0 are in fact greater than 0 but round to 0 when only keeping values greater than $1 \times 10^{-4}$

| Method | AUC Mean | AUC SE | AUC Rank Mean | AUC Rank SE |
|---|---|---|---|---|
| SBayesR | 0.652 | 0.0147 | 8.09 | 0.586 |
| SMTpred | 0.623 | 0.0158 | 11.7 | 0.437 |
| Clump | 0.664 | 0.014 | 5.13 | 0.384 |
| DBLSMM | 0.646 | 0.0139 | 10.2 | 0.42 |
| Double-Weight | 0.628 | 0.0152 | 13.5 | 0.301 |
| LDpred | 0.655 | 0.015 | 6.22 | 0.749 |
| prsCS | 0.674 | 0.0143 | 2.22 | 0.208 |
| WC-Lasso | 0.654 | 0.0148 | 8.09 | 0.596 |
| WC-Likelihood | 0.627 | 0.0158 | 13.3 | 0.39 |
| Tweedie | 0.661 | 0.0145 | 6.17 | 0.293 |
| WC-2D | 0.664 | 0.0139 | 5.09 | 0.466 |
| SBLUP | 0.644 | 0.0148 | 9.43 | 0.376 |
| lassosum | 0.672 | 0.0146 | 2.7 | 0.455 |
| JAMPred | 0.627 | 0.0154 | 12.8 | 0.456 |
| LDpred2 | 0.668 | 0.0157 | 4.64 | 0.823 |

**Supplementary Table S32.** The mean AUC and rank as determined by the AUC across all diseases for each polygenic risk score generative method

| Method | Conc. Mean | Conc. SE | Conc. Rank Mean | Conc. Rank SE |
|---|---|---|---|---|
| SBayesR | 0.637 | 0.0151 | 7.74 | 0.459 |
| SMTpred | 0.606 | 0.0158 | 12 | 0.442 |
| Clump | 0.65 | 0.015 | 5.39 | 0.554 |
| DBLSMM | 0.631 | 0.0143 | 10.1 | 0.437 |
| Double-Weight | 0.613 | 0.015 | 13.3 | 0.5 |
| LDpred | 0.642 | 0.0149 | 6.35 | 0.699 |
| prsCS | 0.662 | 0.0146 | 2.09 | 0.188 |
| WC-Lasso | 0.64 | 0.0153 | 8.09 | 0.586 |
| WC-Likelihood | 0.612 | 0.0159 | 13.2 | 0.377 |
| Tweedie | 0.647 | 0.0153 | 6.43 | 0.411 |
| WC-2D | 0.651 | 0.0143 | 5.17 | 0.513 |
| SBLUP | 0.629 | 0.0151 | 9.22 | 0.44 |
| lassosum | 0.659 | 0.0152 | 2.74 | 0.463 |
| JAMPred | 0.612 | 0.0155 | 12.8 | 0.469 |
| LDpred2 | 0.654 | 0.0161 | 4.64 | 0.743 |

**Supplementary Table S33.** The mean concordance and rank as determined by the concordance across all diseases for each polygenic risk score generative method

| Method | OR Mean | OR SE | OR Rank Mean | OR Rank SE |
|---|---|---|---|---|
| SBayesR | 9.41 | 2.34 | 7.52 | 0.558 |
| SMTpred | 6.3 | 1.41 | 11.4 | 0.719 |
| Clump | 10 | 2.41 | 5.57 | 0.407 |
| DBLSMM | 8.72 | 2.21 | 9.17 | 0.585 |
| Double-Weight | 7.23 | 1.93 | 13 | 0.295 |
| LDpred | 9.87 | 2.52 | 5.83 | 0.836 |
| prsCS | 10.5 | 2.4 | 3 | 0.559 |
| WC-Lasso | 8.9 | 2.17 | 8.22 | 0.644 |
| WC-Likelihood | 7.69 | 2.06 | 13 | 0.557 |
| Tweedie | 9.72 | 2.23 | 5.83 | 0.375 |
| WC-2D | 9.51 | 2.13 | 6.39 | 0.612 |
| SBLUP | 8.78 | 2.32 | 8.91 | 0.632 |
| lassosum | 10.7 | 2.55 | 3.22 | 0.507 |
| JAMPred | 7.4 | 2.05 | 12.9 | 0.48 |
| LDpred2 | 10.4 | 2.41 | 5.36 | 0.887 |

**Supplementary Table S34.** The mean odds ratio and rank as determined by the odds ratio across all diseases for each polygenic risk score generative method

| Method | Cum. Hazard Mean | Cum. Hazard SE | Cum. Hazard Rank Mean | Cum. Hazard Rank SE |
|---|---|---|---|---|
| SBayesR | 0.0422 | 0.00857 | 7.96 | 0.553 |
| SMTpred | 0.0369 | 0.00829 | 12 | 0.393 |
| Clump | 0.0448 | 0.00896 | 5.22 | 0.479 |
| DBLSMM | 0.0404 | 0.00814 | 9.65 | 0.564 |
| Double-Weight | 0.0352 | 0.00744 | 13.3 | 0.438 |
| LDpred | 0.0431 | 0.00875 | 6.52 | 0.665 |
| prsCS | 0.0482 | 0.00963 | 1.96 | 0.194 |
| WC-Lasso | 0.0414 | 0.00798 | 7.78 | 0.647 |
| WC-Likelihood | 0.035 | 0.00742 | 13.1 | 0.402 |
| Tweedie | 0.0442 | 0.0088 | 6.48 | 0.43 |
| WC-2D | 0.0443 | 0.00887 | 5.7 | 0.46 |
| SBLUP | 0.0401 | 0.00826 | 9.3 | 0.424 |
| lassosum | 0.0487 | 0.00973 | 2.43 | 0.426 |
| JAMPred | 0.0362 | 0.00761 | 12.9 | 0.457 |
| LDpred2 | 0.0467 | 0.0096 | 4.95 | 0.763 |

**Supplementary Table S35.** The mean cumulative hazard and rank as determined by the cumulative across all diseases for each polygenic risk score generative method

| Attribute | SMTpred | Clump | DBLSMM | SBayesR | Double-Weight | LDpred |
|---|---|---|---|---|---|---|
| PRS SNPs | 0 | 0 | 0 | 1 | 0 | 0 |
| PRS SNPs Rev. | 0 | 1 | 0 | 0 | 0 | 2 |
| PRS Dist. by Effect | 1 | 3 | 2 | 1 | 1 | 0 |
| PRS Dist. by Effect Rev. | 1 | 3 | 2 | 1 | 1 | 0 |
| PRS Dist. by Length | 1 | 3 | 2 | 1 | 1 | 0 |
| PRS Dist. by Length Rev. | 1 | 3 | 2 | 1 | 1 | 0 |
| GWAS SNPs <1e-6 | 0 | 0 | 0 | 0 | 0 | 0 |
| GWAS SNPs <1e-6 Rev. | 0 | 0 | 0 | 0 | 0 | 1 |
| GWAS SNPs <1e-8 | 0 | 0 | 0 | 0 | 0 | 0 |
| GWAS SNPs <1e-8 Rev. | 0 | 0 | 0 | 0 | 0 | 1 |
| GWAS Dist. by Length | 0 | 0 | 0 | 0 | 0 | 2 |
| GWAS Dist. by Length Rev. | 0 | 0 | 0 | 0 | 0 | 0 |
| GWAS Dist. by Effect | 0 | 0 | 0 | 0 | 0 | 1 |
| GWAS Dist. by Effect Rev. | 0 | 0 | 0 | 0 | 0 | 1 |
| GWAS Sample Size | 0 | 0 | 0 | 0 | 0 | 0 |
| GWAS Sample Size Rev. | 0 | 0 | 0 | 0 | 0 | 1 |
| GWAS SNPs | 0 | 1 | 0 | 0 | 0 | 1 |
| GWAS SNPs Rev. | 0 | 0 | 0 | 0 | 0 | 1 |
| GWAS Heritability | 0 | 1 | 0 | 0 | 0 | 1 |
| GWAS Heritability Rev. | 0 | 0 | 0 | 0 | 0 | 0 |
| UKBB CC Ratio | 0 | 0 | 0 | 0 | 0 | 0 |
| UKBB CC Ratio Rev. | 0 | 0 | 0 | 0 | 0 | 1 |
| GWAS CC Ratio | 0 | 0 | 0 | 0 | 0 | 2 |
| GWAS CC Ratio Rev. | 0 | 0 | 0 | 0 | 0 | 1 |

**Supplementary Table S36.** The count of methods amongst the top ten AUCs weighted by the attribute listed. The best AUC for each disease and method combination was utilized for weighting and ranking.

| Attribute | prsCS | WC-Lasso | WC-Like | Tweedie | WC-2D | SBLUP | lassosum | JAMPred | LDpred2 |
|---|---|---|---|---|---|---|---|---|---|
| PRS SNPs | 1 | 2 | 0 | 0 | 0 | 0 | 3 | 2 | 1 |
| PRS SNPs Rev. | 1 | 0 | 0 | 0 | 1 | 0 | 3 | 0 | 2 |
| PRS Dist. by Effect | 0 | 1 | 0 | 0 | 0 | 1 | 0 | 0 | 0 |
| PRS Dist. by Effect Rev. | 0 | 1 | 0 | 0 | 0 | 1 | 0 | 0 | 0 |
| PRS Dist. by Length | 0 | 1 | 0 | 0 | 0 | 1 | 0 | 0 | 0 |
| PRS Dist. by Length Rev. | 0 | 1 | 0 | 0 | 0 | 1 | 0 | 0 | 0 |
| GWAS SNPs <1e-6 | 3 | 0 | 0 | 0 | 1 | 0 | 4 | 0 | 2 |
| GWAS SNPs <1e-6 Rev. | 3 | 0 | 0 | 0 | 1 | 0 | 4 | 0 | 1 |
| GWAS SNPs <1e-8 | 3 | 0 | 0 | 0 | 1 | 0 | 4 | 0 | 2 |
| GWAS SNPs <1e-8 Rev. | 3 | 0 | 0 | 0 | 1 | 0 | 4 | 0 | 1 |
| GWAS Dist. by Length | 2 | 0 | 0 | 0 | 1 | 0 | 4 | 0 | 1 |
| GWAS Dist. by Length Rev. | 4 | 0 | 0 | 0 | 0 | 0 | 5 | 0 | 1 |
| GWAS Dist. by Effect | 4 | 0 | 0 | 0 | 0 | 0 | 4 | 0 | 1 |
| GWAS Dist. by Effect Rev. | 2 | 0 | 0 | 0 | 1 | 0 | 5 | 0 | 1 |
| GWAS Sample Size | 4 | 0 | 0 | 0 | 0 | 0 | 6 | 0 | 0 |
| GWAS Sample Size Rev. | 2 | 0 | 0 | 0 | 1 | 0 | 4 | 0 | 2 |
| GWAS SNPs | 3 | 0 | 0 | 0 | 1 | 0 | 4 | 0 | 0 |
| GWAS SNPs Rev. | 2 | 0 | 0 | 0 | 0 | 0 | 5 | 0 | 2 |
| GWAS Heritability | 2 | 0 | 0 | 0 | 1 | 0 | 4 | 0 | 1 |
| GWAS Heritability Rev. | 3 | 0 | 0 | 0 | 0 | 0 | 5 | 0 | 2 |
| UKBB CC Ratio | 3 | 0 | 0 | 0 | 0 | 0 | 7 | 0 | 0 |
| UKBB CC Ratio Rev. | 2 | 0 | 0 | 0 | 1 | 0 | 5 | 0 | 1 |
| GWAS CC Ratio | 2 | 0 | 0 | 0 | 1 | 0 | 4 | 0 | 1 |
| GWAS CC Ratio Rev. | 3 | 0 | 0 | 0 | 1 | 0 | 4 | 0 | 1 |

**Supplementary Table S37.** Continued from above. The count of methods amongst the top ten AUCs weighted by the attribute listed. The best AUC for each disease and method combination was utilized for weighting and ranking.

| Disease | Variants in Best PRS |
| --- | --- |
| Lupus | 16205 |
| A. Fib. | 987016 |
| Asthma | 69914 |
| Celiac Disease | 494401 |
| Migraine | 1074509 |
| MS | 16177 |
| Vitiligo | 403 |
| Gout | 21 |
| Crohns Disease | 20553 |
| Ulcerative Colitis | 45734 |
| Type 2 Diabetes | 4422 |
| Stroke | 162615 |
| Breast Cancer | 24433 |
| NAFLD | 403057 |
| CAD | 1079978 |
| Rheumatoid Arthritis | 396743 |
| Type 1 Diabetes | 44166 |
| Ovarian Cancer | 6129416 |
| Prostate Cancer | 15901 |
| Heart Failure | 548929 |
| Psoriasis | 8361 |
| Depression | 514903 |

**Supplementary Table S38.** The number of variants in each polygenic risk score used in the testing phase.

| Disease | Q1 | Q2 | Q3 | Q4 |
|---|---|---|---|---|
| Lupus | 0.108 | 0.164 | 0.234 | 0.494 |
| A. Fib. | 0.0187 | 0.0846 | 0.203 | 0.694 |
| Asthma | 0.0571 | 0.173 | 0.28 | 0.491 |
| Celiac Disease | 0.00016 | 0.00853 | 0.122 | 0.869 |
| Migraine | 0.0329 | 0.117 | 0.233 | 0.617 |
| MS | 0.00528 | 0.0484 | 0.22 | 0.726 |
| Vitiligo | 0.126 | 0.199 | 0.291 | 0.385 |
| Gout | 0.0602 | 0.119 | 0.401 | 0.419 |
| Crohns Disease | 0.0521 | 0.161 | 0.269 | 0.518 |
| Ulcerative Colitis | 0.00389 | 0.0553 | 0.218 | 0.723 |
| Type 2 Diabetes | 0.0554 | 0.167 | 0.265 | 0.513 |
| Stroke | 0.0557 | 0.172 | 0.281 | 0.492 |
| Breast Cancer | 0.0432 | 0.15 | 0.259 | 0.548 |
| NAFLD | 1.78e-06 | 0.000171 | 0.0145 | 0.985 |
| CAD | 0.0326 | 0.115 | 0.228 | 0.624 |
| Rheumatoid Arthritis | 0.00887 | 0.0319 | 0.0767 | 0.883 |
| Type 1 Diabetes | 0.0048 | 0.0235 | 0.0886 | 0.883 |
| Ovarian Cancer | 7.89e-07 | 0.00107 | 0.072 | 0.927 |
| ALS | 0.0131 | 0.101 | 0.218 | 0.669 |
| Prostate Cancer | 0.0397 | 0.136 | 0.244 | 0.58 |
| Heart Failure | 0.0519 | 0.167 | 0.28 | 0.501 |
| Psoriasis | 0.0289 | 0.0931 | 0.185 | 0.693 |
| Depression | 0.0454 | 0.139 | 0.247 | 0.569 |

**Supplementary Table S39.** For each disease the variants of the polygenic risk score used in the testing phase were ordered by absolute effect, and split into four groups such that the sum of effects in each group were equal. The size of each group, standardized such that the minimum was zero and maximum was one, were calculated.

| Disease | Coding | UTR-5 | Cardiovascular Cell Type | Adrenal/Pancreas Cell Type | H3K4me1 Peaks | Flanked Bivalent TSS | Ancient Human Enhancer |
|---|---|---|---|---|---|---|---|
| Lupus | 0.0323 | 0.00564 | 0.0107 | 0.0141 | 0.0176 | 0.0215 | 0.0472 |
| A. Fib. | 0.0452 | 0.0346 | 0.028 | 0.0214 | 0.0193 | 0.0253 | 0.0156 |
| Asthma | 0.0307 | 0.0241 | 0.0178 | 0.0223 | 0.0167 | 0.0421 | 0.0362 |
| Celiac Disease | 0.0339 | 0.0539 | 0.0157 | 0.0136 | 0.0136 | 0.0161 | 0.0199 |
| Migraine | 0.0445 | 0.0316 | 0.0177 | 0.0238 | 0.0137 | 0.0169 | 0.0296 |
| MS | 0.0213 | 0.0319 | 0.0176 | 0.0146 | 0.0167 | 0.0169 | 0.0401 |
| Vitiligo | 0.0284 | 0.0189 | 0.0188 | 0.0237 | 0.0235 | 0.0304 | 0.033 |
| Gout | 0.115 | 0 | 0.00944 | 0 | 0.0104 | 0 | 0 |
| Crohns Disease | 0.0445 | 0.0284 | 0.0152 | 0.0177 | 0.0116 | 0.0277 | 0.0402 |
| Ulcerative Colitis | 0.0282 | 0.0199 | 0.0131 | 0.0151 | 0.0154 | 0.0152 | 0.0503 |
| Type 2 Diabetes | 0.0149 | 0.015 | 0.0264 | 0.032 | 0.0255 | 0.0272 | 0.0342 |
| Stroke | 0.0275 | 0.0216 | 0.0263 | 0.0257 | 0.0187 | 0.0452 | 0.0404 |
| Breast Cancer | 0.0296 | 0.0327 | 0.0215 | 0.0226 | 0.0175 | 0.04 | 0.0517 |
| NAFLD | 0.15 | 0.00206 | 0.0049 | 0.00682 | 0.0275 | 0.00414 | 0.0204 |
| CAD | 0.0439 | 0.028 | 0.0196 | 0.0226 | 0.0154 | 0.0207 | 0.0536 |
| Rheumatoid Arthritis | 0.0448 | 0.0468 | 0.00453 | 0.011 | 0.00978 | 0.0471 | 0.0467 |
| Type 1 Diabetes | 0.0589 | 0.0131 | 0.00918 | 0.0301 | 0.0219 | 0.0537 | 0.00826 |
| Ovarian Cancer | 0.00354 | 0.0123 | 0.0085 | 0.015 | 0.0126 | 0.0808 | 0.00329 |
| ALS | 0.0382 | 0.0337 | 0.0164 | 0.024 | 0.0187 | 0.0265 | 0.0258 |
| Prostate Cancer | 0.0342 | 0.0319 | 0.0164 | 0.0211 | 0.013 | 0.0395 | 0.0302 |
| Heart Failure | 0.0384 | 0.0237 | 0.0225 | 0.0248 | 0.0162 | 0.0551 | 0.0387 |
| Psoriasis | 0.0325 | 0.027 | 0.022 | 0.0252 | 0.0209 | 0.0286 | 0.016 |
| Depression | 0.0398 | 0.0131 | 0.0212 | 0.0225 | 0.0226 | 0.0643 | 0.0381 |

**Supplementary Table S40.** The functional annotation scores. Computed for each disease, with the variants of the polygenic risk score used in the testing phase identified as belonging to any of 44 functional annotation groups. The mean of the absolute effects of the variants in each group were calculated, and normalized such that the sum of resulting functional annotation weights for each disease summed to one.

| Disease | H3K27ac Peaks | Ancient Human Promoter | Skeletal/Muscle Cell Type | Human Promoter | H3K4me3 Peaks | Repressed | Intron |
|---|---|---|---|---|---|---|---|
| Lupus | 0.0193 | 0.0619 | 0.0139 | 0.031 | 0.0286 | 0.0232 | 0.0215 |
| A. Fib. | 0.0191 | 0.028 | 0.0248 | 0.0145 | 0.0206 | 0.00658 | 0.0178 |
| Asthma | 0.0173 | 0.0465 | 0.0233 | 0.0362 | 0.0252 | 0 | 0.00764 |
| Celiac Disease | 0.0153 | 0.0789 | 0.0166 | 0.0573 | 0.0183 | 0 | 0.0102 |
| Migraine | 0.0129 | 0.0892 | 0.02 | 0.0447 | 0.025 | 0 | 0.0109 |
| MS | 0.0179 | 0.0551 | 0.021 | 0.0408 | 0.0342 | 0 | 0.0117 |
| Vitiligo | 0.0251 | 0.0172 | 0.0202 | 0.0241 | 0.025 | 0.0334 | 0.0221 |
| Gout | 0.0167 | 0 | 0.0137 | 0 | 0.0544 | 0.0056 | 0.0391 |
| Crohns Disease | 0.0106 | 0.0365 | 0.0206 | 0.0214 | 0.0192 | 0 | 0.000691 |
| Ulcerative Colitis | 0.0177 | 0.0467 | 0.0181 | 0.0239 | 0.0294 | 0 | 0.00837 |
| Type 2 Diabetes | 0.0216 | 0.0269 | 0.0229 | 0.0316 | 0.0148 | 0 | 0.00765 |
| Stroke | 0.018 | 0.0456 | 0.0288 | 0.0298 | 0.022 | 0 | 0.00841 |
| Breast Cancer | 0.0167 | 0.0475 | 0.024 | 0.0282 | 0.0218 | 0 | 0.00767 |
| NAFLD | 0.0162 | 0.00567 | 0.0044 | 0.00214 | 0.00186 | 0.00423 | 0.00333 |
| CAD | 0.0159 | 0.0348 | 0.0213 | 0.0354 | 0.0247 | 0 | 0.0106 |
| Rheumatoid Arthritis | 0.00984 | 0.0938 | 0.01 | 0.0538 | 0.022 | 0.0011 | 0.0014 |
| Type 1 Diabetes | 0.02 | 0.005 | 0.0151 | 0.013 | 0.0283 | 0 | 0.00966 |
| Ovarian Cancer | 0.0145 | 0.0631 | 0.0167 | 0.0207 | 0.0264 | 0.00274 | 0.0145 |
| ALS | 0.0157 | 0.0526 | 0.0205 | 0.0378 | 0.0174 | 0 | 0.0172 |
| Prostate Cancer | 0.0144 | 0.0803 | 0.0184 | 0.0455 | 0.0186 | 0 | 0.00603 |
| Heart Failure | 0.0187 | 0.0409 | 0.0279 | 0.0279 | 0.0204 | 0 | 0.00736 |
| Psoriasis | 0.0195 | 0.0528 | 0.0198 | 0.0373 | 0.0283 | 0.0273 | 0.0191 |
| Depression | 0.0177 | 0.0405 | 0.0219 | 0.00992 | 0.0277 | 0.00877 | 0.00626 |

**Supplementary Table S41.** Functional annotation weights, continued

| Disease | Dnase I Hyper Peak | Human Enhancer | Promoter-ExAC | UTR-3 | Enhancer-1 | H3K27ac | Transcribed |
|---|---|---|---|---|---|---|---|
| Lupus | 0.0122 | 0.013 | 0.0159 | 0.0161 | 0.0449 | 0.0211 | 0.0232 |
| A. Fib. | 0.0182 | 0.0172 | 0.0396 | 0.0221 | 0 | 0.0188 | 0.0208 |
| Asthma | 0.015 | 0.0331 | 0.0221 | 0.0301 | 0.0379 | 0.0193 | 0.00796 |
| Celiac Disease | 0.0155 | 0.0282 | 0.0148 | 0.021 | 0.0545 | 0.0189 | 0.0114 |
| Migraine | 0.0119 | 0.0141 | 0.0756 | 0.0321 | 0.0125 | 0.0149 | 0.0102 |
| MS | 0.0118 | 0.036 | 0.0164 | 0.0104 | 0.0249 | 0.0206 | 0.014 |
| Vitiligo | 0.0205 | 0.014 | NA | 0.0142 | 0 | 0.0201 | 0.026 |
| Gout | 0.0335 | 0 | 0 | 0 | 0 | 0.0162 | 0.0461 |
| Crohns Disease | 0.0199 | 0.0255 | 0.0519 | 0.0235 | 0.0943 | 0.0126 | 0.00462 |
| Ulcerative Colitis | 0.0197 | 0.0393 | 0.0292 | 0.0204 | 0.0738 | 0.0172 | 0.0129 |
| Type 2 Diabetes | 0.019 | 0.0247 | 0.0607 | 0.0225 | NA | 0.0255 | 0.0207 |
| Stroke | 0.0171 | 0.0273 | 0.0338 | 0.0238 | 0.0271 | 0.0194 | 0.00428 |
| Breast Cancer | 0.016 | 0.0322 | 0.0424 | 0.0173 | 0.0263 | 0.018 | 0.00702 |
| NAFLD | 0.00451 | 0.0107 | 0.0107 | 0.228 | 0.00087 | 0.0211 | 0.0169 |
| CAD | 0.0154 | 0.0297 | 0.0296 | 0.036 | 0.0367 | 0.0182 | 0.0151 |
| Rheumatoid Arthritis | 0.00934 | 0.0277 | 0.0309 | 0.0295 | 0.0547 | 0.0124 | 0 |
| Type 1 Diabetes | 0.0104 | 0.023 | 0.0369 | 0.0331 | 0.0113 | 0.0255 | 0.0112 |
| Ovarian Cancer | 0.0208 | 0.00408 | 0.0713 | 0 | 0.00675 | 0.0116 | 0.0112 |
| ALS | 0.0134 | 0.0146 | 0.0593 | 0.0356 | 0.00935 | 0.0147 | 0.0187 |
| Prostate Cancer | 0.0129 | 0.0195 | 0.0889 | 0.0246 | 0.0263 | 0.0153 | 0.00799 |
| Heart Failure | 0.014 | 0.0283 | 0.0227 | 0.0233 | 0.0273 | 0.0186 | 0.00238 |
| Psoriasis | 0.0193 | 0.016 | 0 | 0.0212 | 0.0229 | 0.019 | 0.0186 |
| Depression | 0.018 | 0.0189 | 0.0219 | 0.0336 | 0.0165 | 0.019 | 0 |

**Supplementary Table S42.** Functional annotation weights, continued

| Disease | Conserved Vertebrate | Dnase I Hyper | Promoter Flanking | Super Enhancer | Dnase I Hypersensitive-Fetal | Other Cell Type | Weak Enhancer |
|---|---|---|---|---|---|---|---|
| Lupus | 0.0302 | 0.0136 | 0 | 0.0248 | 0.014 | 0.0185 | 0.0073 |
| A. Fib. | 0.0372 | 0.0163 | 0.0261 | 0.0207 | 0.0197 | 0.0207 | 0.02 |
| Asthma | 0.0123 | 0.0136 | 0.0227 | 0.0275 | 0.0154 | 0.0188 | 0.0258 |
| Celiac Disease | 0.00421 | 0.0137 | 0.0455 | 0.0287 | 0.0175 | 0.0139 | 0.00966 |
| Migraine | 0.0207 | 0.0125 | 0.0114 | 0.0169 | 0.0151 | 0.0143 | 0.0135 |
| MS | 0.00989 | 0.0134 | 0.0267 | 0.0292 | 0.0158 | 0.0185 | 0.00919 |
| Vitiligo | 0.0269 | 0.0224 | 0.0492 | 0.0204 | 0.0189 | 0.018 | 0.0319 |
| Gout | 0.055 | 0.0244 | 0 | 0 | 0.0131 | 0.00871 | 0.0357 |
| Crohns Disease | 0.0324 | 0.0162 | 0.000153 | 0.0133 | 0.0185 | 0.0171 | 0.0262 |
| Ulcerative Colitis | 0.0265 | 0.0178 | 0.00652 | 0.0205 | 0.0214 | 0.0179 | 0.0242 |
| Type 2 Diabetes | 0.0198 | 0.0176 | 0.0127 | 0.0279 | 0.0168 | 0.0205 | 0.0327 |
| Stroke | 0.0179 | 0.0155 | 0.00886 | 0.0279 | 0.0195 | 0.0208 | 0.03 |
| Breast Cancer | 0.019 | 0.0151 | 0.0123 | 0.0267 | 0.0163 | 0.0195 | 0.0194 |
| NAFLD | 0.00278 | 0.0262 | 0.0192 | 0.00588 | 0.00485 | 0.0256 | 0.00303 |
| CAD | 0.0248 | 0.0145 | 0.0135 | 0.0214 | 0.016 | 0.019 | 0.0192 |
| Rheumatoid Arthritis | 0.0103 | 0.00736 | 0.016 | 0.0161 | 0.0114 | 0.0138 | 0.0174 |
| Type 1 Diabetes | 0.026 | 0.0115 | 0.0322 | 0.0393 | 0.00612 | 0.0242 | 0.0104 |
| Ovarian Cancer | 0.023 | 0.0152 | 0.00823 | 0.0108 | 0.0219 | 0.0118 | 0.0163 |
| ALS | 0.0272 | 0.0125 | 0.0216 | 0.0186 | 0.0139 | 0.0178 | 0.0202 |
| Prostate Cancer | 0.0184 | 0.0122 | 0.0185 | 0.0197 | 0.0168 | 0.0153 | 0.0129 |
| Heart Failure | 0.0251 | 0.0118 | 0.00537 | 0.0294 | 0.0176 | 0.0183 | 0.0258 |
| Psoriasis | 0.0196 | 0.0197 | 0.0229 | 0.0163 | 0.0204 | 0.0205 | 0.02 |
| Depression | 0.0371 | 0.0184 | 0.0194 | 0.0175 | 0.0203 | 0.0175 | 0.0242 |

**Supplementary Table S43.** Functional annotation weights, continued

| Disease | Promoter | Liver Cell Type | Conserved Primate | Conserved | H3K9ac Peaks | TSS | Kidney Cell Type |
|---|---|---|---|---|---|---|---|
| Lupus | 0.0346 | 0.0131 | 0.0274 | 0.0371 | 0.0173 | 0.0583 | 0.012 |
| A. Fib. | 0.0194 | 0.0209 | 0.0469 | 0.0426 | 0.0226 | 0.019 | 0.0167 |
| Asthma | 0.0275 | 0.0236 | 0.0151 | 0.0141 | 0.0267 | 0.0429 | 0.0238 |
| Celiac Disease | 0.0191 | 0.0221 | 0.0107 | 0.00537 | 0.0219 | 0.0739 | 0.0324 |
| Migraine | 0.0183 | 0.0185 | 0.0327 | 0.0284 | 0.0206 | 0.0358 | 0.0237 |
| MS | 0.0368 | 0.0229 | 0.014 | 0.0123 | 0.0397 | 0.0631 | 0.0307 |
| Vitiligo | 0.0241 | 0.0105 | 0.0359 | 0.0294 | 0.0245 | 0.0245 | 0.018 |
| Gout | 0.115 | 0.00389 | 0.0742 | 0.0576 | 0.0119 | 0 | 0.0471 |
| Crohns Disease | 0.0174 | 0.0197 | 0.0333 | 0.0383 | 0.0197 | 0.0243 | 0.021 |
| Ulcerative Colitis | 0.0209 | 0.0215 | 0.0261 | 0.0258 | 0.0228 | 0.0299 | 0.0226 |
| Type 2 Diabetes | 0.0115 | 0.0296 | 0.0209 | 0.0191 | 0.0272 | 0.03 | 0.0259 |
| Stroke | 0.0182 | 0.0231 | 0.0241 | 0.0233 | 0.0259 | 0.0259 | 0.031 |
| Breast Cancer | 0.0179 | 0.0234 | 0.0247 | 0.0187 | 0.0269 | 0.0319 | 0.0313 |
| NAFLD | 0.00164 | 0.0677 | 0.00305 | 0.1 | 0.00414 | 0.00258 | 0.11 |
| CAD | 0.0177 | 0.0274 | 0.0252 | 0.0237 | 0.0242 | 0.0308 | 0.0255 |
| Rheumatoid Arthritis | 0.0463 | 0.0155 | 0.0103 | 0.00662 | 0.0201 | 0.0709 | 0.0262 |
| Type 1 Diabetes | 0.0614 | 0.0291 | 0.038 | 0.0296 | 0.016 | 0.0288 | 0.0151 |
| Ovarian Cancer | 0.0432 | 0.0231 | 0.0277 | 0.0236 | 0.0196 | 0.0684 | 0.0282 |
| ALS | 0.0253 | 0.0191 | 0.0362 | 0.032 | 0.0284 | 0.022 | 0.0273 |
| Prostate Cancer | 0.0198 | 0.0232 | 0.0258 | 0.0212 | 0.0201 | 0.0344 | 0.0285 |
| Heart Failure | 0.024 | 0.0183 | 0.0327 | 0.0256 | 0.0263 | 0.0344 | 0.0251 |
| Psoriasis | 0.036 | 0.0219 | 0.0208 | 0.0186 | 0.0252 | 0.0371 | 0.0295 |
| Depression | 0.00506 | 0.0102 | 0.0392 | 0.0392 | 0.0286 | 0.0226 | 0.0161 |

**Supplementary Table S44.** Functional annotation weights, continued

| Disease | Hematopoetic Cell Type | TFBS | Enhancer-2 | Dispersed Gene Family | GI Cell Type | Conserved Mammal |
|---|---|---|---|---|---|---|
| Lupus | 0.0245 | 0.0252 | 0.0162 | 0.0207 | 0.0167 | 0.0364 |
| A. Fib. | 0.017 | 0.0196 | 0.0206 | 0.0138 | 0.0219 | 0.0402 |
| Asthma | 0.0227 | 0.0197 | 0.0313 | 0.0178 | 0.021 | 0.0139 |
| Celiac Disease | 0.0225 | 0.0222 | 0.0269 | 0.0205 | 0.0181 | 0.00534 |
| Migraine | 0.0126 | 0.016 | 0.019 | 0.0148 | 0.0162 | 0.0274 |
| MS | 0.0295 | 0.0251 | 0.0201 | 0.0161 | 0.0217 | 0.0177 |
| Vitiligo | 0.0237 | 0.0233 | 0.0156 | 0.025 | 0.0156 | 0.0258 |
| Gout | 0.0193 | 0.0128 | 0 | 0.0276 | 0.00981 | 0.0657 |
| Crohns Disease | 0.0122 | 0.0203 | 0.0257 | 0.0177 | 0.0187 | 0.0374 |
| Ulcerative Colitis | 0.0196 | 0.0232 | 0.0311 | 0.0183 | 0.0238 | 0.0277 |
| Type 2 Diabetes | 0.0224 | 0.026 | 0.033 | 0.0201 | 0.0265 | 0.0213 |
| Stroke | 0.0161 | 0.0176 | 0.0254 | 0.0164 | 0.0229 | 0.0224 |
| Breast Cancer | 0.0164 | 0.0203 | 0.0244 | 0.0166 | 0.0221 | 0.0211 |
| NAFLD | 0.0254 | 0.00341 | 0.00471 | 0.00367 | 0.00563 | 0.00101 |
| CAD | 0.0181 | 0.0153 | 0.0197 | 0.0156 | 0.0214 | 0.0266 |
| Rheumatoid Arthritis | 0.0199 | 0.0212 | 0.0226 | 0.0138 | 0.0144 | 0.0121 |
| Type 1 Diabetes | 0.0319 | 0.0209 | 0.0311 | 0.0269 | 0.0185 | 0.0341 |
| Ovarian Cancer | 0.0103 | 0.0223 | 0.0206 | 0.034 | 0.0176 | 0.0249 |
| ALS | 0.0149 | 0.0145 | 0.0155 | 0.0149 | 0.018 | 0.034 |
| Prostate Cancer | 0.0146 | 0.0152 | 0.0247 | 0.0123 | 0.017 | 0.02 |
| Heart Failure | 0.0172 | 0.0165 | 0.0257 | 0.0152 | 0.0209 | 0.0261 |
| Psoriasis | 0.0172 | 0.0212 | 0.0188 | 0.0212 | 0.0213 | 0.0206 |
| Depression | 0.0161 | 0.0204 | 0.0245 | 0.0152 | 0.0207 | 0.0399 |

**Supplementary Table S45.** Functional annotation weights, continued

| Disease | Connective Bone Cell Type | CNS Cell Type | CTCF |
|---|---|---|---|
| Lupus | 0.0202 | 0.0132 | 0.0237 |
| A. Fib. | 0.0223 | 0.0219 | 0.0164 |
| Asthma | 0.02 | 0.0174 | 0.0151 |
| Celiac Disease | 0.0187 | 0.0115 | 0.00802 |
| Migraine | 0.0209 | 0.019 | 0.0198 |
| MS | 0.023 | 0.0166 | 0.0141 |
| Vitiligo | 0.0257 | 0.0313 | 0.0207 |
| Gout | 0 | 0.0175 | 0.041 |
| Crohns Disease | 0.0171 | 0.0111 | 0.0155 |
| Ulcerative Colitis | 0.0168 | 0.0141 | 0.00706 |
| Type 2 Diabetes | 0.0293 | 0.0229 | 0.0125 |
| Stroke | 0.0217 | 0.0202 | 0.0147 |
| Breast Cancer | 0.0244 | 0.0171 | 0.0137 |
| NAFLD | 0.0015 | 0.0317 | 0 |
| CAD | 0.0237 | 0.0198 | 0.0101 |
| Rheumatoid Arthritis | 0.0169 | 0.00924 | 0.0146 |
| Type 1 Diabetes | 0.0139 | 0.0141 | 0.0031 |
| Ovarian Cancer | 0.0115 | 0.0104 | 0.0873 |
| ALS | 0.0192 | 0.0267 | 0.0102 |
| Prostate Cancer | 0.0188 | 0.0142 | 0.0114 |
| Heart Failure | 0.0186 | 0.0203 | 0.0207 |
| Psoriasis | 0.0231 | 0.0209 | 0.014 |
| Depression | 0.0208 | 0.0203 | 0.0244 |

**Supplementary Table S46.** Functional annotation weights, continued

| Disease | SIFT | Polyphen2-HDIV | Polyphen2-HVAR | LRT |
|---|---|---|---|---|
| Lupus | 0.114 | 0.103 | 0.0951 | 0.0926 |
| A. Fib. | 0.122 | 0.0986 | 0.09 | 0.0932 |
| Asthma | 0.116 | 0.102 | 0.094 | 0.0943 |
| Celiac Disease | 0.163 | 0.071 | 0.0384 | 0.0287 |
| Migraine | 0.116 | 0.102 | 0.0941 | 0.0934 |
| MS | 0.117 | 0.0975 | 0.0934 | 0.108 |
| Vitiligo | 0.0979 | 0.107 | 0.107 | 0.0958 |
| Gout | 0.113 | 0.121 | 0.109 | 0.0962 |
| Crohns Disease | 0.127 | 0.0947 | 0.087 | 0.0879 |
| Ulcerative Colitis | 0.117 | 0.109 | 0.104 | 0.0978 |
| Type 2 Diabetes | 0.115 | 0.108 | 0.1 | 0.0972 |
| Stroke | 0.117 | 0.107 | 0.101 | 0.0998 |
| Breast Cancer | 0.109 | 0.12 | 0.104 | 0.0912 |
| NAFLD | 0.114 | 0.0902 | 0.0825 | 0.09 |
| CAD | 0.115 | 0.109 | 0.102 | 0.0959 |
| Rheumatoid Arthritis | 0.114 | 0.0982 | 0.091 | 0.0869 |
| Type 1 Diabetes | 0.107 | 0.0287 | 0.0177 | 0.0407 |
| Ovarian Cancer | 0.101 | 0.0978 | 0.0924 | 0.1 |
| ALS | 0.109 | 0.107 | 0.1 | 0.0956 |
| Prostate Cancer | 0.115 | 0.11 | 0.101 | 0.0976 |
| Heart Failure | 0.109 | 0.109 | 0.103 | 0.0968 |
| Psoriasis | 0.104 | 0.101 | 0.0944 | 0.0961 |
| Depression | 0.115 | 0.103 | 0.0958 | 0.095 |

**Supplementary Table S47.** The deleterious weights. Computed for each disease, with the variants of the polygenic risk score used in the testing phase identified aligned to deleterious scores computed for 9 different methods. The sum of the product between each variant's absolute effect and the deleterious score of were calculated, divided by the total variants aligned to each deleterious score, and then normalized such that the sum of resulting deleterious weights for each disease summed to one.

| Disease | MutationTaster | FATHMM | PROVEAN | VEST4 | MVP |
|---|---|---|---|---|---|
| Lupus | 0.154 | 0.193 | 0 | 0.0886 | 0.0794 |
| A. Fib. | 0.172 | 0.19 | 0 | 0.0851 | 0.074 |
| Asthma | 0.157 | 0.189 | 0 | 0.0886 | 0.0789 |
| Celiac Disease | 0.234 | 0.211 | 0.227 | 0.0252 | 0 |
| Migraine | 0.164 | 0.184 | 0 | 0.0885 | 0.0783 |
| MS | 0.166 | 0.16 | 0 | 0.0934 | 0.0811 |
| Vitiligo | 0.151 | 0.154 | 0 | 0.108 | 0.0879 |
| Gout | 0.098 | 0.165 | 0 | 0.103 | 0.0962 |
| Crohns Disease | 0.184 | 0.207 | 0 | 0.0802 | 0.0659 |
| Ulcerative Colitis | 0.146 | 0.153 | 0 | 0.0968 | 0.0877 |
| Type 2 Diabetes | 0.156 | 0.156 | 0 | 0.0958 | 0.0859 |
| Stroke | 0.164 | 0.144 | 0 | 0.0966 | 0.0848 |
| Breast Cancer | 0.159 | 0.169 | 0 | 0.0917 | 0.0785 |
| NAFLD | 0.18 | 0.229 | 0 | 0.0797 | 0.067 |
| CAD | 0.156 | 0.162 | 0 | 0.093 | 0.0834 |
| Rheumatoid Arthritis | 0.161 | 0.218 | 0 | 0.0847 | 0.0723 |
| Type 1 Diabetes | 0.198 | 0.471 | 0.108 | 0.0262 | 0 |
| Ovarian Cancer | 0.213 | 0.123 | 0 | 0.102 | 0.0863 |
| ALS | 0.157 | 0.164 | 0 | 0.0955 | 0.086 |
| Prostate Cancer | 0.146 | 0.15 | 0 | 0.101 | 0.0913 |
| Heart Failure | 0.156 | 0.151 | 0 | 0.0986 | 0.0894 |
| Psoriasis | 0.145 | 0.199 | 0 | 0.0924 | 0.085 |
| Depression | 0.162 | 0.177 | 0 | 0.09 | 0.0807 |

**Supplementary Table S48.** Deleterious weights continued
