## Supplemental II: Methods for "A systematic framework for assessing the clinical impact of polygenic risk scores"

| Abbreviation | Meaning |
|---|---|
| MS | Multiple Sclerosis |
| NAFLD | Non-Alcoholic Fatty Liver Disease |
| CAD | Coronary Artery Disease |
| A. Fib. | Atrial Fibrillation |
| ALS | Amyotrophic Lateral Sclerosis |
| AUC | Area Under the Reciever Operator Curve |
| PRS | Polygenic Risk Score |
| Imp. | Improvement |
| PRS | Polygenic Risk Score |
| AUC | Area under the Receiver Operator Curve |
| NRI | Net Reclassification Index |
| Diff. | Different |

**Table M1.** Abbreviations used throughout this investigation

### Overview

The detailed analysis of polygenic risk scores began with the acquisition of GWAS summary statistics and supplementary data, involved the adjustment of summary statistics through polygenic risk score generative method and scoring of those statistics, and required diverse analyses of polygenic risk score predictive performance and validity. A detailed reporting of all data required, scripts written, and plots generated is available at https://kulmsc.github.io/pgs_book/index.html.

### Data Preparation

#### Acquisition of Summary Statistics

The majority of summary statistics were acquired from the GWAS Catalog (https://www.ebi.ac.uk/gwas/downloads/summary-statistics). All studies were sought that had relatively high sample size, studied relatively prevalent, binary, disease traits, contained both minor and major alleles, and did not use UK Biobank data. In total 21 traits were chosen. All summary statistics were downloaded directly from the FTP server. Two additional summary statistics were acquired that were not within the GWAS Catalog. Migraine data from Gormley et al. came from a 23andMe data agreement, and Multiple Sclerosis data from the Interntional Multiple Sclerosis Genetics Consortium came from their own website.

A conversion script was deployed to regularize all of the various summary statistics. To only retain the highest quality single nucleotide polymorphisms (SNPs), a set of stringent criteria were assumed. If a SNP broke any of the following rules it was removed from the larger summary statistics:

- Longer than a single base pair
- Ambiguous (https://www.snpedia.com/index.php/Ambiguous_flip)
- Did not contain a rsID
- Was not found within the UK Biobank Imputed dataset

| Disease | GWAS First Author | Sample Size | No. SNPs |
|---|---|---|---|
| Lupus | Bentham | 14267 | 5596514 |
| A. Fib. | Christophersen | 133073 | 7596547 |
| Asthma | Demenais | 142486 | 1972099 |
| Celiac Disease | Dubois | 15283 | 510529 |
| Migraine | Gormley | 375752 | 9461477 |
| MS | IMSGC | 34892 | 21940 |
| Vitiligo | Jin | 9735 | 6366969 |
| Gout | Kottgen | 69374 | 2051407 |
| Crohns Disease | Liu-1 | 20883 | 83995 |
| Ulcerative Colitis | Liu-2 | 27432 | 95788 |
| Type 2 Diabetes | Mahajan | 344144 | 44700 |
| Stroke | Malik | 524354 | 6066909 |
| Breast Cancer | Michailidou | 139274 | 6870259 |
| NAFLD | Namjou | 9677 | 2319122 |
| CAD | Nikpay | 187599 | 6528252 |
| Rheumatoid Arthritis | Okada | 79799 | 2078058 |
| Type 1 Diabetes | Onengut | 18856 | 94982 |
| Ovarian Cancer | Phelan | 85426 | 7443207 |
| ALS | Rheenen | 36052 | 6276146 |
| Prostate Cancer | Schumacher | 140254 | 6360250 |
| Heart Failure | Shah | 977323 | 6073796 |
| Psoriasis | Tsoi | 33394 | 55697 |
| Depression | Wray | 142646 | 3254439 |

**Table M2.** The genome wide association studies analyzed throughout this investigation along with their basic meta-statistics, sample size of the analyzed cohort and the number of single-nucleotide polymorphisms in the corresponding summary statistics.

- Was not on chromosome 1-22
- Was removed by the LDSC munge procedure (https://github.com/bulik/ldsc/wiki)
- Standard error was significantly different from the same SNP for complementary FinnGenn summary statistics (https://privefl.github.io/bigsnpr/articles/LDpred2.html)
- Could not be flipped or reversed to match UK Biobank alleles

In addition to the quality control, the alleles were either flipped or reversed to match the UK Biobank alleles by utilizing the snp_match function from the bigsnpr function. Lastly, the summary statistic were broken into a set for each chromosome.

#### Preparation of UK Biobank Genetic Data

Before the UK Biobank imputed data was utilized to adjust summary statistics and create scores, it was carefully quality controlled. The individual-level quality control involved the creation of a custom population sorting algorithm. In short, the 40 genetic principal components were clustered using k-nearest neighbor and each cluster was easily identified as being either Asian, European or African by comparing self-reported ethnicity. Individuals whose self-described ethnicity did not match their cluster label were removed from all downstream analyses. In addition, individuals labeled by the UK Biobank as being hetrozygosity outliers, having sex chromosome aneuploidy, or excess relatives were removed. Lastly, any individuals in a specific scoring or adjustment computation that held more than a 10% missing genotype rate were removed. With respect to genetic variants, any SNPs in a specific scoring or adjustment computation that had an allele frequency less than 0.01 or a Hardy-Weinberg Equilibrium p-value (calculated through the midpoint test) less than $1 \times 10^{-50}$ were removed.

Processing of genetic material, except when required by specific adjustment methods, utilized the bgenix and PLINK utilities. The most efficient, and thereby utilized, combination of these utilities started with bgenix to subset the necessary SNPs, PLINK2 to complete additional QC, and PLINK1.9 to complete scoring, clumping or other computations.

#### Preparation of UK Biobank Phenotypic and Covariate Data

The process of binary labeling of individuals as being either affected or unaffected by a disease of interest started with six data sources and ultimately generated five different labels. The six data sources include non-cancer, cancer-specific and medication self-reports of disease status at time of enrollment in the UK Biobank and ICD10, ICD9 and OPCS codes from hospital inpatient records. If a specified code was present in any of the records then that records was noted as 1, otherwise it would be a 0. In addition, the hospital inpatient records were linked to accurate dates of disease onset. For non-electronic health record data the date of enrollment was considered the date for the disease onset.

From these six vectors of ones and zeros the five phenotype definitions were created by asking whether there was a single one, or multiple ones in a subset of columns. The date of onset was taken to be the minimum date in the subset of columns.

The covariate data was created in a virtually identical manner. However, there was the additional complication of ensuring the covariate occurred before the disease. The date of onset of covariates was therefore considered directly within calling the

| Disease | Sex | Cancer | Non-Cancer | ICD9 | ICD10 | OPCS | Medications |
|---|---|---|---|---|---|---|---|
| Lupus | A | NA | 1381 | 710 | M321|M328|M329 | NA | NA |
| A. Fib. | A | NA | 1471 | 4273 | I48 | K622|K623 | 1140888482 |
| Asthma | A | NA | 1111 | 493 | J45|J46 | NA | 1141168340 |
| Celiac Disease | A | NA | 1456 | 579 | NA | NA | NA |
| Migraine | A | NA | 1265 | 346 | G43 | NA | 1141163670|1141167932| 1141172728|1141185436| 1141192666|1141150620| 1141151284 |
| MS | A | NA | 1261 | 340 | G35 | NA | 1141188640|1141188642| 1141150596|1141172620| 1141165546|1141167618| 1141189254|1141189256 |
| Vitiligo | A | NA | 1661 | 7091 | L80 | NA | NA |
| Gout | A | NA | 1466 | 274 | M10 | NA | 1140875408 |
| Crohns Disease | A | NA | 1463 | 556 | K51 | NA | 1141153242 |
| Ulcerative Colitis | A | NA | 1462 | 555 | K50 | NA | NA |
| Type 2 Diabetes | A | NA | 1223 | NA | E11 | NA | NA |
| Stroke | A | NA | 1081 | 431|432|433| 434|435|436| 437|438 | I60|I61|I62| I633|I64|I65| I66|I67|I68|I69 | U543|Z35 | NA |
| Breast Cancer | F | 1002 | NA | 174 | C50 | B27|B28|B29 | 1140923018|1141190734 |
| NAFLD | A | NA | NA | 5718 | K760 | NA | NA |
| CAD | A | NA | 1075|1076 | 410|411|412 | I21|I22|I23| I241|I252 | K40|K41|K45| K49|K50|K75 | NA |
| Rheumatoid Arthritis | A | NA | 1464 | 714 | M05|M06 | U504 | 1141145896|1140909702| 1141188588|1141180070| 1140871188 |
| Type 1 Diabetes | A | NA | 1222 | NA | E10 | NA | NA |
| Ovarian Cancer | F | 1039 | NA | 183 | C56 | NA | NA |
| ALS | A | NA | NA | 3352 | G122 | X852 | 1141195974 |
| Prostate Cancer | M | 1044 | NA | 185 | C61 | NA | 1141150594|1140870274| 1140921100 |
| Heart Failure | A | NA | 1076 | 428 | I50 | NA | NA |
| Psoriasis | A | NA | 1453 | 6960|6961 | L40 | NA | NA |
| Depression | A | NA | 1286 | NA | F33 | NA | NA |

**Table M3.** The element within each data type used to identify the disease phenotypes. The "|" character represents an "or" operator, in other words that if any of the elements were recorded in the data type the phenotype was considered positive. The sex column identifies whether only one sex was analyzed for a given disease.

| | Self-Reported | ICD | ICD or Self-Reported | Any | Double-Reported |
|---|---|---|---|---|---|
| Non-Cancer | X | | X | X | X |
| Cancer | X | | X | X | X |
| ICD9 | | X | X | X | X |
| ICD10 | | X | X | X | X |
| OPCS | | X | X | X | X |
| Medication | | | | X | X |

**Table M4.** The data types considered for each phenotype definition. For the double-reported definition two or more of the data types had to contain a positive record of the phenotype.

| Disease | Extra Covariates |
|---|---|
| Lupus | sex |
| A. Fib. | age,hypertension,congenital heart disease,cardiac arrest,coronary artery disease,alcohol,sleep apnea |
| Asthma | allergic rhinitis,smoking,bmi |
| Celiac Disease | type 1 diabetes |
| Migraine | age,sex,age menopause,hormone replacement therapy |
| MS | age,sex,epstein barr virus |
| Vitiligo | use of sun protection,melanona,non-hodgkins lymphoma |
| Gout | age,sex,obesity,hypertension,diabetes |
| Crohns Disease | smoking |
| Ulcerative Colitis | smoking |
| Type 2 Diabetes | age,sex,bmi,exercise,hypertension,hypocholesteroemia |
| Stroke | age,sex,bmi,age started oral contraceptive,hypertension,hypocholesteroemia,smoking,alcohol,diabetes |
| Breast Cancer | age,sex,bmi,alcohol,age menarche,age menopause,pregnant |
| NAFLD | age,obesity,hypertension,hypocholesteroemia,diabetes |
| CAD | age,sex,bmi,hypocholesteroemia,hypertension,diabetes,smoking |
| Rheumatoid Arthritis | age,sex,smoking,obesity,pregnant |
| Type 1 Diabetes | none |
| Ovarian Cancer | age,bmi,hormone replacement therapy,pregnant,breast cancer |
| ALS | age,diabetes,obesity |
| Prostate Cancer | age,obesity |
| Heart Failure | cardiac arrest,hypertension,congenital heart defects,obesity,diabetes,arrythmia |
| Psoriasis | smoking |
| Depression | alcohol,hormone replacement therapy,age menopause |

**Table M5.** The extra covariates included in the analysis of each disease

ones and zeros, however this date information was not retained later for future modelling. In the final extra covariate analysis the ICD or Self-Reported definition was used to convert the ones and zeros into a final covariate.

#### Additional Data

Additional data was utilized in various analyses, including external data directly downloaded and data that was computed a priori to downstream analyses. The external data include 1000 Genomes genotypes (https://www.cog-genomics.org/plink/2.0/resources#1kg_phase3, functional annotation boundaries (https://storage.googleapis.com/broad-alkesgroup-public/LDSCORE/baseline_v1.1_bedfiles.tgz, https://storage.googleapis.com/broad-alkesgroup-public/LDSCORE/ct_and_ctg_bedfiles.tgz), and variant severity measures (https://pcingola.github.io/SnpEff/ss_dbnsfp/). Additional data such as external adjusted summary statistics and linkage disequilibrium matrices are specified in their respective analysis section.

The internal data that was generated included heritability values for each measure, generated from LDSC and HDL, and genetic correlations estimated from LDSC. The heritability values were meta-analyzed when two non-NA or non-zero values were available. When not available the single value was utilized.

### Adjusting Summary Statistics

The summary statistics for each chromosome and GWAS were adjusted using various methods for the purpose of generating polygenic risk scores. Therefore, these methods will henceforth be referred to as generative methods. The process of adjusting

| Covariate | UKBB Phenotype Col. | ICD10 | ICD9 | Non-Cancer | Cancer |
|---|---|---|---|---|---|
| age | 34-0.0 | NA | NA | NA | NA |
| sex | 31-0.0 | NA | NA | NA | NA |
| hypertension | NA | I10 | 401 | 1065,1072 | NA |
| congenital_heart_disease | NA | Q24 | 746 | NA | NA |
| cardiac_arrest | NA | I21,I46 | 410,4275 | NA | NA |
| coronary_artery_disease | NA | I25 | 414 | NA | NA |
| alcohol | 1558-0.0 | NA | NA | NA | NA |
| sleep_apnea | NA | G473 | NA | 1123 | NA |
| allergic_rhinitis | NA | J30 | 477 | 1387 | NA |
| smoking | 20116-0.0 | NA | NA | NA | NA |
| bmi | 21001-0.0 | NA | NA | NA | NA |
| type_1_diabetes | NA | E10 | 250 | 1222 | NA |
| exercise | 884-0.0 | NA | NA | NA | NA |
| hypocholesteroemia | NA | E780 | NA | 1473 | NA |
| age_started_oral_contraceptive | 2784-0.0 | NA | NA | NA | NA |
| age_menarche | 2714-0.0 | NA | NA | NA | NA |
| age_menopause | 3581-0.0 | NA | NA | NA | NA |
| pregnant | 2754-0.0,3140-0.0 | NA | NA | NA | NA |
| hormone_replacement_therapy | 2814-0.0 | NA | NA | NA | NA |
| breast_cancer | NA | C50 | 2330 | NA | 1002 |
| epstein_barr_virus | 23053-0.0 | NA | NA | NA | NA |
| use_of_sun_protection | 2267-0.0 | NA | NA | NA | NA |
| diabetes | NA | E10,E11 | 250 | 1220,1222,1223 | NA |
| arrhythmia | NA | I47,I48 | 427 | 1077 | NA |
| sle | NA | M32 | 7100 | 1381 | NA |
| melanoma | NA | C43 | 172 | NA | 1059 |
| non-hodgkins_lymphoma | NA | C82,C83 | NA | NA | 1053 |

**Table M6.** The UK Biobank codes used to determine each extra covariate. The ICD and self-reported codes were pulled very similar to the normal phenotypes, and if any of these data sources recorded the phenotype it was assumed to be correct, similar to the any phenotype method. The UKBB Phenotype Col. refers to the UID as designated by the UK Biobank.

| Disease | ldsc_h2 | ldsc_h2_se | hdl_h2 | hdl_h2se | h2 |
|---|---|---|---|---|---|
| Lupus | 0.6196 | 0.0829 | 0.3376 | 0.0421 | 0.433 |
| A. Fib. | 0.1127 | 0.019 | 0.0827 | 0.0188 | 0.0976 |
| Asthma | 0.096 | 0.0117 | 0.0397 | 0.0092 | 0.0645 |
| Celiac Disease | 0.2938 | 0.0527 | 0 | 0 | 0.294 |
| Migraine | 0.0447 | 0.0032 | 0.0341 | 0.0027 | 0.039 |
| MS | 1.3517 | 0.2212 | 0 | 0 | 0.01 |
| Vitiligo | 0.9885 | 0.2083 | 0.7602 | 0.084 | 0.826 |
| Gout | -0.1573 | 0.0995 | 0 | 0 | 0.01 |
| Crohns Disease | 13.0787 | 1.8819 | 0.1917 | 0.0432 | 0.192 |
| Ulcerative Colitis | 6.2819 | 0.6996 | 0.0879 | 0.0235 | 0.0879 |
| Type 2 Diabetes | 0.1713 | 0.0212 | 0 | 0 | 0.171 |
| Stroke | 0.0281 | 0.0027 | 0.0117 | 0.0023 | 0.0192 |
| Breast Cancer | 0.2184 | 0.0181 | 0.2342 | 0.0216 | 0.226 |
| NAFLD | 0.5875 | 0.2227 | 0 | 0 | 0.588 |
| CAD | 0.0734 | 0.0049 | 0.0497 | 0.0045 | 0.061 |
| Rheumatoid Arthritis | 0.5941 | 0.0366 | 0.0397 | 0.0154 | 0.204 |
| Type 1 Diabetes | 4.002 | 0.8874 | 0.0549 | 0.0213 | 0.0549 |
| Ovarian Cancer | 0.0546 | 0.0114 | 0.0543 | 0.0095 | 0.0544 |
| ALS | 0.0552 | 0.0138 | 0.0308 | 0.0073 | 0.0392 |
| Prostate Cancer | 0.1423 | 0.0215 | 0.1049 | 0.0149 | 0.12 |
| Heart Failure | 0.0424 | 0.0035 | 0.0379 | 0.0028 | 0.0399 |
| Psoriasis | 0.9485 | 0.2299 | 0 | 0 | 0.948 |
| Depression | 0.1298 | 0.0096 | 0.0449 | 0.0031 | 0.0656 |

**Table M7.** Computed heritabilities from all cleaned summary statistics. The colum names ldsc refers to the Linkage Disequilibrium Score Regression method and hdl refers to the High-Definition Likelihood method.

requires the preparation of UK Biobank, and possibly additional data, the computation that does the adjustment, and lastly data-clean up that leaves a well-formatted summary statistic file. For brevity, only the second step in this process is described as the other two are trivial and repeated. As most generative methods require a number of hyperparameters, a grid of such parameters is defined and therefore more than just one set of adjusted summary statistics is generated per method and disease. The generative methods utilized and their descriptions of just the adjustment step follow.

### Clumping

Implementation of the clumping method was applied through the PLINK software. The "-—clump" flag was selected with a series of "-—clump-p1" p-value and "-—clump-p2" R2 thresholds. The variants in the output file were used to subset the primary summary statistics.

Official Documentation: `https://www.cog-genomics.org/plink/1.9/postproc`

## WC-2d

Implementation of the WC-2d (winners curse two dimensions) method roughly followed the steps outlined in the clumping method, except for specification of which regions clumping applies to. Specifically, variants identified as being conserved (listed within `http://compbio.mit.edu/human-constraint/data/gff/`) and pleiotropic (listed within supplementary table 2 of the respective publication) were both clumped with a higher p-value threshold. Only one of these varieties of variants

| Name | Time to Run (s) | Model Based? | Ease to Implement | Description |
|---|---|---|---|---|
| Clumping[1] | 3, 19, 71 | No | 1 | Greedy Selection of SNPs by P-Value in LD Region |
| Double Weight[2] | 3, 26, 74 | No | 2 | Reduce Winners Curse with Empirical Beta Distribution |
| WC-2D[3] | 36, 97, 266 | No | 2 | Clumping with thresholds based on functional annotation |
| WC-Likelihood[3] | 24, 199, 802 | No | 2 | Reduce Winners Curse via Thresholding Likehood Stat |
| WC-Lasso[3] | 5, 28, 0113 | No | 2 | Reduce Winners Curse with LASSO-like penalty |
| Tweedie[4] | 8, 44, 203 | No | 2 | Threshold Efron's Statistic Created via Kernel Method |
| lassosum[5] | 27, 188, 579 | No | 2 | LD-Aware Penalization over effect distribution |
| LDPred[6] | 1463, 12640, 30846 | Yes | 3 | Bayesian Estimate of LD-Aware Multiple Regression |
| LDPred2[7] | 37, 280, 1001 | Yes | 3 | LDPred, improved speed and numeric stability |
| prsCS[8] | 548, 4264, 10507 | Yes | 3 | Bayesian Estimate with a Continous Prior of Effect |
| SBLUP[9] | 176, 1604, 4326 | Yes | 3 | Approximation of multiple regression BLUP-style effects |
| DBSLMM[10] | 43, 368, 602 | Yes | 3 | Deterministic Bayesian Sparse Linear Mixed Model |
| SBayesR[11] | 5, 30, 87 | Yes | 4 | Approximation of Bayesian multiple regression framework |
| JAMPred[12] | 160, 1370, 1406 | Yes | 4 | Flexible, Bayesian Adjustment of Variable LD |
| SMTPred[13] | 14, 18, 21 | Partly | 3 | Leverages correlated trait effect estimates |

**Table M8. The polygenic risk score generative methods**. The time to run, measured in seconds, was evaluated by running each method on 10,000, 100,000 and 250,000 SNPs from the first chromosome of the atrial fibrillation summary statistics. Model-based refers to methods that attempts to recover predictions that would be constructed from full knowledge of the genetic information. An ease to implement value of 1 indicates a near-minimum level of data manipulation, 2 indicates minor supplementary coding required, 3 indicates the added organization of multiple program inputs/outputs, and 4 indicates extra tuning requiring to assure method convergence. Additional method descriptions are provided in the Methods section.

were allowed to have a different threshold at a time.

### WC-lasso

To implement WC-lasso (winners curse lasso), the original summary statistic file was first clumped with a p-value cut-off of 0.01 and R2 cut-off of 0.1. The vector of remaining effect sizes was then modified according to the following equation that was extracted from the original publication.

$$\hat{\beta}_m^{lasso} = sign(\hat{\beta}_m)\left|\hat{\beta}_m\right| - \lambda I\left(\left|\hat{\beta}_m\right| > \lambda\right)$$

The lambda value ranged from 0.001 to 0.1, and the output effect sizes were directly extracted without additional modification.

### WC-likelihood

The WC-likelihood (winners curse likelihood) method was implemented similar to WC-lasso, with the same initial clumping step. The following effect adjustment step however required minimizing a likelihood function. The full function is available in the originating publication. Computationally, the minimization was accomplished by the Python function "minimize" and the "nelder-mead" method.

#### Double Weight

The Double Weight method, was implemented very similarly to the WC-likelihood and WC-lasso methods, even though it is not formally a member of the WC family. After the initial clumping, the effects and standard errors were read into a custom written R script that simulated many samples of effects for each variant. Then a variant was selected to be in the final adjusted summary statistics if it was in the top range of SNPs with a specified probability. The value for the range of SNPs varied.

#### Tweedie

Implementation of the Tweedie method began by an application of the clumping method, following the steps as described in the original publication. For the initial clumping step the p-value threshold was set at 0.05 and R2 at 0.25. The modified summary statistic file was then used within the main tweedie R script that minimized a likelihood function similar to WC-likelihood. In order to extract the beta values the published script was modified slightly, and is located at https://github.com/kulmsc/PRS-Ithaca/blob/master/tweedy.R. Three variations of the betas were created, one for each of the FDR, Tweedie, and FDR x Tweedie sub-methods.

Official Documentation: https://sites.google.com/site/honcheongso/software/empirical-bayes-risk-prediction

#### LDpred

To implement the LDpred method the starting summary statistic file was first converted into the STANDARD format required by LDpred (columns of chromosome, position, reference allele, alternative allele, reference allele frequency, info, rsID, p-value, and effect of the alterative allele). Generating this file simply required reorganizing the starting summary statistic file. The LD range was calculated as the number of SNPs divided by 4500. The "ldpred coord" step was first run, followed by a series of "ldpred gibbs" applications with various "f" values (the proportion of true causal variants).

Official Documentation: https://github.com/bvilhjal/ldpred

#### LDpred2

The implementation of LDpred2 directly followed the vignette that accompanied the original publication. While the theory was nearly identical to LDpred, all of the coding was done in R. Additional hyperparameters were fit to investigate a greater number of possible fractions of causal SNPs and heritabilities.

Official Documentation: https://privefl.github.io/bigsnpr/articles/LDpred2.html

#### lassosum

Implementation of lassosum essentially required a single function call with UK Biobank used as reference data. The lassosum.pipeline function generated the adjusted effects with minimal intervention. A grid of "s" and "lambda" parameters were tried directly within this pipeline.

Official Documentation: https://github.com/tshmak/lassosum

#### PRScs

Implementation of the PRScs first required converting the format of the summary statistics. The primary computation was encapsulated in the single python prscs call. The reference data employed was the European LD data listed within the PRScs documentation. The phi parameter was changed over three different function calls, whereas the a and b parameter were held steady.

Official Documentation: https://github.com/getian107/PRScs

#### sBLUP

Implementation of the sBLUP method began by down sizing the original summary statistic to only the variants included within the HapMap (https://www.broadinstitute.org/medical-and-population-genetics/hapmap-3), and the columns were re-arranged to the MA format used throughout the GCTA tool kit. The actual sblup option was then run within gcta, with the wind option (the LD distance parameter) set to 100.

Official Documentation: https://cnsgenomics.com/software/gcta/#SBLUP

#### SBayesR

Implementation of the SBayesR method started similarly to sBLUP, with conversion of the summary statistics to the necessary MA format. The primary SBayesR computations were then run within the gctb toolkit. The ldm files were generated from the UK Biobank and down sized to HapMap variants, and could be downloaded directly from the documentation. Ldm files with more variants were not used due to size limitations.

Official Documentation: https://cnsgenomics.com/software/gctb/#SummaryBayesianAlphabet

#### DBSLMM

Additional information was first derived, such as allele frequencies from a PLINK call applied to the UK Biobank data. Implementation of the DBSLMM algorithm was completed easily within a single R function call. Once generated p-value and R-squared hyperparameters were iterated over, and the adjusted effect sizes directly computed.

Official Documentation: https://github.com/biostat0903/DBSLMM

#### SMTpred

Implementation of SMTpred is unique in that it required not just the primary summary statistics being adjusted, but also summary statistics of similar diseases. The exact set of similar summary statistics are determined a priori through genetic correlation calculations. After proper formatting of the primary and similar summary statistics, they are all entered into a single python function call that adjusts the effect sizes. The number of similar sets of summary statistics varied. In addition, SBLUP adjusted effects are also utilized in this process.

Official Documentation: https://github.com/uqrmaie1/smtpred

#### JAMPred

Implementation of the JAMPred algorithm is analogous to LDpred2 as both chiefly complete their computations within R. However, JAMPred is far less readily able to handle the large genotypic matrices necessary. Therefore, LD pruning in PLINK was first carried out, to reduce the size of the genotypic files. Next, the genotypic data is read into R using the bignsnpr package, and is then carefully converted into the necessary matrix specifications while still being in a memory-efficient format. The primary JAMPred function is then called over several iterations of lambda values, generating the adjusted effect sizes.

Official Documentation: https://github.com/pjnewcombe/R2BGLiMS

### Creating Polygenic Risk Scores

With the original GWAS summary statistics adjusted under various generative methods and respective hyperparameters, polygenic risk scores could be created. This computation was easily accomplished by using the "–score" option within PLINK1.9, and by following the genetic data processing workflow previously described. The "sum" option was included in the PLINK call to prevent normalization before the polygenic risk score for each chromosome were added together. To mitigate possible allele and variant mismatching, one genotypic file was created for each score that was needed. While this specificity slowed down the scoring, it appeared to reduce round-off error.

### Tuning to the Best Score

The best generative method and set of respective hyperparameters for each disease was determined by tuning. This process computed several statistics for each polygenic risk score within a cross-validation framework. Specifically, the tuning process began with only 60% of all British individuals who passed QC. Then three folds were iterated through in which two thirds of the data was used for training and the remaining third for testing. These folds were themselves repeated three times, shifting the start of the index that determined the three groups by one ninth of the total population. This method is commonly called repeated cross fold validation.

The training and testing groups within each iteration were used to fit and assess models, creating statistics that judged both total model fit and the ability to stratify individuals at the tail of risk. These statistics were first computed under a survival mode of the analysis. The survival process started by creating the data frame that contains the start and end times for each individual. All individuals were assumed to have entered the study January 1st 1999, the earliest time that we could judge all individuals had reliable electronic health record data. Any health event listed before this time was removed. The end time is either the individual's time of respective disease diagnosis, the individual's date of death, the individual's date of study removal request, or the last available date of electronic health records (May 31st 2020). A Fine and Gray model adjustment was made following the methods in the survival package, leaving anything other than the date of diagnosis as a censored time. From the survival data frame a Fine and Gray model was fit with covariates of age, sex and the top ten genetic principal components. This model is referred to as the base model (if the disease was sex-specific, for example breast cancer, sex was not included and only the specific sex was analyzed). Then a new model was generated that additionally included each of the polygenic risk scores as a covariate, referred to as the score models. All statistics were generated for both of these models in identical fashion. The concordance was determined through the "survConcordance" function with the test data set as an argument. The final cumulative hazard was determined for a high, intermediate and low risk score group by a simple application of the cox proportional hazard equation at the final time available. The groups were defined according to the lowest and highest quintiles of hazards being selected as the high and low risk groups, respectively.

Similar to the survival analysis, binary analyses were completed to determine the predictive performance in terms of both model fit and at the tail of risk. The survival dataframe was simply collapsed into a binary dataframe that did not include any date information. Logistic regression base and score models were fit. Predictions from these models upon the test set was compared to the true test labels to determine the area under the receiver operator curve (AUC), specifically using the pROC library. The predictions were then used to create exposed or non-exposed groups. The non-exposed group was fixed as individuals whose prediction was less than the 20th percentile value of the entire prediction vector. The exposed group varied with the upper percentile used as a cut-off. The exposed groupings and the test labels were then used to construct a contingency table and from there the odds ratio.

Once all folds were complete the statistics were averaged over the nine folds. The score model that generated the highest AUC was selected to be the best score for the given disease. To give a better depiction of all models, the best score for each model and disease was maintained for later analysis and plotting.

### Testing the Predictive Performance

For each disease, the polygenic risk score determined to be the best in the tuning section was included in models that were used to predict the witheld 40% of data. The specific fitting and prediction process was largely identical to the tuning process. The major difference was that instead of having cross-validation, the models were directly fit upon 60% of the data used in the tuning section, and predicted upon the full 40% of withheld data. The four statistics, concordance, cumulative hazard, AUC and odds ratios were generated for score and base models in an identical fashion.

Along with the processes replicated from the tuning section a few additional analyses were conducted. First, disease-specific covariates were included within both the base and score models to create extra-base and extra-score models. The predictions from these models were used to compute AUCs, odds ratios, and other statistics described in the following paragraph. Second, other scores that were externally derived were analyzed by exchanging the polygenic risk score determined to be the best in the tuning phrase by each of the external scores. The AUCs, odds ratios, and other statistics were again computed for these other models. All of the generated statistics were included in plots for each specific disease, and across all diseases.

In addition to the statistics generated in the tuning section other statistics were included to the testing analysis to better evaluate how well the polygenic risk score improves the base model. First, precision-recall curves were computed directly by applying the test set predictions and labels to the "pr.curve" function. Second, net reclassification improvement and integreated

| Disease | Score Name | First Author | Pub. Year |
|---|---|---|---|
| Lupus | PGS000196 | Knevel | 2020 |
| A. Fib. | PGS000016 | Khera | 2018 |
| Asthma | PGS000037 | Belsky | 2013 |
| Gout | PGS000199 | Knevel | 2020 |
| Type 2 Diabetes | PGS000014 | Khera | 2018 |
| Type 2 Diabetes | PGS000020 | Lall | 2016 |
| Type 2 Diabetes | PGS000036 | Mahajan | 2018 |
| Stroke | PGS000039 | Abraham | 2019 |
| Breast Cancer | PGS000007 | Mavaddat | 2018 |
| Breast Cancer | PGS000015 | Khera | 2018 |
| Breast Cancer | PGS000045 | Kuchenbaecker | 2017 |
| Breast Cancer | PGS000052 | Lakeman | 2019 |
| Breast Cancer | PGS000072 | Graff | 2020 |
| CAD | PGS000011 | Tada | 2015 |
| CAD | PGS000013 | Khera | 2018 |
| CAD | PGS000058 | Morieri | 2018 |
| Rheumatoid Arthritis | PGS000194 | Knevel | 2020 |
| Rheumatoid Arthritis | PGS000195 | Knevel | 2020 |
| Ovarian Cancer | PGS000082 | Graff | 2020 |
| Prostate Cancer | PGS000044 | Pashayan | 2015 |
| Prostate Cancer | PGS000067 | Seibert | 2018 |
| Prostate Cancer | PGS000084 | Graff | 2020 |

**Table M9.** The other, external scores available from the PGS Catalog (https://www.pgscatalog.org/browse/all/) that were compared to the internally constructed scores

discrimination improvement was computed by comparing the score-included and base model through the reclassification function within the PredictABEL package. The net reclassification improvement was specifically of the categorical type with the same cut-offs used to compute the odds ratio employed here. Third, decision curves were constructed by comparing the phenotype to predictions from the base and score-included models. The options cohort and opt-in were set within the decision_curve function of the rmda package. Fourth, the number of individuals reclassified was calculated for each cut-off used to compute the odds ratios. Specifically, the intersection of individuals within the high risk group (above the cut-off) as defined by the score-included and base models was calculated then compared to the total number of individuals in the high risk group. Fifth, true positive rates within the high risk group (individuals with a risk greater than the cut-off used in the odds ratio calculations) were calculated by simply dividing those with disease by the total size of the risk group, as all individuals in this group were assumed to be predicted positive. Lastly, the true positive and false positive rates over all individuals were extracted from the ROC curve at the point where one minus the true positive rate minus the false positive rate was a minimum.

### Translation Analyses

The first part of our translation analyses involved a decision curve analysis. The computations required to generate decision curves have already been described in the previous paragraph.

The two following sections employed a common methodological system that interrogated lifestyle factors and medications/supplements. First, we approximately split each lifestyle factor into 3 groups that roughly fall into the 1st, 2nd-4th, and

| Lifestyle Factor | Low Risk Group | Intermediate Risk Group | High Risk Group |
|---|---|---|---|
| Alcohol Consumed | >1 per day | 1-4 per week | <1 per week |
| Smoking Status | Never | Previous | Current |
| Days Mod. Activity Per Week | <2 | 2-5 | >5 |
| Hours per Day Watch TV | <3 | 3 | >3 |
| Hours per Day Driving | 0 | 1 | >1 |
| Hours Sleep | <7 | 7 | >7 |
| BMI | <23.5 | 23.5-30.6 | >30.6 |
| Min. Walked Per Day | <20 | 20-80 | >80 |
| Walking Pace | Slow | Average | Brisk |
| Processed Meat Intake | <1 per week | 1 per week | >1 per week |
| Glasses of Water Per Day | <2 | 2-3 | >3 |
| Cheese Intake | <1 per week | 1 per week | >1 per week |
| Pieces Fruit Eaten Per Day | <2 | 2 | >2 |
| Tbsp Raw Veg. Per Day | <2 | 2 | >2 |

**Table M10.** The cut-offs used to define the lifestyle measure low, intermediate and high risk groups

5th quintile. Similarly, the PRS adjusted for age, sex and the top ten genetic principal components were split into 3 groups that again fall into the 1st, 2nd-4th, and 5th quintiles. The absolute risk was computed for nine groups of individuals defined by the intersections of the three lifestyle factor groups and three PRS risk groups. Following previous work, the absolute risk was approximated by the net incidence, or the number of events in the time of the study. In addition to the net incidence calculations, a fisher exact test was computed on all of the three PRS groups where the predicted group had the high lifestyle measure and non-predicted group had the low lifestyle measure. A disease and lifestyle modification was considered significant if the absolute risk reduction brought by the modification was greater in the high compared to low PRS group, and either all of the fisher p-values were less than 0.05, any two p-values were than 0.005, or any one p-value was less than 0.0005.

The methodology used to analyze the lifestyle modifications was largely replicated for the medication and supplement analysis. Except this time the groupings were simply individuals that either self-reported taking or not taking the respective medication or supplement. Therefore, the fisher exact test did not pull from the high and low risk groups but rather just the two present risk groups. The same significant thresholds were also used. In both the lifestyle and medication analyses the net incidence was computed from phenotypes that were only reported from electronic health record data gathered after the primary time of assessment.

### Checking Polygenic Risk Score Validity

The validity of all polygenic risk scores were checked by generally stratifying the scores by various features and checking to see whether performance, or some other measure, remained the same in each group. In all of these validity tests the polygenic risk scores utilized in the testing phase were the focus, although the best score for each disease and generative method combination was also included for method-specific analyses. First, sex was used to stratify the training (60% of the entire dataset) and testing (40% of the entire dataset) into male or female specific groups. Models were trained, fit, and AUCs derived on each of these groups.

| Disease | Age Split |
|---|---|
| Lupus | 70.47 |
| A. Fib. | 75.63 |
| Asthma | 69.96 |
| Celiac Disease | 71.47 |
| Migraine | 68.46 |
| MS | 67.96 |
| Vitiligo | 71.63 |
| Gout | 73.8 |
| Crohns Disease | 71.96 |
| Ulcerative Colitis | 70.71 |
| Type 2 Diabetes | 73.8 |
| Stroke | 75.22 |
| Breast Cancer | 72.13 |
| NAFLD | 70.96 |
| CAD | 74.8 |
| Rheumatoid Arthritis | 73.3 |
| Type 1 Diabetes | 72.63 |
| Ovarian Cancer | 73.3 |
| ALS | 74.3 |
| Prostate Cancer | 75.38 |
| Heart Failure | 75.96 |
| Psoriasis | 70.71 |
| Depression | 68.46 |

**Table M11.** The ages at which the population was split into young and old groups for each disease

Second, age was used for stratification. For each disease the age that divided cases into equally sized groups was used to define young and old groups. The specific ages determined in this manner are listed in table M13. Same as for the sex stratification, models were fit and AUCs derived.

Third, socially relevant variables were used in series to carry out the same style of stratified analysis. These variables included time at current address (split at 20 years), income (split at £40,000), number in household (split at 2), and age finished final education (split at 19 years).

Fourth, census variables were similarly used to carry out the same style of stratified analysis. The census data was obtained by linking the reported home location of each individual (accurate to within 1 km) to the individual's local super output area. The census data for that local super output area, obtained from the United Kingdom Office for National Statistics, was then ascribed to that individual. The specific census features utilized include median age (split at 42 years), unemployment (split at 38), very good health measure (split at 719), and population density (split at 32 persons per hectare).

Fifth, the polygenic risk score distribution within population groups were compared. Specifically, African, Asian, Non-British European, and British individuals' scores were grouped and their means and standard deviations compared. The method for defining these population groups was described in the Preparation of UK Biobank Phenotypic and Covariate Data section.

Sixth, the predictive accuracy of various disease labels were compared. Specifically, all of the phenotype definitions described in table M4 were substituted into the typical analysis data frame used throughout the testing process. However, only the AUC measure from these new definitions were recorded.

Seventh, the average AUC across all of the folds in the tuning section and the AUC computed within the test section for each score was compared. While not an analysis of validity akin to the other checks, the training/tuning comparison was completed and results grouped by method, disease and overall best score to ensure that overfitting did not occur in the larger polygenic risk score analysis process.

### Finding Patterns to Aid in Polygenic Risk Score Development

Various analyses were completed in the effort to form empirically-backed, polygenic risk score development recommendations. The first such analysis started with the method ranking system used throughout this investigation. Specifically, the AUC rankings that determined the best score for each method that was used in all downstream analyses. The ranking was modified to find the best method for a given GWAS summary statistic attribute by multiplying the AUC by the attribute after each was normalized to range from 0 to 1. The methods ranked in the top 10 by these attribute weighted AUCs were considered to be well-performing under the attribute and simply counted. The specific attributes considered include the sample size or number of individuals, number of SNPs less than 1e-8 or the number of SNPs with an associated p-value less than $1 \times 10^{-8}$, number of SNPs less than 1e-6 or the number of SNPs with an associated p-value less than $1 \times 10^{-6}$, the number of SNPs, heritability as reported in table M7, and case control ratio or the number of individuals with the disease divided by those without. In addition there were two distribution metrics including distribution by length and effect. These metrics are computed by ordering the variants by their absolute effect then splitting the variants into four groups such that the sum of the absolute effect in each group is equal. The metric for length then divides the mean length in the top effect group by the mean length in the bottom group, and the metric for effect divides the mean effect in the top effect group by the mean effect in the bottom group. Lastly all of the attributes used in these rankings were reversed such that a value of 1 becomes 0 and 0 becomes 1.

Second, we continued in the general path of connecting polygenic risk score attributes to general performance. Specifically, by first quantifying the number of variants that generated each polygenic risk score, and secondly generating the distribution metrics as defined in the first overall analysis.

Lastly, SNPs from the best polygenic risk score for each disease was mapped to a function annotation group. Once, the SNPs were assigned to multiple, one, or none functional annotation groups, a simple average of the absolute effect was computed for each group. This average was then normalized by dividing by setting the range of functional annotation scores for each disease range from 0 to 1, instead of the raw computed score. This was process was repeated, but with deleterious scores, and the normalization process involved the product of the deleterious score and variant's absolute weight. The functional annotations were pulled from https://alkesgroup.broadinstitute.org/LDSCORE/ and the deleterious scores originated at https://pcingola.github.io/SnpEff/ss_dbnsfp/.

| Disease | sig6_snps | sig8_snps | len_pdiff | effect_pdiff | sample_size | snps | h2 | cc_ratio |
|---|---|---|---|---|---|---|---|---|
| Lupus | 0.293 | 0.245 | 0.62 | 0.000168 | 0.00474 | 0.591 | 0.45 | 0.429 |
| A. Fib. | 0.0844 | 0.0544 | 0.315 | 0.000597 | 0.128 | 0.802 | 0.0934 | 0.0983 |
| Asthma | 0.0693 | 0.0521 | 0.927 | 2.17e-05 | 0.137 | 0.207 | 0.0581 | 0.135 |
| Celiac Disease | 0.00298 | 0.00153 | 1 | 0 | 0.00579 | 0.0518 | 0.302 | 0.309 |
| Migraine | 0.0267 | 0.0204 | 0 | 1 | 0.378 | 1 | 0.0308 | 0.125 |
| MS | 0.116 | 0.122 | 0.306 | 0.000621 | 0.0261 | 0 | 0 | 0.404 |
| Vitiligo | 0.0535 | 0.0402 | 0.6 | 0.000183 | 5.99e-05 | 0.672 | 0.869 | 0.0368 |
| Gout | 0.0163 | 0.0121 | 0.339 | 0.000534 | 0.0617 | 0.215 | 0 | 0 |
| Crohns Disease | 0.322 | 0.282 | 0.534 | 0.00024 | 0.0116 | 0.00657 | 0.194 | 0.291 |
| Ulcerative Colitis | 0.212 | 0.156 | 0.605 | 0.000179 | 0.0183 | 0.00782 | 0.083 | 0.245 |
| Type 2 Diabetes | 0 | 0.000612 | 0.156 | 0.00149 | 0.346 | 0.00241 | 0.172 | 0.22 |
| Stroke | 0.0316 | 0.0151 | 0.678 | 0.00013 | 0.532 | 0.64 | 0.00985 | 0.0914 |
| Breast Cancer | 0.885 | 0.722 | 0.441 | 0.000347 | 0.134 | 0.725 | 0.23 | 0.931 |
| NAFLD | 0.00223 | 0.00312 | 0.675 | 0.000132 | 0 | 0.243 | 0.615 | 0.0772 |
| CAD | 0.134 | 0.0858 | 0.534 | 0.000239 | 0.184 | 0.689 | 0.0544 | 0.359 |
| Rheumatoid Arthritis | 1 | 1 | 0.331 | 0.000553 | 0.0725 | 0.218 | 0.207 | 0.226 |
| Type 1 Diabetes | 0.1 | 0.0735 | 0.635 | 0.000157 | 0.00949 | 0.00774 | 0.0478 | 0.41 |
| Ovarian Cancer | 0.109 | 0.116 | 0.0159 | 0.0166 | 0.0783 | 0.786 | 0.0473 | 0.171 |
| ALS | 0.00349 | 0.00343 | 0.684 | 0.000127 | 0.0273 | 0.663 | 0.0312 | 0.399 |
| Prostate Cancer | 0.665 | 0.484 | 0.00811 | 0.0325 | 0.135 | 0.671 | 0.117 | 1 |
| Heart Failure | 0.0202 | 0.0105 | 0.639 | 0.000155 | 1 | 0.641 | 0.0319 | 0.0154 |
| Psoriasis | 0.091 | 0.0873 | 0.197 | 0.00112 | 0.0245 | 0.00358 | 1 | 0.342 |
| Depression | 0.0185 | 0 | 0.471 | 0.000308 | 0.137 | 0.342 | 0.0593 | 0.344 |

**Table M12.** The metrics computed then normalized for each distribution that were utilized in the weighted method rankings

### References


1. Chang, C. C. *et al.* Second-generation PLINK: rising to the challenge of larger and richer datasets. *GigaScience* **4**, 7, DOI: 10.1186/s13742-015-0047-8 (2015).

2. Läll, K., Mägi, R., Morris, A., Metspalu, A. & Fischer, K. Personalized risk prediction for type 2 diabetes: the potential of genetic risk scores. *Genet. Medicine* **19**, 322–329, DOI: 10.1038/gim.2016.103 (2016).

3. Shi, J. *et al.* Winner's Curse Correction and Variable Thresholding Improve Performance of Polygenic Risk Modeling Based on Genome-Wide Association Study Summary-Level Data. *PLOS Genet.* **12**, e1006493, DOI: 10.1371/journal.pgen.1006493 (2016).

4. So, H.-C. & Sham, P. C. Improving polygenic risk prediction from summary statistics by an empirical Bayes approach. *Sci. Reports* **7**, 1–11, DOI: 10.1038/srep41262 (2017).

5. Mak, T. S. H., Porsch, R. M., Choi, S. W., Zhou, X. & Sham, P. C. Polygenic scores via penalized regression on summary statistics. *Genet. Epidemiol.* **41**, 469–480, DOI: 10.1002/gepi.22050 (2017).

6. Vilhjálmsson, B. J. *et al.* Modeling Linkage Disequilibrium Increases Accuracy of Polygenic Risk Scores. *Am. J. Hum. Genet.* **97**, 576–592, DOI: 10.1016/j.ajhg.2015.09.001 (2015).

7. Privé, F., Arbel, J. & Vilhjálmsson, B. J. LDpred2: better, faster, stronger. *Bioinformatics* DOI: 10.1093/bioinformatics/btaa1029 (2020).

8. Ge, T., Chen, C.-Y., Ni, Y., Feng, Y.-C. A. & Smoller, J. W. Polygenic prediction via Bayesian regression and continuous shrinkage priors. *Nat. Commun.* **10**, 1–10, DOI: 10.1038/s41467-019-09718-5 (2019).

9. Robinson, M. R. *et al.* Genetic evidence of assortative mating in humans. *Nat. Hum. Behav.* **1**, DOI: 10.1038/s41562-016-0016 (2017).

10. Yang, S. & Zhou, X. Accurate and scalable construction of polygenic scores in large biobank data sets. *The Am. J. Hum. Genet.* **106**, 679–693, DOI: 10.1016/j.ajhg.2020.03.013 (2020).

11. Lloyd-Jones, L. R. *et al.* Improved polygenic prediction by Bayesian multiple regression on summary statistics. *Nat. Commun.* **10**, 1–11, DOI: 10.1038/s41467-019-12653-0 (2019).

12. Newcombe, P. J., Nelson, C. P., Samani, N. J. & Dudbridge, F. A flexible and parallelizable approach to genome-wide polygenic risk scores. *Genet. Epidemiol.* **43**, 730–741, DOI: 10.1002/gepi.22245 (2019).

13. Maier, R. M. *et al.* Improving genetic prediction by leveraging genetic correlations among human diseases and traits. *Nat. Commun.* **9**, 1–17, DOI: 10.1038/s41467-017-02769-6 (2018).
